# Past dynamics of HIV transmission among men who have sex with men in Montréal, Canada: a mathematical modelling study

**DOI:** 10.1101/2021.08.29.21262800

**Authors:** Rachael M Milwid, Yiqing Xia, Carla Doyle, Joseph Cox, Gilles Lambert, Réjean Thomas, Sharmistha Mishra, Daniel Grace, Nathan J Lachowsky, Trevor A Hart, Marie-Claude Boily, Mathieu Maheu-Giroux

## Abstract

**Background:** Gay, bisexual, and other men who have sex with men (gbMSM) experience disproportionate risks of HIV acquisition/transmission. In 2017, Montréal became the first Canadian Fast-Track city, setting the 2030 goal of zero new HIV infections. To inform local elimination efforts, we estimate the evolving role of prevention/risk behaviours and HIV transmission dynamics among gbMSM in Montréal between 1975-2019.

**Methods:** Data from local bio-behavioural surveys were analyzed to develop, parameterize, and calibrate an agent-based model of sexual HIV transmission. Partnership dynamics, the HIV natural history, and treatment and prevention strategies were considered. The model simulations were analyzed to estimate the fraction of HIV acquisitions/transmissions attributable to specific age-groups and unmet prevention needs.

**Results:** The model-estimated HIV incidence peaked in 1985 (2.2%; 90%CrI: 1.3-2.8%) and decreased to 0.1% (90%CrI: 0.04-0.3%) in 2019. Between 1990-2017, the majority of HIV acquisitions/transmissions occurred among men aged 25-44 years, and men aged 35-44 thereafter. The unmet prevention needs of men with >10 annual anal sex partners contributed 92-94% of transmissions and 63-73% of acquisitions annually. The primary stage of HIV played an increasing role over time, contributing to 12%-27% of annual transmissions over 1990-2019. In 2019, approximately 75% of transmission events occurred from men who had discontinued, or never initiated ART.

**Conclusions:** The evolving HIV landscape has contributed to the recent low HIV incidence among MSM in Montréal. The shifting dynamics identified in this study highlight the need for continued population-level surveillance to identify unmet prevention needs and core groups on which to prioritize elimination efforts.

## Introduction

From the beginning of the HIV epidemic, gay, bisexual, and other men who have sex with men (gbMSM) have been disproportionately impacted^1^. In 2019, 60% of new diagnoses in Québec were recorded in Montréal, Canada’s second most populated city^2^. Of the new diagnoses, 70% occurred among men, of which 71% were reported among gbMSM^3^.

In 2017, Montréal became the first Canadian Fast-Track city, pledging to reach zero new HIV infections by 2030^4^. The successful achievement of this ambitious goal largely depends on the implementation of effective public health initiatives to reduce the acquisition and transmission potential for HIV. Understanding the impact of sexual behaviors and specific HIV interventions on the epidemic can aid in the development of relevant HIV elimination plans^5^. This objective is complicated, however, by the continued evolution of the HIV prevention and treatment landscape, which includes: HIV testing (1985)^6^, anti-retroviral therapy (ART; 1996)^7^, post-exposure prophylaxis (PEP; 2001)^8^, and pre-exposure prophylaxis (PrEP; 2013)^9^.

Characterizing HIV epidemics and their complex transmission dynamics is difficult using traditional epidemiological methods. Tools such as agent-based models (ABM) –computer simulations of epidemics– are appropriate to understand and characterize past and future transmission dynamics^10^. Mathematical models of HIV transmission have been used to describe the drivers (e.g., age, infection stage, knowledge of status, and treatment status^11,12^) and dynamics of global and local HIV epidemics. These models have focused on different population subgroups, have been applied to a range of geographical locations, and have studied, both prospectively and retrospectively, the impact of various interventions on HIV dynamics^13-16^. The relevance and applicability of the models is largely dependent on the quality and quantity of the data used to parameterize and inform the population-specific model^17^.

Despite the large quantity of available Montréal-based sexual behavioral data, there have been no recent attempts to systematically analyze and triangulate data and surveys on HIV transmission among gbMSM in Montréal to understand evolving transmission dynamics. The purpose of this study was to describe the epidemiological characteristics of the HIV epidemic and understand sources of HIV acquisition and transmission between 1975-2019.

## Methods

### Data analysis

Data from the peer-reviewed and grey literature, and individual-level data from the two cross-sectional *Argus* surveys (2005; n=1,957 and 2008; n=1,873), the *Engage* cohort (2017; n=1,179, 2018; n=887, 2019; n=850), and the *L’Actuel PrEP Cohort* (2013-2019; n=2746) studies were analyzed to identify temporal trends in sexual behaviors and HIV intervention uptake rates. Details on the studies can be found elsewhere^18-24^. Briefly, the *Argus* surveys used venue-based convenience sampling to recruit gbMSM aged 18+ years from the island of Montréal who reported sex with another man during their lifetime. The longitudinal *Engage* cohort used respondent-driven sampling to recruit gbMSM aged 18+ years from Montréal who reported sex with another man within the past six months. The *Argus* data were standardized (by age and ethnicity) to the *Engage* data which were analyzed using Volz-Heckathorn weights^25^. The data analysis was used to inform model development, parameterization, and calibration (see *Supplementary Materials*).

### Agent based model

An ABM was developed to simulate in discrete time, the sexual transmission dynamics of HIV among gbMSM aged 15+ years in Montréal. The model represents an open population, matching the demographic and epidemiological characteristics reported in the surveys and health and census data. A detailed model description can be found in the *Supplementary Materials*. The main characteristics of the partnership formation, natural disease progression, and HIV interventions are described below. A complete list of model parameters (demographic, sexual behaviors, biological, and interventions) is presented in the *Supplementary Materials*.

### Partnership formation and duration

Upon entry into the population, men are assigned an annual partner change rate, informed by the survey data. Only partnerships in which anal sex is performed are modelled. As individuals age, their partner change rate increases slightly up to age 30 and then decreases, in line with the survey data. Men having ≤5, 6-10, and >10 anal sex partners per year are categorized as being in the low, medium, and high sexual partnering groups, respectively^26^. Men are assigned a partner based on data-driven mixing matrices that vary by age, perceived serostatus, and preferred anal sex role (insertive, receptive, or versatile). If no available partners match all three criteria, the search continues with either age or serostatus dropped. If there are still no suitable partners, the man does not form a partnership at that time.

Men can form casual or regular partnerships. Casual partnerships begin and end within the same time step (but can involve more than one sex act), while the duration of regular partnerships is assumed to be longer than one month. Men can only have one concurrent regular partnership. Therefore, upon partnership formation, if either man is already part of a regular partnership, then the partnership is assigned a casual status. Otherwise, a Bernoulli distribution is used to determine the partnership type.

### HIV transmission and progression

It is assumed that there are no external sources of sexual HIV transmission (e.g., in the context of sex with female partners or sharing injecting needles). Following HIV acquisition, individuals go through an initial short, highly infectious period^27^. Subsequently, people living with HIV (PLHIV) progress through distinct CD4 cell count categories, with some starting at a lower CD4 cell count^28^: >500 cells *µL*^−1^, 350-500 cells *µL*^−1^, 200-350 cells *µL*^−1^, and <200 cells *µL*^−1^. Men on ART maintain their CD4 cell count from treatment initiation until ART discontinuation or death^29^. The AIDS-related death rate varies by age, stage of infection, treatment status, and time on ART^30,31^.

The probability of HIV transmission within sero-discordant partnerships is computed as a function of the number of sex acts in the partnership, the HIV-positive partner’s stage of infection and ART status, and the HIV-negative partner’s sex role, PrEP use, the proportion of sex acts protected by condoms within the partnership, and whether the man seeks PEP afterward.

### HIV interventions

All men have similar HIV testing rates prior to 1996. Subsequently, the annual rates of HIV testing were estimated from survey data and were allowed to vary by sexual partnering levels (low versus moderate/high). Once tested for HIV, a minimum inter-testing interval is assigned. From 1985 onwards, testing occurs for undiagnosed symptomatic PLHIV on average six months after progressing to the late stage of infection, irrespective of calendar time.

Upon HIV diagnosis, eligible men are assigned an ART initiation date. The local time-varying ART eligibility criteria is incorporated in the model such that PLHIV with a CD4 cell count <350 cells *µL*^−1^ (1996-2013), <500 cells *µL*^−1^ (2013-2016), and all PLHIV (2016 onwards) are eligible for ART. The effectiveness of ART in reducing HIV transmission is assumed to be 80% for the first 3-6 months after initiation^32^, and 90-100% thereafter^33,34^ as not all those on ART will achieve sustained viral suppression^35^. Men can discontinue and later reinitiate ART. Following ART discontinuation, PLHIV continue to progress through lower CD4 cell count categories.

Survey data regarding the frequency of condom use were used to estimate the per-partner and per-act probability of using condoms. The categorical survey responses (never, occasionally, often, and always) for each stratum (partnership type, age, sero-status, sexual partnering level, and time) were converted into a numerical equivalent and averaged. An all-or-nothing approach is implemented for men in casual partnerships, while regular partners may protect only a proportion of their sex acts with condoms. Condoms are assumed to be 91% efficacious in preventing HIV transmission^36^.

PEP, which is assumed to be 80% efficacious in preventing HIV acquisition^37^, became available for non-occupational use for gbMSM at the end of 2000. Given the low PEP uptake at that time^8^, PEP was initiated in the model in 2001 and uptake rates increased linearly until 2017. PEP eligibility is modelled according to local clinical protocols.

In 2013, Québec recommended PrEP and partly reimbursed it using public funds^8^. The PrEP eligibility criteria incorporated within the model is consistent with local guidelines^38^ and includes post-initiation HIV testing. PrEP is assumed to be 86% effective in inhibiting HIV acquisition^39^. A detailed description of the PEP and PrEP model components is presented in the *Supplementary Materials*.

#### Model calibration

An approximate Bayesian computation-sequential Monte Carlo algorithm was used to select the parameter sets that best reproduce the following epidemiological features: age-stratified HIV prevalence (2005, 2008, 2017-2019), ART coverage (2005, 2017-2019), the proportion who ever used PrEP (2017-2019), the PrEP coverage (2017-2019), the CD4 cell count at diagnosis (2013-2017), and the annual proportion diagnosed with HIV (2005, 2008, 2017). Additional cross-validation was performed by comparing the model outputs with surveillance data on: HIV incidence, the proportion of PLHIV with knowledge of status (2018), the proportion of PLHIV who used PrEP in the past six months (2017-2019), the proportion of PLHIV with viral load suppression (2017-2019), and the age-stratified CD4 cell count at diagnosis (2013-2017)^40^, and number of new diagnoses (2002-2017)^40^. The model calibration resulted in 50 posterior parameter sets. A full description of the calibration process can be found in the *Supplementary Materials*.

The model was initialized in 1975 and progressed in two-week increments. A total of 10,000 agents are simulated and results are scaled-up to the estimated population size of gbMSM. Model simulations were run once for each posterior parameter set. The model was coded in R, version 3.6.3^41^, and incorporated C++ through the Rcpp interface, version 1.0.4.6^42^

#### Model analysis

##### Description of the HIV epidemic

To capture the salient features of the epidemic, the HIV dynamics are characterized from 1975-2019. The annual prevalence, incidence, and mortality rates are computed. To identify the groups most affected by HIV, each measure is stratified by age group (15-24, 25-34, 35-44, 45-54, and 55+ years) and sexual partnering level (low, medium, and high).

##### Drivers of the HIV epidemic

The drivers of the epidemic are calculated from 1990 onwards, following the epidemic peak observed during the calibration process. Model outcomes are calculated from the beginning of the respective calendar year until the end of the year. We examined both who acquired (Eq. 1) and who transmitted infections (Eq. 2). To characterize who acquired infections (*A*_*ti*_), we divided the number of men with characteristic *i* (e.g., age, sexual partnering level) who acquired HIV over a defined period by the cumulative number of new HIV infections (*I*_*t*_) within that time period (*t*_0_ → *t*_*n*_).

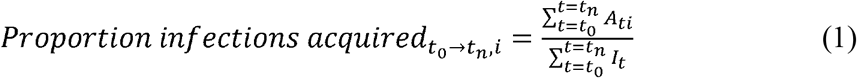

The proportion of incident infections transmitted was computed by dividing the cumulative number of transmission events *(T*_*ti*_) from PLHIV with characteristic *i* (i.e., age, sexual partnership level, infection stage, knowledge of HIV status, ART status) over a defined time period (*t*_0_ → *t*_*n*_) by the cumulative number of new infections (*I*_*t*_) within that time period (Eq. 2).

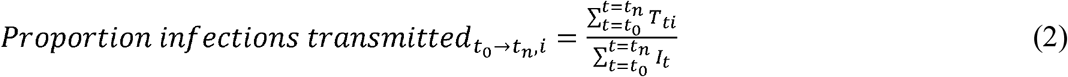

The mean and 90% credible interval (CrI), calculated as the 5^th^ and 95^th^ percentiles, of the 50 simulations are reported for each measure.

#### Ethics

This study was approved by McGill University’s Faculty of Medicine and Health Sciences Institutional Review Board (A12-E84-18A).

## Results

The calibrated model adequately replicated the age-specific epidemiological outcomes and intervention coverage (*Supplementary Materials*), providing credence in the ability of the model to capture population-level HIV transmission dynamics over time.

### Description of the epidemic

The simulated HIV incidence peaked in 1985 (2.2% year^-1^, 90%CrI: 1.3-2.8%) and decreased to 0.1% (90%CrI: 0.04-0.3%) in 2019 (Figure 1). Throughout the epidemic, the incidence among the 35-44-year age group was 2-3 times that of the 15-24-year age group.

**Figure 1:**
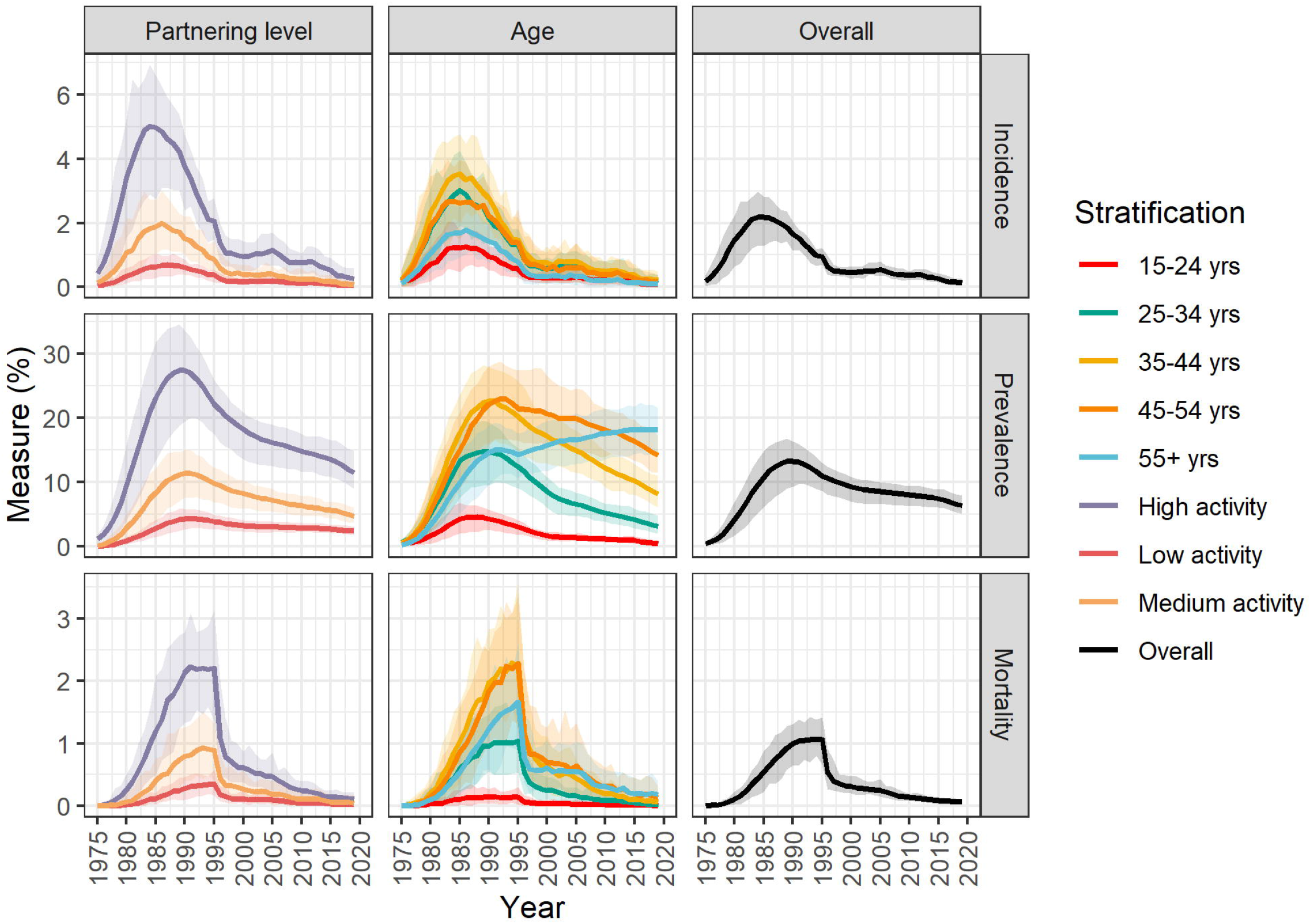
Overall, age and sexual partnering level stratified estimates of HIV incidence, prevalence, and mortality among men who have sex with men in Montréal over 1975-2019. The solid line represents the mean of 50 simulations, and the shaded area represents the 90% credible interval. The HIV incidence peaked in 1985, while the prevalence and mortality peaked in 1990 and 1993, respectively. The 35-44-year age group and high sexual partnering group had the highest prevalence and incidence at the epidemic peak, while the 45-54-year age group had the highest mortality. By 2017, the incidence was still highest within the 35-44-year age group. Subsequent to 1990, the HIV prevalence decreased for all age groups, with the exception of the 55+ year age group for which the prevalence continues to increase.

Additionally, the incidence ratio between the high and low sexual partnering groups ranged from 10-11 between 1975-1981, 8-10 between 1981-1985, and 5-7 thereafter, while the incidence ratio between the high and medium sexual partnering groups ranged between 2-4 throughout the epidemic.

The annual HIV prevalence peaked circa 1990 at 13.3% (90%CrI: 9.3-16.4%) (Figure 1). During this time, the prevalence was highest among men aged 35-44 years (22.6%), and those in the high sexual partnering group (27.4%). Following this 1990 peak, the prevalence decreased for all strata, excluding men aged 55+ years who continue to experience an increase in prevalence. By 2019, the overall prevalence had decreased to 6.3% (90%CrI: 5.1-7.9%), and the oldest age group and high sexual partnering group had the highest prevalence at 18.1% and 11.5%.

HIV-specific mortality rates peaked in 1993 at 1.1% (90%CrI: 0.7-1.4%). The mortality rate was highest among the 45-54-year age group, up to 10 times that of the 15-24-year olds from 1975-1988 and 11-25 times greater thereafter. From 1978-1987, the death rate among men in the high sexual partnering group was 9-24 times greater than that of the low sexual partnering group, and 4-8 times greater from 1986 onwards. The HIV-related mortality rate among the high sexual partnering group was 2-4 times that of the medium sexual partnering group from 1978 onwards.

### Drivers of the HIV epidemic

Over 1990-2019, the proportion of infections attributed to the primary stage of HIV infection increased from 12% (90%CrI: 7-19%) to 27% (90%CrI: 0-58%) (Figure 2). The proportion of incident infections attributed to the late stage of disease decreased from 29% (90%CrI: 19-37%) to 12% (90%CrI: 0-35%).

**Figure 2:**
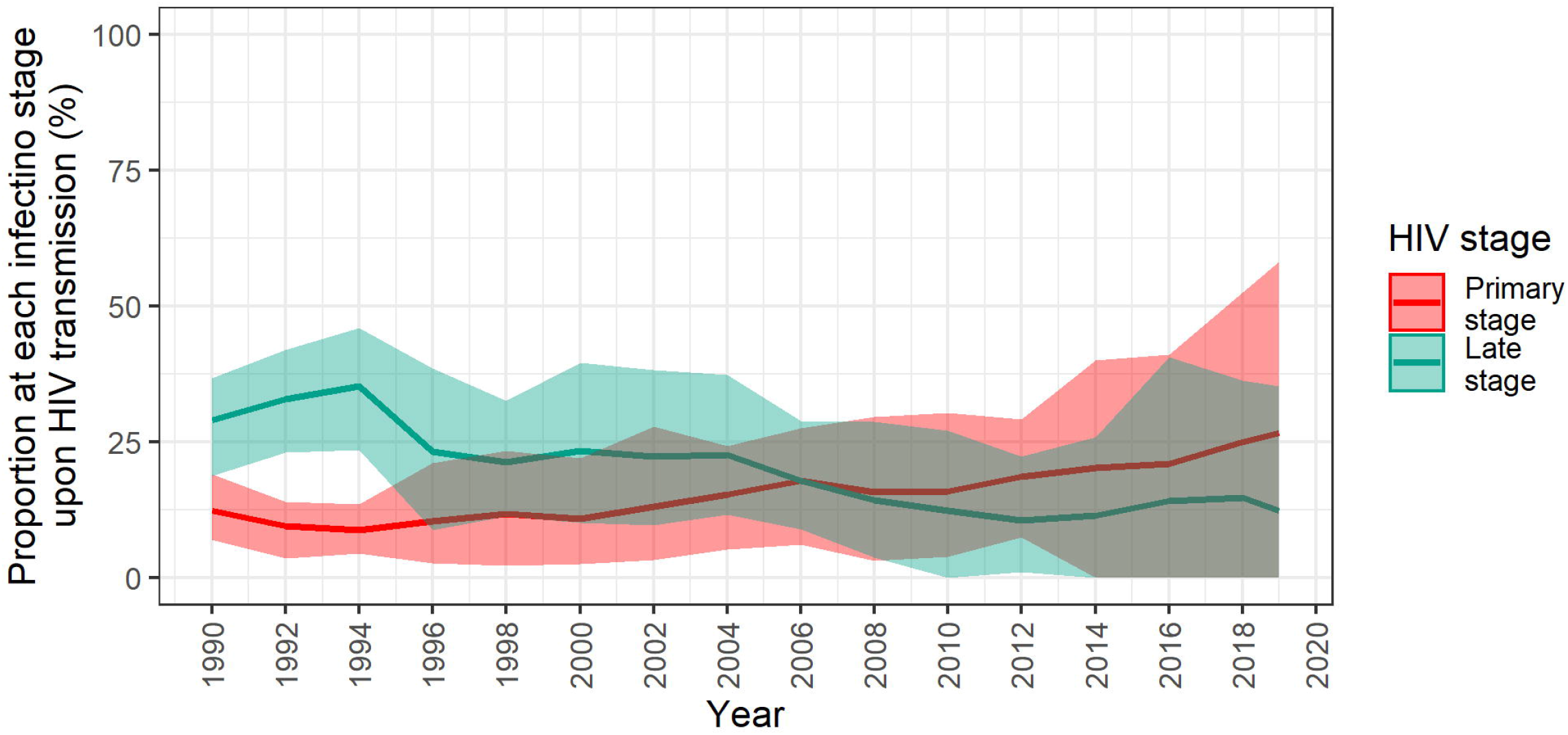
Proportion of annual HIV transmission events from individuals in the primary and late stages of infection. The mean (solid line) and 90% credible interval (shaded region) of 50 simulations are shown. The proportion of infections from men at the primary stage of HIV infection increased over time from a mean of 12% in 1990 to 27% in 2019. In contrast, men at the late stage of infection played a decreasing role in new infections.

Within a given year, 16-30% and 70-84% of transmission events occurred within regular and casual partnerships respectively (Figure 3). Regarding age, the majority of infections were transmitted and acquired by individuals aged 25-34 years from 1990 to 2017, and those aged 35-44 thereafter (Figure 4a). HIV transmissions and acquisitions among men aged 25-34 years peaked in 2008 and decreased until 2019. Over 1990-2019, the 35-44-year age group played an increasing role in HIV transmissions (28-38%), and acquisitions (24-38%). Finally, the high partnering group transmitted (92-94%) and acquired (63-73%) most new infections throughout the epidemic. Of note, while the low/medium partnering groups each transmitted <7% of infections, they acquired up to 20% of new HIV infections (Figure 4b).

**Figure 3:**
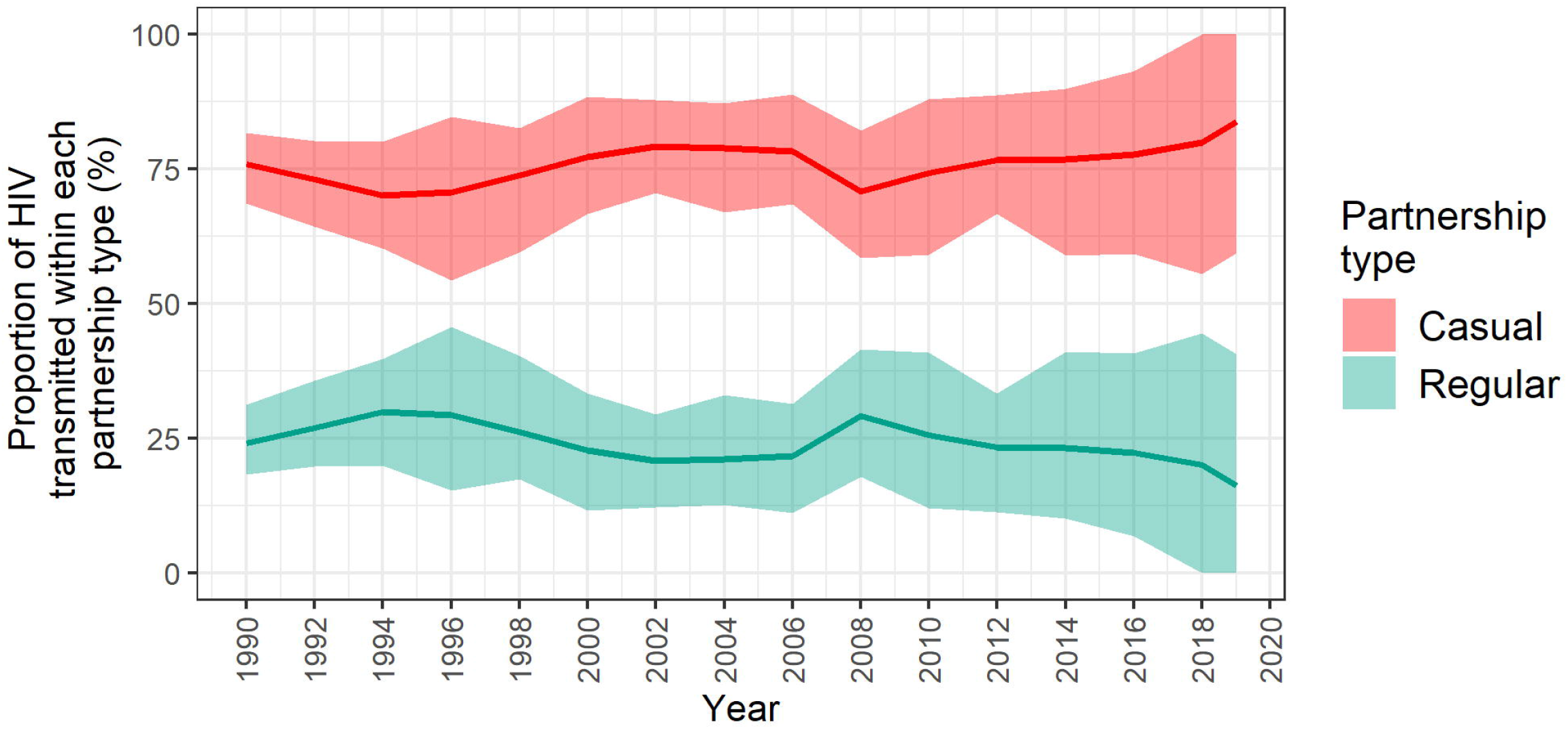
Proportion of annual incident HIV infections transmitted among casual and regular partners. Regular partnerships were defined as having a duration ≥1 month. The mean proportion of transmissions (solid line) among each partnership type and the 90% credible interval (shaded region) are shown. Approximately 25% of new infections were transmitted within regular partnerships, while 75% of new infections were transmitted within casual partnerships.

**Figure 4:**
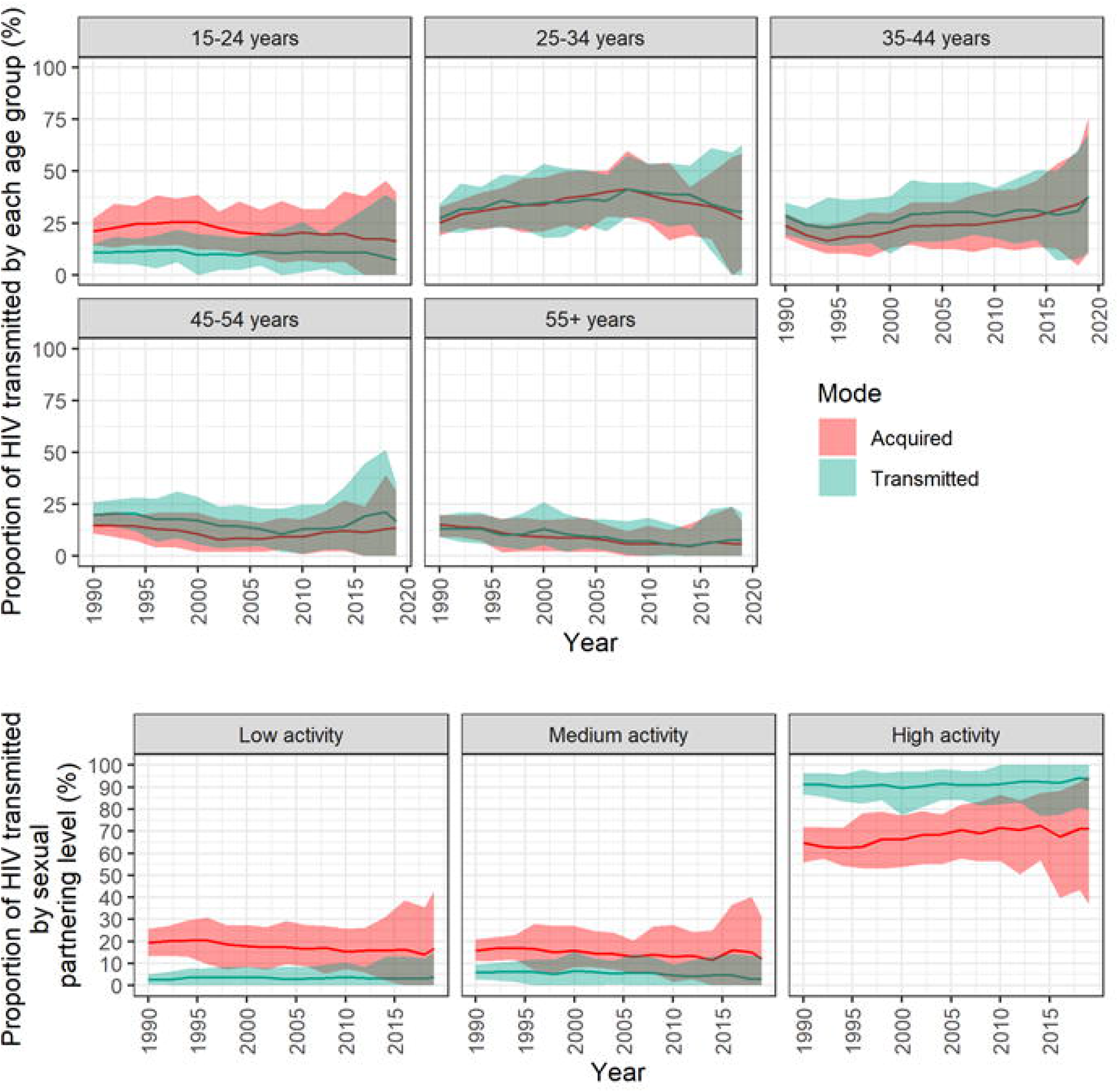
Mean and 90% credible interval of annual proportion of new infections transmitted and acquired by people living with HIV (PLHIV) stratified by age group (panel A) and sexual partnering level (panel B). Most new infections were transmitted and acquired by PLHIV aged 25-34 years between 1990 and 2017, and men aged 35-44 thereafter. With respect to sexual partnering level, through the epidemic, the majority of infections were transmitted by those with more than 10 partners per year.

The proportion of gbMSM unaware of their HIV status decreased from 74% (90%CrI: 71-77%) in 1990 to 4% (90%CrI: 2-6%) in 2019. Between 1996-2019, the proportion who had never initiated ART decreased from 63% (90%CrI: 55-89%) to 5% (90%CrI: 3-7%), while the proportion who had discontinued ART for any length of time at the time of HIV transmission decreased from 5% (90%CrI: 1-8%) to 1% (90%CrI: 0-3%). The proportion of transmission events from those unaware of their HIV-positive status decreased from 67% (90%CrI: 59-76%) in 1990 to 50% (90%CrI: 20-84%) in 2019. Over 1996-2019, the majority of transmission events occurred among PLHIV who had never been on ART. This proportion dropped from 96% to 60%, while the proportion of transmission events among men who had either discontinued ART or were on treatment (without a suppressed viral load) increased from 2% and 2% to 14% and 26% (Figure 5).

**Figure 5:**
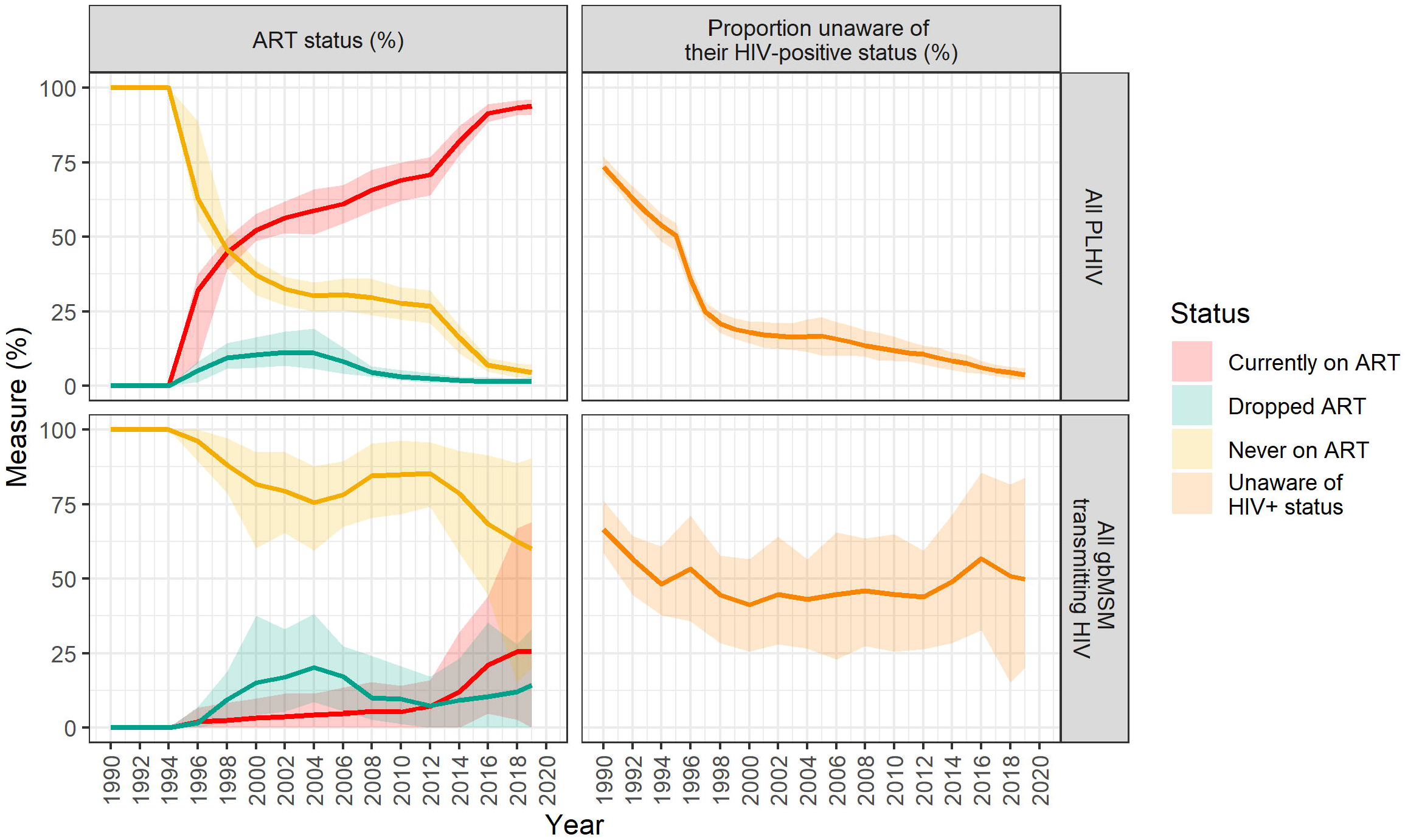
Proportion of people living with HIV (PLHIV) and those transmitting HIV who were unaware of their HIV-positive status, and their antiretroviral therapy (ART) status. The solid line represents the mean of 50 simulations and the shaded band represents the 90% credible interval. The proportion of PLHIV unaware of their status decreased from 74% in 1990 to 4% in 2019, while the equivalent proportion among those transmitting HIV decreased from 67% to 50% between 1990 and 2019. The proportion of PLHIV who had never been on ART decreased from 63% to 5% between 1996 and 2019 while the proportion who had never been on ART at the time of transmitting HIV decreased from 96% to 60%. gbMSM: gay, bisexual, and other men who have sex with men

## Discussion

Achieving city-level HIV elimination requires a granular understanding of local past and present transmission dynamics, the identification of unmet prevention needs, and data-driven implementation of appropriate interventions among the populations most impacted by HIV^43^. Through the development and analysis of a detailed model of HIV dynamics among gbMSM in Montréal, we characterized trends in HIV incidence, prevalence, mortality, as well as the individual characteristics of HIV acquisitions and transmissions. Our model points to important improvements in epidemiological and continuum of care outcomes, with low HIV incidence in recent years.

At the beginning of the epidemic, the rising mortality rates between 1985-1995, combined with some saturation of groups with higher partnering levels, played an important role in reducing the proportion of individuals who could contribute to onward HIV transmission. The latter translated into a decreasing HIV incidence and subsequent fall in prevalence over 1990-2000. The introduction of ART in 1996 lead to a sharp drop in mortality, consistent with national estimates of deaths due to AIDS^1^. With the advent of these life-saving treatments and concomitant incidence reductions, the AIDS epidemic aged^44^. This was particularly salient with the increasing HIV prevalence among men aged 55+ years. This trend is similar to the national one in Canada where, despite an overall increase in the number of PLHIV, the number of gbMSM PLHIV has stabilized^45^. Nationally, other groups of individuals such as people who inject drugs, Indigenous, and other heterosexual males and females contribute to increasing national prevalence.

The large incidence ratio between the high and low sexual partnering groups is indicative of the underlying mixing patterns within the population and the increased likelihood of HIV transmission and acquisition among individuals with more sexual partners^46^. Consequently, the high partnering group experienced a greater reduction in incidence after the introduction of ART. The low/medium sexual partnering groups acquired approximately 20% of new HIV infections but transmitted few. This highlights their smaller role in ongoing transmission of HIV but suggests that renewed HIV prevention efforts that engage men in the low/medium sexual partnering groups are needed to further reduce HIV acquisition. Improving PrEP use among eligible men (e.g., those not using condoms) in these lower sexual activity groups could address such prevention needs. In addition, the model simulations indicate that the majority of transmission and acquisition events took place among gbMSM aged 25-44. This result is in line with local HIV surveillance data of new HIV diagnoses^3^ and should be considered as we approach HIV elimination.

Our analyses identified opportunities to strengthen HIV prevention. Although we evaluate that diagnosis coverage has achieved high levels (>90%) in recent years, approximately half of HIV transmission events in 2019 could have occurred from undiagnosed men. With the current low HIV incidence and high HIV testing rates, this undiagnosed fraction has considerably decreased over the last decade. ART coverage among gbMSM in Montréal was also estimated to be high in 2019 (>90%). However, not all men on ART have a suppressed viral load and our model suggests that close to 1 out 4 transmission events could be from PLHIV on treatment. There is a short 3-6 month period following ART initiation where viral load suppression has not been achieved and clinical data of gbMSM in Montréal suggest that 93% of those on ART are virally suppressed ^35^. Reflecting this setting of low HIV incidence, high ART coverage, and low AIDS incidence, our model suggests that those in the acute infection stage have contributed a relatively larger proportion of transmission events over the last decade. These estimates are comparable with estimates found among gbMSM in the UK (population attributable fraction: 3-28% in 2014-15)^15^ and Baltimore (8-35% over 2008-17)^11^. Given the heightened HIV transmissibility during the primary stage of infection^47^, identifying new clusters of infections through the routine use of recency assays^48^, phylogenetic testing^49^, and genetic sequencing^50^ could help achieve HIV elimination. Additionally, continued efforts are necessary to increase timely diagnosis, prompt linkage to ART, the continued promotion of “U=U” (i.e., undetectable equals untransmittable)^51^ to reduce stigma, and sustaining viral load suppression among those who have achieved it.

The study results should be interpreted considering several limitations. First, the paucity of recorded data from the early years of the epidemic (e.g., condom use, sexual behaviors, and HIV testing) necessitated the use of informed assumptions. Although the calibrated model resulted in good fits, we were unable to cross-validate our earlier inferences with empirical data. Further, the inherent biases of self-reported data (e.g., sex role preference), could affect some of our results. Second, the survey instruments had different eligibility criteria and, in some cases, metrics had slightly different definitions across surveys. However, statistical methods were used to standardize survey data, ensuring their comparability and improving representativeness. Third, although ethnicity was not incorporated in the model, studies suggest that in Canada, there are minimal differences in HIV prevalence by race^52^ although we recognize that some minoritized populations could face distinct barriers. Finally, the lower HIV incidence in recent years resulted in large uncertainty when performing stratified analyses.

Despite achieving low HIV incidence, Montréal gbMSM continue to account for the majority of all new HIV diagnoses in Québec^3^. To achieve the goal of HIV elimination, continued surveillance efforts and the strategic implementation of combination HIV prevention strategies, including increased frequency of HIV testing among high sexual partnering individuals, and increased PrEP among all PLHIV, should be prioritized. By characterizing the HIV dynamics, it was possible to identify the population subgroups most impacted by HIV and the need to further improve ART retention. The implementation of interventions that focus on gbMSM aged 25-45, and gbMSM in the high partnering group could help to achieve HIV elimination among gbMSM in Montréal.

## Data Availability

The data used in this work is not publicly available

## Acknowledgements

R.M.M and M.M-G developed the model structure in consultation with M-C.B, Y.X., C.D., J.C, S.M., and G.L. Y.X and C.D performed the data analysis. R.M.M and M.M-G drafted the manuscript, which was edited by Y.X, C.D, J.C, G.L, R.T, S.M, D.G, T.A.H., N.J.L, and M-C.B. The supplementary appendix was drafted by R.M.M, Y.X and C.D. and edited by Y.X, C.D, J.C, G.L, R.T, S.M, D.G, T.A.H., N.J.L, and M-C.B.

## List of supplementary digital content

Supplementary Digital Content: supplementary materials.docx

## References

1. Public Health Agency of Canada. Chapter 2: Population-Specific HIV/AIDS status report: People living with HIV/AIDS - epidemiological profile of HIV and AIDS in Canada. 2015.

2. Statistics Canada. Municipalities in Canada with the largest and fastest-growing populations between 2011 and 2016. 2017.

3. Institut national de santé publique du Québec. Programme de surveillance de l’infection par le virus de l’immunodéficience humaine (VIH) au Québec: rapport annuel 2019. 2020.

4. Montreal sans sida. Common action plan, 2019-2020; Summary. 2019.

5. Rotheram-Borus MJ, Swendeman D, Chovnick G. The past, present, and future of HIV prevention: integrating behavioral, biomedical, and structural intervention strategies for the next generation of HIV prevention. Annu Rev Clin Psychol. 2009;5:143–167.

6. Paz-Bailey G, Mendoza MC, Finlayson T, et al. Trends in condom use among MSM in the United States: the role of antiretroviral therapy and seroadaptive strategies. AIDS. 2016;30(12):1985–1990.

7. Hogg RS, Heath K, Lima VD, et al. Disparities in the burden of HIV/AIDS in Canada. PLoS One. 2012;7(11):e47260.

8. Xia Y, Greenwald Z, Milwid R, et al. Pre-exposure prophylaxis uptake among men who have sex with men who used nPEP: A longitudinal analysis of attendees at a large sexual health clinic in Montréal (Canada). JAIDS Journal of Acquired Immune Deficiency Syndromes. 2020;85:408–415.

9. Xia Y, Greenwald ZR, Milwid RM, et al. Pre-exposure prophylaxis uptake among men who have sex with men who used non-occupational post-exposure prophylaxis: a longitudinal analysis of attendees at a large sexual health clinic in Montreal (Canada). J Acquir Immune Defic Syndr. 2020.

10. Willem L, Verelst F, Bilcke J, Hens N, Beutels P. Lessons from a decade of individual-based models for infectious disease transmission: a systematic review (2006-2015). BMC Infect Dis. 2017;17(1):612.

11. Silhol R, Boily MC, Dimitrov D, et al. Understanding the HIV epidemic among MSM in Baltimore: a modelling study estimating the impact of past HIV interventions and who acquired and contributed to infections. J Acquir Immune Defic Syndr. 2020.

12. Goodreau SM, Rosenberg ES, Jenness SM, et al. Sources of racial disparities in HIV prevalence in men who have sex with men in Atlanta, GA, USA: a modelling study. The Lancet HIV. 2017;4(7):e311–e320.

13. Baggaley RF, Garnett GP, Ferguson NM. Modelling the impact of antiretroviral use in resource-poor settings. PLoS Med. 2006;3(4):e124.

14. Law MG, Prestage G, Grulich A, Van de Ven P, Kippax S. Modelling the effect of combination antiretroviral treatments on HIV incidence. Aids. 2001;15(10):1287--1294.

15. Punyacharoensin N, Edmunds WJ, De Angelis D, et al. Modelling the HIV epidemic among MSM in the United Kingdom: quantifying the contributions to HIV transmission to better inform prevention initiatives. AIDS. 2015;29(3):339–349.

16. Sorensen SW, Sansom SL, Brooks JT, et al. A mathematical model of comprehensive test-and-treat services and HIV incidence among men who have sex with men in the United States. PLoS One. 2012;7(2):e29098.

17. Garnett PG. An introduction to mathematical models in sexually transmitted disease epidemiology. Sexually transmitted infections. 2002;78(1):7–12.

18. Public Health Agency of Canada. M-Track: Enhanced surveillance of HIV, sexually transmitted and blood-borne infections, and associated risk behaviours among men who have sex with men in Canada. Phase 1 report. 2011.

19. Public Health Agency of Canada. Survey of HIV, viral hepatitis, STIs and associated risk behaviours among Quebec men who have sex with men. 2009; http://argusquebec.ca/english/index.html. Accessed 2019-080-06.

20. Lambert G, Cox J, Miangotar Y, et al. ARGUS 2008-2009 : Enquête sur l’infection par le VIH, les hépatites virales et les infections transmissibles sexuellement et par le sang (ITSS) ainsi que sur les comportements à risques associés chez les hommes québécois ayant des relations sexuelles avec des hommes. 2011.

21. Lambert G, Cox J, Messier-Peet M, Apelian H, Moodie E, Engage research team. Engage Montréal, Portrait of the sexual health of men who have sex with men in Greater Montréal, Cycle 2017-2018, Highlights. 2019.

22. Doyle CM, Maheu-Giroux M, Lambert G, et al. Combination HIV prevention strategies among Montreal gay, bisexual, and other men who have sex with men in the PrEP era: A latent class analysis. AIDS Behav. 2020.

23. Laniece Delaunay C, Cox J, Klein M, et al. Trends in hepatitis C virus seroprevalence and associated risk factors among men who have sex with men in Montreal: results from three cross-sectional studies (2005, 2009, 2018). Sex Transm Infect. 2020.

24. Greenwald ZR, Maheu-Giroux M, Szabo J, et al. Cohort profile: l’Actuel pre-exposure prophylaxis (PrEP) cohort study in Montreal, Canada. BMJ Open. 2019;9(6):e028768.

25. Volz E, Heckathorn DD. Probability based estimation theory for respondent driven sampling. Journal of official statistics. 2008;24:79.

26. Wong NS, Kwan TH, Tsang OTY, et al. Pre-exposure prophylaxis (PrEP) for MSM in low HIV incidence places: should high risk individuals be targeted? Sci Rep. 2018;8(1):11641.

27. Grace D, Steinberg M, Kwag M, et al. Diagnostic technologies in practice: gay men’s narratives of acute or recent HIV infection diagnosis. Qual Health Res. 2015;25(2):205–217.

28. Cori A, Pickles M, van Sighem A, et al. CD4+ cell dynamics in untreated HIV-1 infection: overall rates, and effects of age, viral load, sex and calendar time. AIDS. 2015;29(18):2435–2446.

29. Sucharitakul K, Boily MC, Dimitrov D, Mitchell KM. Influence of model assumptions about HIV disease progression after initiating or stopping treatment on estimates of infections and deaths averted by scaling up antiretroviral therapy. PLoS One. 2018;13(3):e0194220.

30. Palella Jr FJ, Deloria-Knoll M, Chmiel JS, et al. Survival benefit of initiating antiretroviral therapy in HIV-infected persons in different CD4+ cell strata. Annals of internal medicine. 2003;138:620–626.

31. Avenir Health. Spectrum system of policy models. 2019; https://www.avenirhealth.org/software-spectrum.php. Accessed 2019-10-25.

32. Wilson DP, Law MG, Grulich AE, Cooper DA, Kaldor JM. Relation between HIV viral load and infectiousness: a model-based analysis. The Lancet. 2008;372(9635):314–320.

33. Rodger AJ, Cambiano V, Bruun T, et al. Sexual activity without condoms and risk of HIV transmission in serodifferent couples when the HIV-positive partner is using suppressive antiretroviral therapy. JAMA. 2016;316(2):171–181.

34. Cohen MS, Chen YQ, McCauley M, et al. Antiretroviral therapy for the prevention of HIV-1 transmission. N Engl J Med. 2016;375(9):830–839.

35. Linthwaite B, Sangare N, Trottier H, et al. In-care HIV cascades for the city of Montreal: Data from the Cohorte Montréalaise Executive Summary - November 29, 2018. 2018.

36. Johnson WD, O’Leary A, Flores SA. Per-partner condom effectiveness against HIV for men who have sex with men. AIDS. 2018;32(11):1499–1505.

37. Centers for Disease Control and Prevention. Updated guidelines for antiretroviral postexposure prophylaxis after sexual, injection drug use, or other nonoccupational exposure to HIV— United States, 2016. 2016.

38. Ministère de la Santé et des Services sociaux. La prophylaxie préexposition au virus de l’immunodéficience humaine : Guide pour les professionnels de la santé du Québec. 2019.

39. Molina JM, Capitant C, Spire B, et al. On-demand preexposure prophylaxis in men at high risk for HIV-1 infection. N Engl J Med. 2015;373(23):2237–2246.

40. Institut national de santé publique du Québec. INSPQ Public health expertise and reference centre. https://www.inspq.qc.ca/en/institute/about-us. Accessed 08-12, 2019.

41. R: A language and environment for statistical computing [computer program]. R Foundation for Statistical Computing,; 2019.

42. Eddelbuettel D, Francois R. Rcpp: Seamless R and C++ integration. Journal of Statistical Software. 2011;40(8):1–18.

43. Joint United Nations Programme on HIV/AIDS (UNAIDS). Cities on the road to success: Good practices in the Fast-Track cities initiative to end AIDS. 2019.

44. Cahill S, Valadez R. Growing older with HIV/AIDS: new public health challenges. Am J Public Health. 2013;103(3):e7–e15.

45. Public Health Agency of Canada. Estimates of HIV incidence, prevalence and Canada’s progress on meeting the 90-90-90 HIV targets, 2018. 2020.

46. Hertog S. Heterosexual behavior patterns and the spread of HIV/AIDS: the interacting effects of rate of partner change and sexual mixing. Sex Transm Dis. 2007;34(10):820–828.

47. Hollingsworth TD, Anderson RM, Fraser C. HIV-1 transmission, by stage of infection. J Infect Dis. 2008;198(5):687–693.

48. Brenner BG, Ibanescu RI, Hardy I, et al. Large cluster outbreaks sustain the HIV epidemic among MSM in Quebec. AIDS. 2017;31(5):707–717.

49. Dawson L, Benbow N, Fletcher FE, et al. Addressing ethical challenges in US-based HIV phylogenetic research. J Infect Dis. 2020;222(12):1997–2006.

50. Andersson E, Shao W, Bontell I, et al. Evaluation of sequence ambiguities of the HIV-1 pol gene as a method to identify recent HIV-1 infection in transmitted drug resistance surveys. Infect Genet Evol. 2013;18:125–131.

51. Grace D, Nath R, Parry R, Connell J, Wong J, Grennan T. ‘… if U equals U what does the second U mean?’: sexual minority men’s accounts of HIV undetectability and untransmittable scepticism. Cult Health Sex. 2020:1–17.

52. Millett GA, Peterson JL, Flores SA, et al. Comparisons of disparities and risks of HIV infection in black and other men who have sex with men in Canada, UK, and USA: a meta-analysis. The Lancet. 2012;380(9839):341–348.

