## Supplemental document for "Past dynamics of HIV transmission among men who have sex with men in Montréal, Canada: a mathematical modelling study"

**Modelling HIV transmission among Montreal gbMSM**

**Description of the individual-based model**

### Table of figures

### Table of Tables

**Table 1:** Summary of Montreal population level surveys used to inform the model structure and parametrization.	10

**Table 2**: Montreal’s total male population size and growth rate between 1986 and 2018.	12

**Table 3:** Male population size (15+ years) in Montreal stratified by age in 1986.	13

**Table 4:** Serosorting mixing matrix (S).	20

**Table 5:** Proportion of the population whose sexual position preference was insertive, receptive, or versatile..	20

**Table 6:** AIDS related death rates per person-year for antiretroviral therapy (ART) naïve people living with HIV, stratified by age and CD4 count.	22

**Table 7:** Death rates per person-year for people living with HIV who are on antiretroviral therapy (ART), stratified by age, CD4 count, and time on ART.	23

**Table 8:** Window period between HIV infection and the ability to detect HIV	24

**Table 9:** The proportion of men who reported ever having received an HIV test	24

**Table 10:** The proportion of men who reported having been tested for HIV within the previous year.	25

**Table 11:** HIV testing rates per year stratified by year and sexual partnering level.	25

**Table 12:** Per partnership probability of condom use stratified by time, type of relationship, serostatus, and sexual partnering level.	30

**Table 13:** Per act probability of condom use for men in a regular partnership for the respective proportion of sex acts protected with condoms	31

**Table 14:** Model parameters and their prior distributions, if applicable.	34

**Table 15:** Table of calibration outcomes and cross validation outcomes	39

**Table 16:** Approximate Bayesian Computation Sequential Monte Carlo calibration settings and parameters.	41

### 1 Context

The disease that is now known as *Acquired Immunodeficiency Syndrome* (AIDS) was first reported in Canada in 1982 ^1^. The *Human Immunodeficiency Virus* (HIV) is the necessary cause for AIDS. HIV reporting began in Canada in 1985 and the number of reported HIV cases has increased such that each year there are approximately 2,000-2,700 new diagnoses ^2,3^. In 2016, the national prevalence of HIV was 173 per 100,000 people^4^. Geographically, Québec, Ontario, and British Columbia account for 86% of people living with HIV (PLHIV) in Canada ^5^. Approximately 30% of the country’s PLHIV reside in Québec ^6^. More than half of new diagnoses in Québec occur among gay, bisexual and other men who have sex with men (gbMSM), and this group represents 71% of Québec’s male PLHIV population ^7^. This overrepresentation is thought to result, among other factors, from the increased risk of HIV transmission through anal intercourse: risk of HIV acquisition is 18 times greater during receptive anal intercourse as compared to receptive vaginal intercourse ^8^. Recent trends in new diagnoses suggest that the number of new diagnoses among gbMSM has been persistently stable over the last 15 years. Additionally, gbMSM continue to represent the highest number of new HIV diagnoses ^9^. For these reasons gbMSM remain a priority key population in the fight against HIV/AIDS.

While there is currently no cure for HIV, prevention strategies have been an integral component of programs that aim to curb HIV incidence. The majority of early prevention strategies focused on behavioural interventions, however, biomedical interventions are increasingly being used to decrease both acquisition and transmission risks ^10^. Behavioural strategies include, but are not limited to: condom use, serosorting and sero-positioning, and reducing an individual’s number of concurrent partners ^11^. Similarly, biomedical interventions include: antiretroviral therapy (ART) for PLHIV and pre- and post-exposure prophylaxis (PrEP and PEP) for uninfected individuals ^10^. Despite this multitude of potential interventions, no single strategy is sufficient on its own to control the HIV/AIDS epidemic. Rather, it has been suggested that combining HIV interventions into packages presents the optimal method to control HIV transmission at the population-level ^12^.

This technical document is organized as follows. First, a general overview of the model is provided. Second, the model structure is described in detail, including the demographic processes (i.e., aging, immigration into the model, and natural death), sexual behaviours (i.e., partnership formation and dissolution), HIV transmission and progression, and intervention implementation (i.e., condom use, HIV testing, PrEP, PEP, and ART). Finally, the model calibration process, targets, and results are presented.

### 2 Model overview

An individual-based modelling framework was chosen for its flexibility and ability to comprehensively model heterogeneity and complex interventions. The model is coded in R version 3.6.0 ^13^ using a C++ back-end through the *Rcpp* package ^14^. The main characteristics of our agent-based model (ABM) are described below.

*Main characteristics*

The ABM simulates demographic processes, partnership formation and dissolution, HIV transmission and acquisition, natural disease progression, and the impact of HIV interventions. The model was simulated among an initial population of 10,000 gbMSM in Montréal over the 1975-2019 period in 2-week increments. Men enter the model at sexual debut (15 years of age) and leave the population as a result of natural or HIV-related death. Each of these processes incorporates chance events, resulting in a stochastic representation of population and transmission dynamics.

In addition to simulating the natural history of HIV/AIDS (Figure 1), the population-level demographic processes, and the gbMSM sexual network, the model mirrors the evolving HIV prevention landscape over time. Specific interventions include HIV testing and treatment, condom use, PrEP, and PEP. HIV treatment became available in Québec in 1996 ^5,15^. The treatment eligibility criteria have changed over time from PLHIV whose CD4 cell count was less than 350 cells $\mu L^{-1}$(1996–2013) ^16^, to PLHIV with a CD4 cell count <500 cells $\mu L^{-1}$ (2013-2016) ^17,18^, to all PLHIV from 2016 onwards ^19^. With optimal adherence (>95%) ^20^, HIV treatment has been shown to inhibit disease progression and transmission ^21^. PEP, has been prescribed in Québec since the early 2000s ^22^ and can be used by HIV negative men following unprotected sexual contact with HIV positive or status unknown men. PEP has been shown to reduce HIV acquisition by approximately 81% ^23^. Finally, although available prior to 2013, provincial level guidelines and funding for PrEP were introduced in Québec in 2013 ^24^. PrEP is used for men who engage in condomless anal sex with multiple partners, and has an efficacy of approximately 86% in preventing the acquisition of HIV, and possibly close to 100% among those fully adherent ^25^. In contrast to PEP, PrEP is used prior to sexual activity.

*Population level surveys*

The model was parameterized with data obtained from Montréal-based studies, the peer-reviewed literature, and the grey literature. The following section summarizes key points regarding the four main surveys used to parameterize the model. Further information about the study design for each survey can be found in Table 1.

The model incorporates standardized data from the *Argus* *I* (2005) and *Argus II* (2008) studies which were part of the *Public Health Agency of Canada*’s M-Track bio-behavioral surveillance system ^26,27^. The eligibility criteria included gbMSM who were 18 years or older, who lived on the Island of Montréal, and had had sex with another man during their lifetime. A detailed description of the study and study results can be found in ^26,27^. As these surveys recruited participants using a time-location convenience sampling scheme, they were standardized by age and ethnicity, to the baseline data from the *Engage* cohort (2017-19). The *Engage* cohort is an ongoing, multi-city study ^28^ conducted in Montréal by the *Direction de Santé Publique de Montréal*, in collaboration with representatives from community organizations and universities. Study eligibility included gbMSM 18 years of age and older who had had sex with another man within the past 6 months. The study uses a respondent-driven sampling methodology that can result in a more representative sample population and was analyzed using Volz-Heckathorn weights. Inverse probability weights were used to account for loss to follow up in the longitudinal analysis. A detailed description of the study methods and results can be found in ^28,29^. Lastly, clinical data from the *l’Actuel PrEP Cohort* (2013-2019) a prospective open cohort of patients consulting for and prescribed PrEP at Montréal’s Clinique l’Actuel was analyzed and standardized by age to the *Engage* cohort. A description of the *L’Actuel PrEP* cohort can be found in (73). The studies were analyzed to identify time varying trends in sexual behaviours among gbMSM in Montreal.

**Table 1:** Summary of Montréal population level surveys used to inform the model structure and parametrization.

| Survey |  | Recruitment year | Sample size | Study design | Sampling method | Ref |
| --- | --- | --- | --- | --- | --- | --- |
| *Argus (M-Track)* | Argus I | 2005-2007 | 1957 | Cross- sectional | Venue based convenience sampling | ^26,27^ |
|  | Argus II | 2008-2010 | 1873 |  |  |  |
| *Engage* |  | 2017-2018 | 1179 | Cohort | Respondent driven sampling | ^28,29^ |
| *L’Actuel* |  | 2013-2019 | 2746 (restricted to gbMSM) | Cohort | Clinic-based convenience sampling | ^30^ |

*Calibration*

Approximate Bayesian Computation-Sequential Monte Carlo sampling (ABC-SMC) model fitting techniques were used to estimate parameters and replicate the HIV epidemic trajectory. Prior distributions for these model parameters were elicited by analyzing the *Argus surveys (2005, 2008)* and the *Engage* cohort (2017-2019), as well as using data from the *Institut national de santé publique du Québec* (INSPQ) (2002-2017). Fitting outcomes included both partnership dynamic outcomes (e.g., number of annual sexual partners and the duration of regular partnerships), as well as disease dynamics outcomes (e.g., HIV prevalence and CD4 cell count at diagnosis).

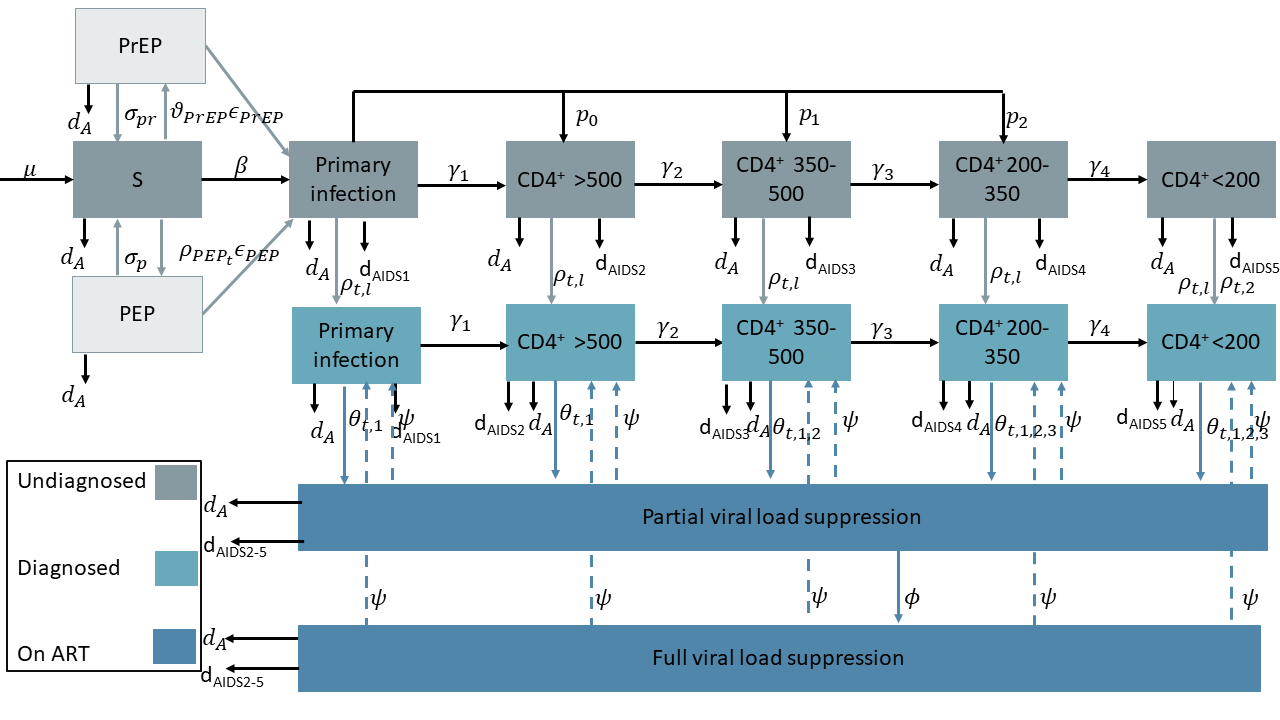

**Figure 1**: HIV natural history and HIV intervention model structure. Susceptible men (S) can acquire HIV with a probability $\beta$, through sexual contact with PLHIV. At the end of the primary stage of infection, a proportion ($p_{i_{i=0..2}})$ of men will transfer directly to the mid stages of infection. All other men can transition through each disease state at a state specific rate (${\gamma_{i}}_{i=1..4})$. Susceptible men who engage in sexual practices that carry a substantial risk of HIV transmission (i.e. unprotected sex with a casual partner) can take pre-exposure prophylaxis (PrEP), at a time-varying rate of $\vartheta_{{PrEP}_{t}}$, or post-exposure prophylaxis (PEP) at a time-varying rate of $\rho_{PEP_{t}.}$ Given the efficacy ($\epsilon_{PEP}$) of PEP,$\rho_{PEP_{t}}\epsilon_{PEP}$ men will be protected from potential HIV infection. All men can get tested for HIV, which occurs at a rate of $\rho_{t,l}$and$\rho_{t,2}$ for symptomatic diagnoses. The testing rate was stratified by sexual partnering level, $l$. The test sensitivity and specificity are assumed to be 100%. Once tested positive, men can initiate antiretroviral therapy (ART). Men initiate treatment at a time-varying rate, $\theta_{t,1,2}$. Since the ART guidelines have changed over time with respect to CD4 cell count eligibility, the eligibility is indexed by the respective guidelines with 3 =1996 – 2013, 2 = 2013-2016, and 1= 2016 onwards. During the first $\phi$ months of ART use, men are categorized as short-term ART users (*sART*), after which they are categorized as long-term ART users (*lART*). Neither *sART* nor *lART* progress through the natural disease history. Men can discontinue ART use at a rate $\psi$ in which case they regress to the disease stage at which they initiated ART. Finally, at each stage of the model, men can die from natural causes at a death rate $d_{A}$, or from AIDS related causes, the rate of which varies at each disease stage and ART status.

### 3 Model structure

#### 3.1 Demography

##### 3.1.1 Population size

INSPQ’s survey *Enquête québecoise sur la santé des populations* (EQSP) conducted in 2014-2015 estimated that 5.6% (4.5-6.8%) of sexually active men aged 15 and older residing in Montréal reported having had at least one sexual partner of the same sex in the last 12 months. Additionally, the survey estimated that 0.8% (0.5-1.4%) of men had sexual contact with both men and women within the preceding 12 month period ^31^. For the purposes of this model, it is assumed that the proportion of men in Montréal who are gbMSM has remained constant over time at 6.4%.

*Census Canada* and the *Institut de la statistique du Québec* reports for 1976, 1981, 1986, 1991, and 1996-2018 indicate an overall decrease in the Island of Montréal’s population between 1975 and 1986 of approximately 10%, and an overall increase thereafter (Table 2) ^32-36^. The time varying growth rate at which individuals enter the population was calculated for 5-year periods assuming an exponential growth rate. Given the small decrease in the population size and the uncertainty regarding the characteristics of people that emigrated from Montréal between 1975 and 1986, the model assumed a constant population size during this time period. Consequently, the initial model population size was calculated as the population size circa 1986. An examination of the population age distribution resulted in similar trends over time. Therefore, the proportion of men in each age group was estimated from the 1986 Québec census ^32^ and was used to calculate the respective number of men who are gbMSM (Table 3).

**Table 2**: Montréal’s total male population size and growth rate between 1986 and 2018. The growth rate was calculated assuming an exponential growth rate within each 5-year period.

| **Year** | **Male population size** | **Growth rate (per year) between period** | **Reference** |
| --- | --- | --- | --- |
| 1986 | 832,150 | NA | ^32^ |
| 1991 | 846,305 | 0.003 | ^33^ |
| 1996 | 863,685 | 0.004 | ^36^ |
| 2001 | 895,487 | 0.007 | ^36^ |
| 2006 | 915,017 | 0.004 | ^36^ |
| 2011 | 938,554 | 0.005 | ^36^ |
| 2016 | 965,804 | 0.006 | ^36^ |
| 2018 | 1,002,391 | 0.019 | ^36^ |

**Table 3:** Male population size (15+ years) in Montréal stratified by age in 1986. The proportion of the population who identified as gay, bisexual and other men who have sex with men (gbMSM) was assumed to remain constant (6.4%) over time (33, 34).

| Age group (years) | Proportion of men in each age group | Male population size in 1986 | Estimated number of gbMSM in 1986 |
| --- | --- | --- | --- |
| 15-19 | 8.5% | 58,765 | 3,761 |
| 20-24 | 12.3% | 85,020 | 5,441 |
| 25-34 | 23.5% | 162,270 | 10,385 |
| 35-44 | 16.9% | 116,945 | 7,484 |
| 45-54 | 13.6% | 94,170 | 6,027 |
| 55-64 | 13.0% | 89,975 | 5,758 |
| 65-74 | 8.0% | 55,015 | 3,521 |
| > 74 | 4.2% | 28,840 | 1,846 |
| Total | 100.0% | 691,000 | 44,223 |

##### 3.1.2 Entry into the model, aging, and natural death

The demographic processes included in the model (i.e., entry into the model, natural death, and aging) are modelled using an annual time step. All men who entered the model population after the model initialization did so at 15 years of age (sexual debut) and were assumed to be susceptible to HIV infection. As men aged, their chances of death due to natural causes increased. The probability associated with dying from natural causes, $p_{death\_age}$ was derived from Québec-specific male life expectancies (Figure 2) ^37^. For simplicity, we assumed that the maximum age a man could live to was 100 years old. A binomial distribution was used to determine which men died each year, such that $p_{death(age)}=\frac{1}{life expectancy(age)}$. In addition to natural death, men could exit the model due to HIV-related death. This process is described in Section 3.4.

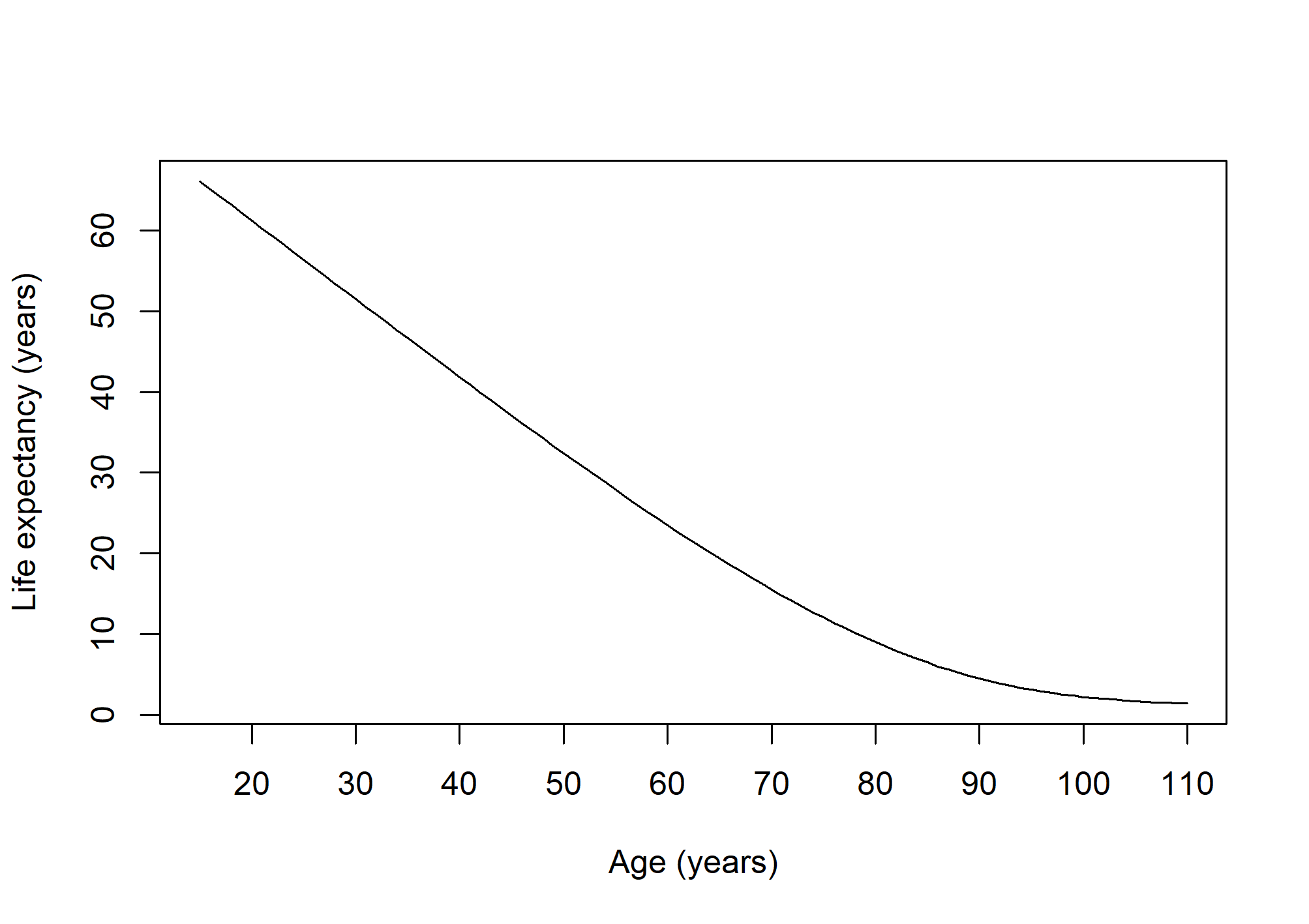

**Figure 2:** Life expectancy (years) for Québec males aged 15+ years old. The life expectancy was assumed to be 0 for men aged 100 years and older (50).

##### 3.1.3 Demographic stratification

###### 3.1.3.1 Transgendered individuals

Although the term gbMSM encompasses a broad range of sexual orientations (e.g., gay, bi-sexual) and gender identities (e.g., transgender, two-spirit) ^38^, this study focuses on cis-gender gbMSM. There is a striking paucity of data on transgender women in Montréal and Canada more generally. As such, we cannot reliably model HIV transmission among transgender women. However, we believe that this should not affect our inferences since several recent phylogenetic studies suggest that HIV infections among transgender women often belong to different phylogenetic clusters than that of gbMSM ^39^. Additionally, HIV prevalence among trans men and trans gbMSM has been shown to be low (1.4 – 3.2%) ^40,41^ and as such, trans men were not included in the model.

###### 3.1.3.2 Ethnicity

A study by Millett et al., (2012) suggested minimal racial disparities in HIV prevalence among gbMSM in Canada ^42^. This result was mirrored in the standardized *Argus* and RDS-adjusted *Engage* cohort in which there were no major differences in HIV prevalence within key ethnicities (Figure 3). Furthermore, when differences were observed, the temporal trends in prevalence were inconsistent and the uncertainty intervals of the different estimates overlapped widely. As such, there is currently no compelling evidence of diverging trends among ethnicities that would need to be incorporated in the model to reflect the available survey data.

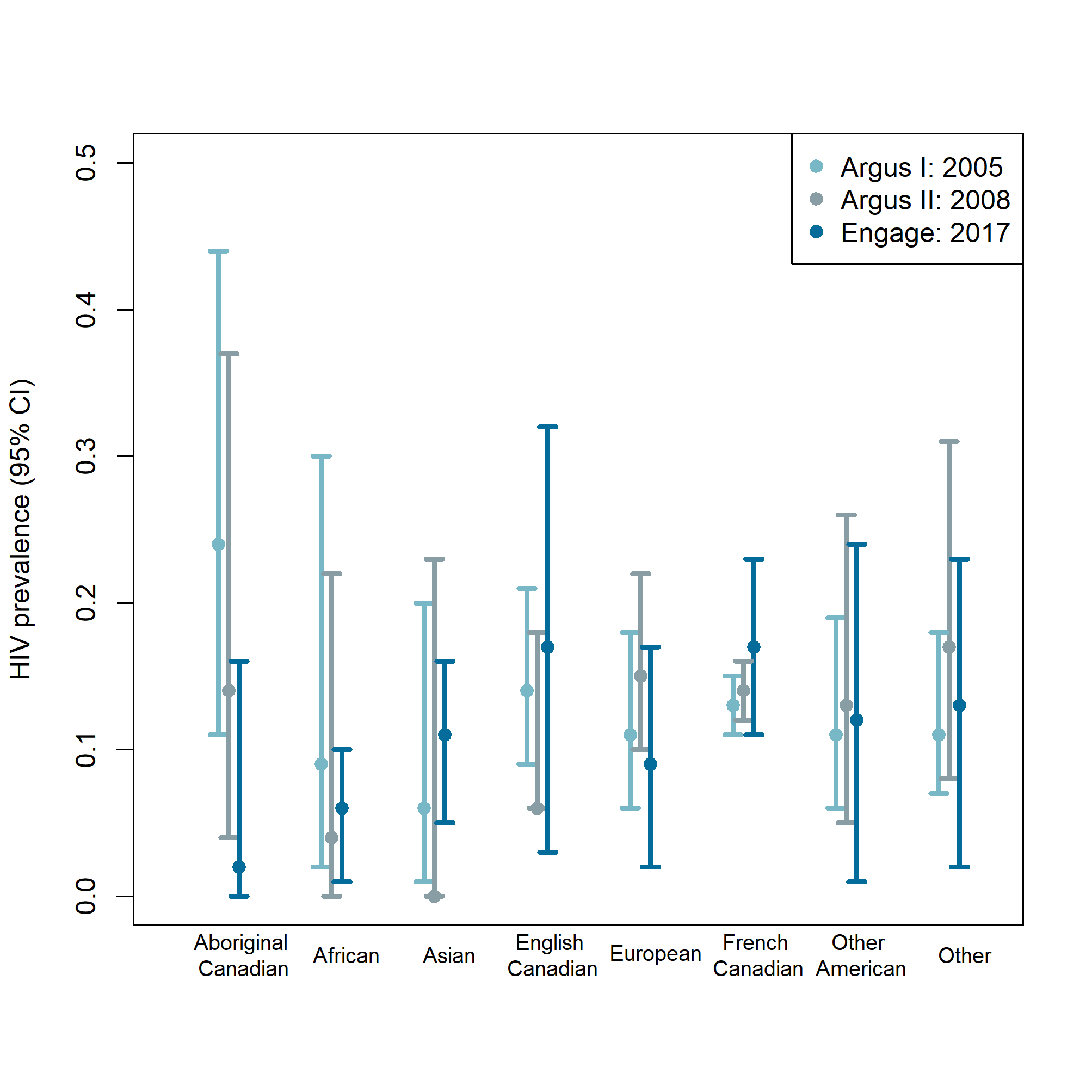

**Figure 3:** HIV prevalence by ethnicity. The prevalence was obtained from the standardized Argus I ^26^, Argus II ^26^ and RDS-adjusted Engage ^28^ population surveys. The HIV prevalence ranged between 6% (Asian) and 24% (Aboriginal Canadian) in 2005, 0% (Asian) and 17% (other) in 2008, and 2% (Aboriginal Canadian) and 17% (English and French Canadian) in 2017.

#### 3.2 Sexual behaviors

##### 3.2.1 Partner change rate

A key contributor to HIV acquisition risk is an individual’s partnership change rate ^43,44^. Using data from two cross-sectional surveys (*Argus I, Argus II*) and baseline results from one cohort (*Engage*), we did not detect important changes regarding the average reported number of anal sex partners between 2005 and 2017 (Figure 4). Therefore, the partnership change rate was calculated from the combined *Argus I*, *Argus II*, and *Engage* cohort. Specifically, the partnership change rate was inferred from the question:

- Argus I, Argus II, and Engage: “*During the past 6 months, how many guys have you had anal sex with*?”.

A negative binomial distribution was fit to the data and used to assign to each man, upon entry into the model, an average number of anal sex partners over a 6-month period (Eq. 1). This number was scaled to represent each man’s average number of annual anal sex partners. Since the partner distribution varied as a function of age, multipliers were estimated and used to scale the mean of the negative binomial distribution for 5-year age bands (Figure 5). The multipliers ($RR\_age$) were calculated using meta regression fitted to the log-transformed age-specific self-reported number of anal sex partners. As individuals age, their partnership change rate is scaled accordingly.

$Average annual number partners(age) \sim NB(RR\_age*mean\_partners, size\_partners)$	(1)

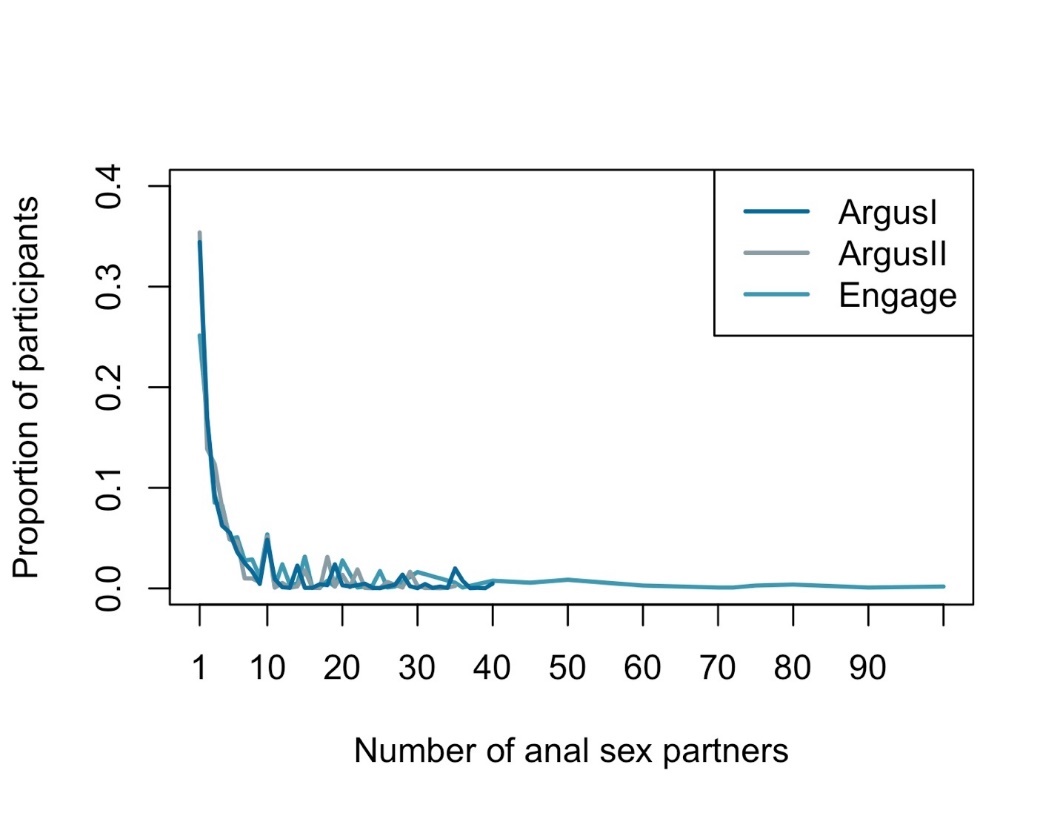
**Figure 4:** Time varying distribution of the number of anal sex partners that each participant reported having within the 6 months prior to completing the respective survey. The data was obtained from the standardized Argus I, Argus II, and baseline data from the respondent-driven sampling adjusted Engage cohort. There were no important temporal differences in the number of reported anal sex partners.

In addition to an annual partner change rate, the maximum number of partnerships (both concurrent and serial) that each man could have at each time step was assigned from a Poisson distribution (Eq. 2).

$Number\_partners\sim Poisson(\frac{Ave\_annual \_num\_partners(age)}{dt})$	(2)

Following other models, we categorized men into “sexual partnering” groups based on their annual number of anal sex partners. Specifically, we defined the “*low partnering*” gbMSM as men who have 5 anal sex partners or less per year, “*medium partnering*” as those with 6-10 anal sex partners per year, and “*high partnering*” as those who have 11 or more anal sex partners per year ^45^.

| A)  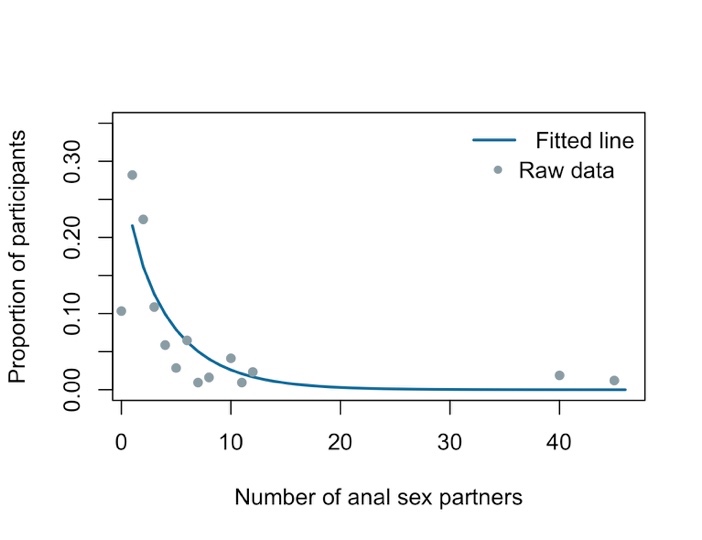 | B)  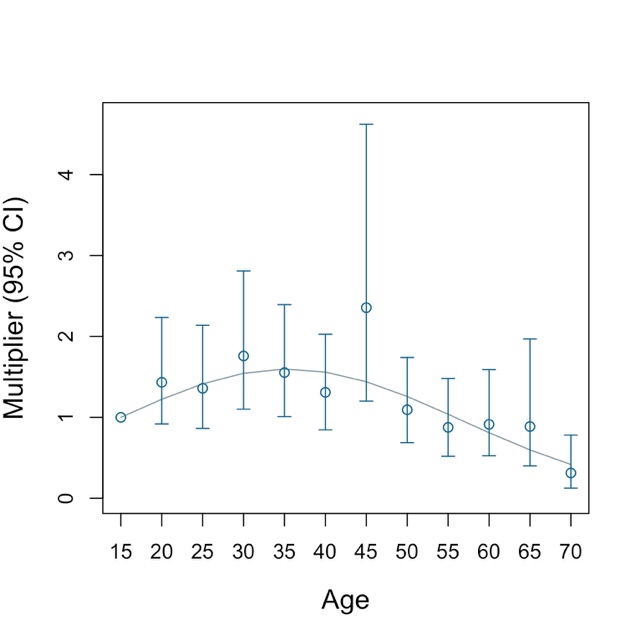 |
| --- | --- |
| **Figure 5:** Number of anal sex partners that each participant reported having within the 6 months prior to completing the respective survey. The data was obtained by aggregating the standardized *Argus I* and *Argus II* surveys, and baseline data from the respondent-driven sampling adjusted *Engage* cohort. A) Fitted negative binomial distribution and raw data of number of anal sex partners in the referent group (men aged 15-19 years old). B) Meta regression line and calculated multiplier with 95% confidence intervals for each age group. | |

##### 3.2.2 Partnership duration

The model considers two types of partnerships: casual and regular. Casual partnerships are assumed to be instantaneous (i.e., they begin and end within the 2-week time step), while regular partnerships are assumed to have a minimum duration of 1 month. Each partnership is assigned a duration upon formation. Since it was assumed that each man can have only one regular partner at a time, if either partner is already in a regular partnership, any new sexual partner will automatically be of casual status. If, however, neither partner is in a relationship with a regular partner, a Bernoulli distribution is used to determine whether the partnership will be assigned a status as casual or regular (Eq. 3).

$partnership\_type \sim Bernoulli( p_{main})$	(3)

The probability informing this distribution was obtained from the Argus I, Argus II, and Engage surveys. A negative binomial distribution is used to assign a duration to regular partnerships (Eq. 4). The parameters of this distribution were obtained using maximum likelihood estimates fit to the censored data from the combined *Argus I*, *Argus II*, and Engage (Figure 6).

$duration\_main \sim NB(\mu_{duration},s_{duration})$	(4)

Specifically, the following questions were used:

- Argus I and Argus II surveys: “*since what date have you been part of a couple with this man?*” and “*for how long have you been part of a couple with this man*?”
- Engage survey: “*how long have you been with this [*regular*] partner*?”.

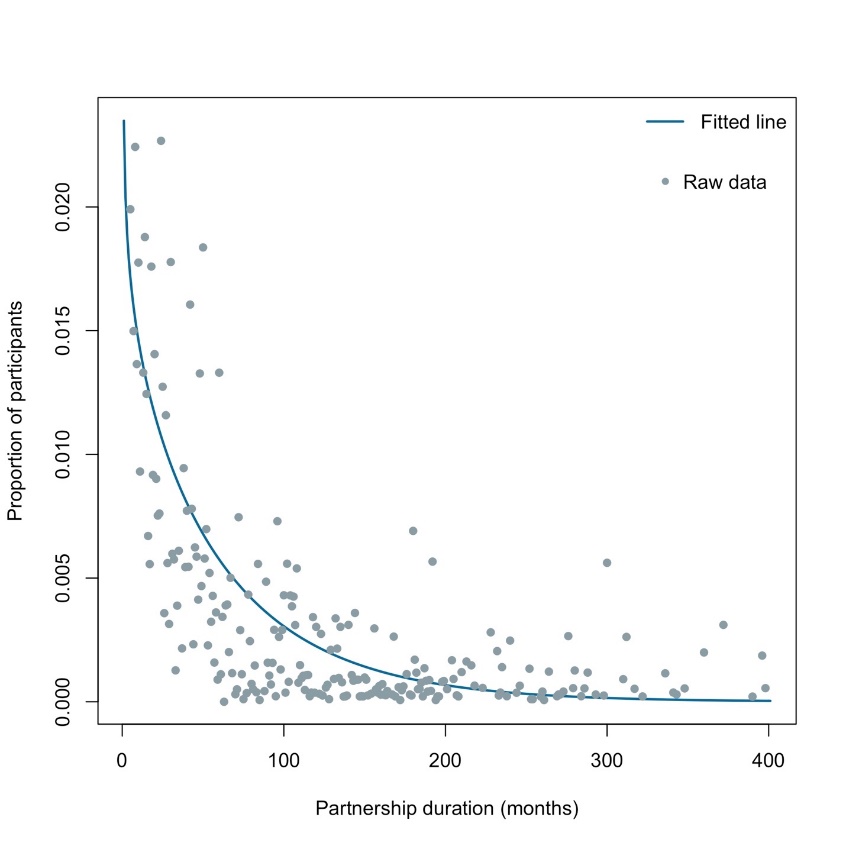

**Figure 6:** Duration of participants’ ongoing regular partnership (minimum of 1 month) at the time of the survey. The data was obtained from the pooled standardized Argus I and Argus II surveys and baseline data from the respondent-driven sampling adjusted Engage cohort and was right censored. Approximately 18% of participants had been in a relationship for more than 8 years.

##### 3.2.3 Mixing

Three mixing matrices were used in the partner formation process: an age preference matrix (A), a serosorting matrix (S), and a sexual role mixing matrix (R). The age mixing matrix was calculated using data from the Argus I survey which asked participants to indicate the approximate age of the last man that they had sex with. The data was aggregated into 5-year age groups in which the proportion of men in each age group whose partner fell into the corresponding age group was calculated (Figure 7).

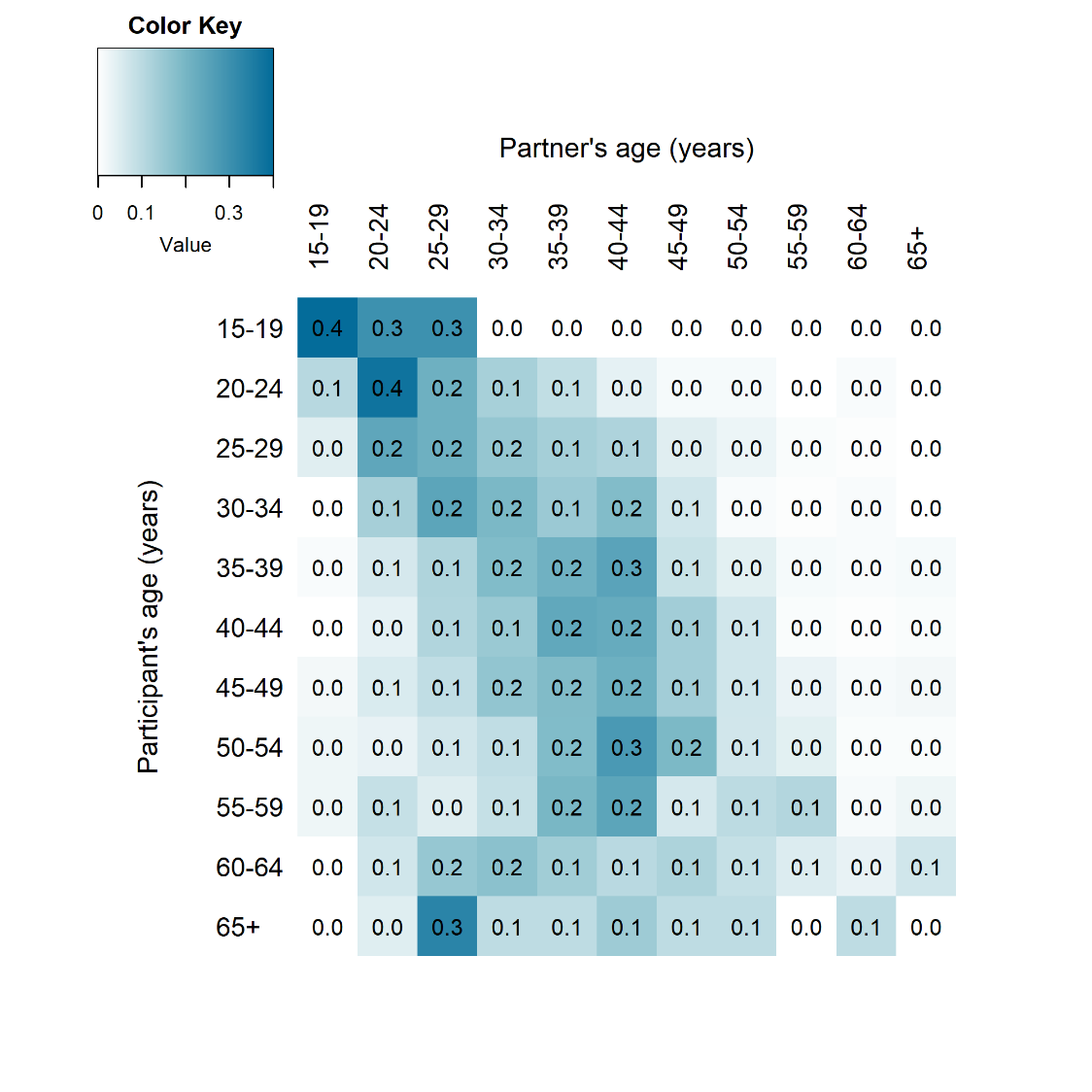

**Figure 7:** Age mixing matrix (A) representing the proportion of men in each age category who indicated that the age of their last sexual partner fell within the corresponding age category. The majority of men indicated that their sexual partners fell within a similar age group to themselves, as is indicated by the darker band down the diagonal of the heatmap. The data was obtained from the *Argus I* survey.

The serosorting matrix (S) describes the proportion of sero-concordant and sero-discordant partnerships experienced by participants in the combined *Argus II* survey and Engage cohort (Table 4). Specifically, the two following questions were used to construct the mixing matrix:

- Argus II: “*during the past 6 months, have you [fucked / been fucked] by a [regular/ occasional/ one-night stand] partner who was [infected by HIV/ not infected by HIV/ of unknown HIV status]”*.
- Engage: “*the most recent time you had sex with the partner named above, did you know what his HIV status was before you guys had sex?”*.

Since the Engage cohort collected data on the participants’ 5 most recent sexual partners’ serostatus, each partner’s serostatus was taken into consideration in the mixing matrix. The *Engage* data was combined with the *Argus II* data to calculate the serosorting mixing matrix. Men with an unknown HIV status were assumed to be HIV negative and all known HIV positive men, regardless of their viral load were considered to be HIV positive.

**Table 4:** Serosorting mixing matrix (S). The mixing matrix represents the proportion of partnerships among men of a given sero-status with HIV negative and HIV positive partners. An HIV negative designation represents men with a negative test result, or men that do not know their HIV status. The data was calculated from the standardized *Argus II* ^26^ survey and baseline data from the respondent-driven sampling adjusted *Engage* cohort ^28^.

|  | Partner’s HIV status | | |
| --- | --- | --- | --- |
| Participant’s HIV status |  | **HIV- or unknown HIV status** | **HIV+** |
|  | **HIV- or unknown HIV status** | 0.924 | 0.076 |
|  | **HIV+** | 0.660 | 0.340 |

Upon entry into the model, each man is assigned a life-long sexual role (i.e. each man is strictly insertive, receptive, or versatile). The proportion of men in each preference category was obtained from the Engage survey which asked participants:

- “*on a scale of 1 (always get fucked) to 5 (always fuck the other guy), what kind of anal sex do you prefer?*”.

For the purposes of this model, men who responded *“always get fucked”* were assumed to be strictly receptive, men who answered “*always fuck the other guy*” were assumed to be strictly insertive, and all other responses were assumed to indicate versatility (Table 5). If a man’s assigned role is insertive or receptive, and he is partnered with a man whose role is receptive or insertive respectively, then each man retains their status for the duration of the relationship. If a versatile man forms a partnership with an insertive man or a receptive man, then the versatile man takes on a receptive or insertive role respectively for the duration of the partnership. Finally, if two versatile men form a partnership, then one man is assigned a status of insertive or receptive at random, and the second man is assigned the opposing sexual role. In a partnership of two versatile men, the men can change their sexual roles at each time step.

**Table 5:** Proportion of the population whose sexual position preference was insertive, receptive, or versatile. The data was obtained using baseline data from the respondent-driven sampling adjusted *Engage* cohort ^28^.

|  | Proportion | 95% CI |
| --- | --- | --- |
| Insertive | 19.6% | (14.8-24.4%) |
| Receptive | 16.9% | (12.9-21.0%) |
| Versatile | 63.5% | (58.0-69.0%) |

A multinomial distribution implemented using a uniform distribution was used to determine each man’s preferred partner age, serostatus, and sexual role (Eq. 5). If no eligible partners exist given the selected age, serostatus and sexual role, then the search criteria are reduced to include sexual role and age. Similarly, if no partner is found using the updated criteria, then the preferred serostatus and role are used to find an eligible partner. If the man does not find a partner using any of the above criteria, then he is not assigned a partner for the duration of the time step.

$Partner\_characteristics \sim Mult(A,S,R)$	(5)

##### 3.2.4 Number of anal sexual acts per partnership

Every man is allotted a specific number of anal sex acts for each assigned partner for the duration of the partnership using a Poisson distribution. This number is scaled to provide an annual number of anal sex acts with each partner. The Poisson distribution was fit to the Engage cohort data which asked:

- “*How many times have you had anal sex (as top or bottom) with the partner named above in the past 6 months?*”

All casual partnerships are randomly assigned between 1-3 anal sex acts per partnership, while the assigned number of anal sex acts for two regular partners is averaged. The number of anal sex acts is reduced by 10% for PLHIV having reached the AIDS stage ^46^.

#### 3.3 HIV transmission

Although the model tracks all ongoing relationships, it only simulates sexual activity between sero-different couples, as only men in these partnerships can transmit and/or acquire HIV. The probability of HIV transmission through oral sex is negligible (0.04%) ^8,47^ and only sexual transmission through anal intercourse is modelled. External sources of infection, such as from female sexual partners, or from people who inject drugs, are not modelled.

A Bernoulli process model is used to determine the probability that the HIV susceptible men in sero-discordant partnerships acquire HIV from their partner at each time step. The probability of acquisition, $P_{t}$, is a multiplicative function based on: 1) the base acquisition probability ($p_{insertive}$) for insertive partners, 2) a multiplier $(RR_{receptive})$ for receptive partners, 3) a relative risk multiplier for the HIV positive partner’s disease stage ($RR_{CD4_{1-2}}$), and 4) the efficacy of PEP ($\epsilon_{PEP}),$ PrEP ($\epsilon_{PrEP}$), condom use ($\epsilon_{condoms}$) and ART in reducing transmission (Eq. 6). The ART efficacy is dependent on the duration of ART use, used as a proxy for viral load suppression (VLS). In this model, men were classified as short-term ART users (*sART*), as a proxy for partial viral load suppression (pVLS), or long-term ART users (*lART*), as proxy for viral load suppression. A man’s ART categorization was used to compute the ART efficacy in preventing HIV transmission such that $\epsilon_{ART\_sART}$ = 20% for *sART* and $\epsilon_{ART\_lART}$= 90-100% ^21,48^ for *lART*. The $\epsilon_{ART\_sART}$ parameter was computed assuming that VLS is defined as <200 copies/ml ^49^, and pVLS as 200-349 copies/ml ^50^. The probability of transmission for lART and *sART* was calculated using the equation: ${2.45}^{\log VL-4.5}$. Finally, the ratio comparing the probability of transmission for a *sART* to that of a *lART* was $80\%$ , resulting in $\epsilon_{ART\_sART}=20\%$. Condom use was implemented such that the per-act probability of condom use varied between 0 and 1 for regular partners and was fixed at 0 or 1 for casual partners. Therefore, number of sex acts per time step ($n$) that each couple had was multiplied by the proportion of time that a couple used a condom ($p_{c})$.

$P_{t}=1-\left( 1-{RR_{receptive}*p}_{insertive}*RR_{CD4_{1-2}}*\left( 1-\epsilon_{PEP} \right)*\left( 1-\epsilon_{PrEP} \right)*(1-\epsilon_{ART\_sART})*(1-\epsilon_{ART\_lART}) \right)^{n\left( 1-p_{c} \right)}\left( 1-{RR_{receptive}*p}_{insertive}*RR_{CD4_{1-2}}*(1-\epsilon_{PEP})*(1-\epsilon_{PrEP})*(1-\epsilon_{ART\_sART})*(1-\epsilon_{ART\_lART})*(1-\epsilon_{condoms}) \right)^{np_{c}}$	(6)

#### 3.4 HIV natural history

Once an individual acquires HIV, they experience a short highly infectious primary stage of infection which lasts for approximately 3 months ^51^. Following primary infection, men can transition to the acute, asymptomatic stage of infection which lasts for approximately 3 months ^52^. The acute stage of infection precedes the early stage of infection (CD4 cell counts >500 cells $\mu L^{-1}$ and duration of 3.3 years), and is followed by the asymptomatic mid-stages of disease, (CD4^+^ counts between 350-500$\mathrm{cells} \mu L^{-1}$; duration ($d_{350-500})=$2.7 years, and CD4 cell counts between 200-350 cells$\mu L^{-1}$; duration ($d_{200-350})=$5.5 years)), and the advanced stages of disease (CD4^+^ counts <200 cells$\mu L^{-1}$; $d_{200}=$ 5.1 years) ^53^. Additionally, 76%, 19% and 5% of men transition directly from the acute stage of infection to the each of the three mid stages of infection respectively ^53^. A Bernoulli distribution is used to determine which men transition between the relevant disease stages (Eq. 7). Men can die from HIV related causes at any point in the disease progression. In accordance with other studies ^54^, the time invariant death rate is age-dependent and varies based on time on ART (Table 6 and 7). A Bernoulli distribution is used to determine which men die from disease related causes using a stage specific probability of death (Eq. 8).

$p_{transition} \sim Bernoulli\left( p_{i_{i=0-2}}{*_{i}}_{i=1-4} \right)$(7)

$p_{death\_age\_CD4\_timeART} \sim Bernoulli(p_{death\_age\_CD4\_timeART})$	(8)

**Table 6:** AIDS related death rates per person-year for antiretroviral therapy (ART) naïve people living with HIV, stratified by age and CD4 count ^55^.

| CD4 count (cells$\boldsymbol{\mu}\boldsymbol{L}^{\boldsymbol{-1}}$) | Age 15-24 | Age 25-34 | Age 35-44 | Age 45+ |
| --- | --- | --- | --- | --- |
| > 500 | 0.005 | 0.004 | 0.005 | 0.005 |
| 350-500 | 0.011 | 0.010 | 0.013 | 0.013 |
| 200-349 | 0.044 | 0.048 | 0.066 | 0.056 |
| <200 | 0.399 | 0.675 | 0.933 | 0.670 |

**Table 7:** Death rates per person-year for people living with HIV who are on antiretroviral therapy (ART), stratified by age, CD4 count, and time on ART ^55^.

| Time on ART | CD4 count (cells$\boldsymbol{\mu}\boldsymbol{L}^{\boldsymbol{-1}}$) | Age 15-24 | Age 25-34 | Age 35-44 | Age 45+ |
| --- | --- | --- | --- | --- | --- |
| 0-6 months | **> 500** | 0.004 | 0.006 | 0.008 | 0.016 |
|  | **350-500** | 0.004 | 0.005 | 0.008 | 0.015 |
|  | **200-349** | 0.005 | 0.006 | 0.009 | 0.019 |
|  | **<200** | 0.009 | 0.011 | 0.016 | 0.033 |
| 7-12 months | **> 500** | 0.002 | 0.003 | 0.004 | 0.009 |
|  | **350-500** | 0.002 | 0.003 | 0.004 | 0.008 |
|  | **200-349** | 0.003 | 0.003 | 0.005 | 0.010 |
|  | **<200** | 0.005 | 0.006 | 0.009 | 0.017 |
| > 12 months | **> 500** | 0.001 | 0.002 | 0.003 | 0.006 |
|  | **350-500** | 0.001 | 0.002 | 0.003 | 0.005 |
|  | **200-349** | 0.002 | 0.002 | 0.003 | 0.006 |
|  | **<200** | 0.003 | 0.004 | 0.006 | 0.011 |

#### 3.5 HIV prevention and treatment

##### 3.5.1 Risk compensation

To date, studies focusing on the effect of ART ^56-62^ and PEP ^63-65^ on men’s sexual behaviour have not proven conclusive. For this reason, the model does not incorporate risk compensation behaviours.

##### 3.5.2 HIV testing

There are 4 situations in which people can get tested for HIV. For each of the scenarios, test sensitivity and specificity of 100% were assumed ^66^. Additionally, given the HIV testing window period, PLHIV will receive a negative test result if tested within a defined period after infection (Table 8).

1. **HIV negative men or men who do not know their HIV status who are neither on PEP nor PrEP**. Studies have estimated that between 1985 and 1989, and by 1995, approximately 25% ^67^ and 68-82% ^68^ of all participants reported having been tested for HIV at least once in their lifetime. Linear interpolation using these data combined with data from the *Argus I*, *Argus II*, and *Engage* surveys was used to calculate the proportion of men who reported ever having been tested (Table 9). Additionally, linear interpolation using data from the Argus I, Argus II, and Engage surveys was used to calculate the proportion of men who were tested each year (Table 10). Each year, the proportion of men who get tested for the first time is calculated by subtracting the proportion who reported ever having received an HIV test the previous year from the proportion ever tested that year. The remainder of the men to get tested that year are randomly chosen from those who have been tested previously. It was assumed that men could only be tested once prior to 1996.

A Bernoulli distribution, using the computed annual testing rate ($\rho_{test_{t}})$ and a multiplier for medium and high partnering level men ($RR_{test})$, was used to determine which of the pre-determined men were tested (Eq. 9). The testing rate was converted to a probability for use in the distribution. Additionally, a zero truncated Poisson distribution, such that $\lambda=$ $\frac{1}{\rho_{test_{t}}}$ , was used to assign men a minimum amount of time between HIV tests (Eq. 10).

$HIV\_test_{t} \sim Bernoulli(RR_{test}*\rho_{test_{t}})$	(9)

$Time\_between\_tests \sim ZTPoisson(\frac{1}{\rho_{test_{t}}})$	(10)

1. **Men on PrEP** get tested for HIV after one month on PrEP, and every three months thereafter. It was assumed that all men on PrEP are 100% compliant with the testing requirements.
2. **Men who attempt to get a PEP** **prescription** **or who have taken PEP for a period of 1 month**. All men get test prior to starting PEP. Additionally, the model assumes an 68% compliance rate with the PEP testing requirements ^69^, which require men to get tested at 1 and 3 month periods after initiating PEP ^70^.
3. **Symptomatic men at the late stage of HIV infection:** studies indicate that 6-8% of people at the late stage of infection are asymptomatic. Therefore, men who reach the late stage of infection (CD4 ^+^ cell count <200) and are symptomatic will get tested at a rate of 1 per 6 months after attaining CD4 <200.

**Table 8:** Window period between HIV infection and the ability to detect HIV ^71^

| **Year** | **1985** | **1987** | **1991** | **1997** | **2015** |
| --- | --- | --- | --- | --- | --- |
| **Detection delay after infection** | 10 weeks | 6 weeks | 3 weeks | 2 weeks | 2 weeks |

**Table 9:** The proportion of men who reported ever having received an HIV test.

| **Year** | **1989** | **1995** | **2005** | **2008** | **2017** |
| --- | --- | --- | --- | --- | --- |
| **Proportion ever tested for HIV (priors)** | 25.3% | ~U(68.0, 82.0%) | ~U(41.0, 100%)^*^ | ~U(90.4, 91.4%) | ~U(88.6, 94.1%) |
| **Reference** | ^67^ | ^68^ | Argus I | Argus II | Engage |

^*Note that these priors were inflated by 50% during the calibration process to facilitate attaining the calibration outcomes.^

| **Year** | **2005** | **2008** | **2017** |
| --- | --- | --- | --- |
| **Proportion tested within the past year** | ~U(72.5, 73.8%) | ~U(69.1, 71.1%) | ~U(68.8, 78.9%) |
| **Reference** | Argus I | Argus II | Engage |

**Table 10:** The proportion of men who reported having been tested for HIV within the previous year.

An analysis of the *Argus I* and *Argus II* survey results indicated minimal change in testing rates between 2005 and 2008. In contrast, there was a large increase in testing rates between the two *Argus* surveys and the Engage cohort (2017) (Figures 8 and 9). Therefore, the testing rates reported in the *Argus I* and *II* surveys were combined. Testing rates for each year were interpolated using linear regression which was fitted by testing rates of the following four years: 1) 1985 (the year that testing was first introduced to North America ^72^), 2) 1995, 3) 2004, and 4) 2017. Testing was not shown to vary by age but varied by sexual partnering groups. In terms of sexual partnering groups, there was minimal difference between the medium and high partnering groups, however, there was an important difference between the low partnering group and the combined medium and high partnering groups (Figure 10). To that end, the model stratifies testing rates based on men who have 1-5 partners per year, and those that have >5 partners per year. The testing rate was computed for the low partnering group (referent group), and a rate ratio multiplier was computed for the combined medium and high-partnering group (Table 11). As the low partnering group is less likely to use PEP or PrEP, the base testing rates did not exclude those who reported having ever used PEP or PrEP. The rate ratio was calculated from the combined *Argus I*, *Argus II*, and *Engage* surveys which asked participants to record the number of times that they had been tested within the past 2 years.

**Table 11:** HIV testing rates per year stratified by year and sexual partnering level.

| Sexual partnering groups | 1985 | 1995 | 2004 | 2017 | Rate ratio^1^ multiplier for the combined medium and high partnering groups |
| --- | --- | --- | --- | --- | --- |
| Low anal sexual partnering group (0-5 partners per year) | $\frac{1}{time unit}$ | $\frac{1}{time unit}$ | ~U(1.036, 1.162) | ~U(1.103, 1.471) | 1.436 |
| Reference | assumed | assumed | ArgusI, ArgusII | Engage | ArgusI, ArgusII, Engage |

**^1^** Rate ratio for calculating HIV testing rates for the high sexual partnering group

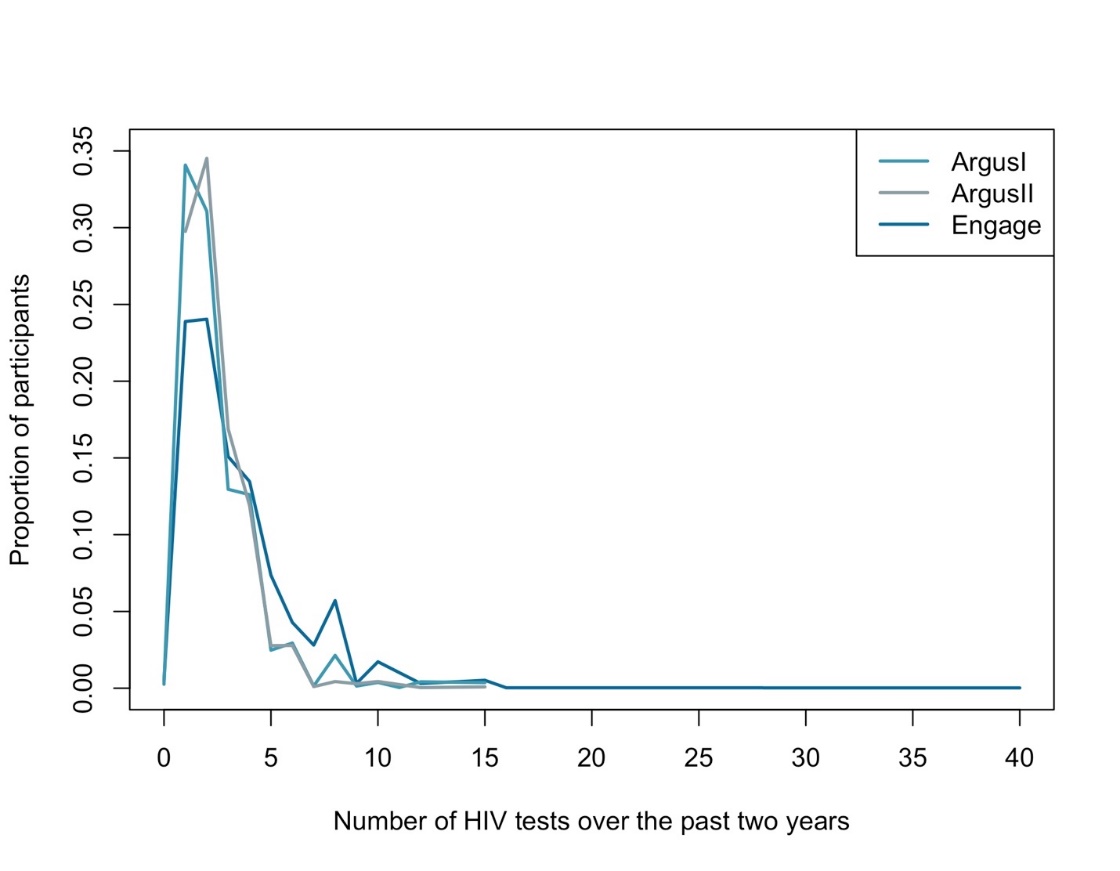

**Figure 8**: The number of HIV tests participants reported receiving over the 2-year period prior to completing the standardized *Argus I* and *Argus II* surveys, and baseline data from the respondent-driven sampling Engage cohort (weighted).

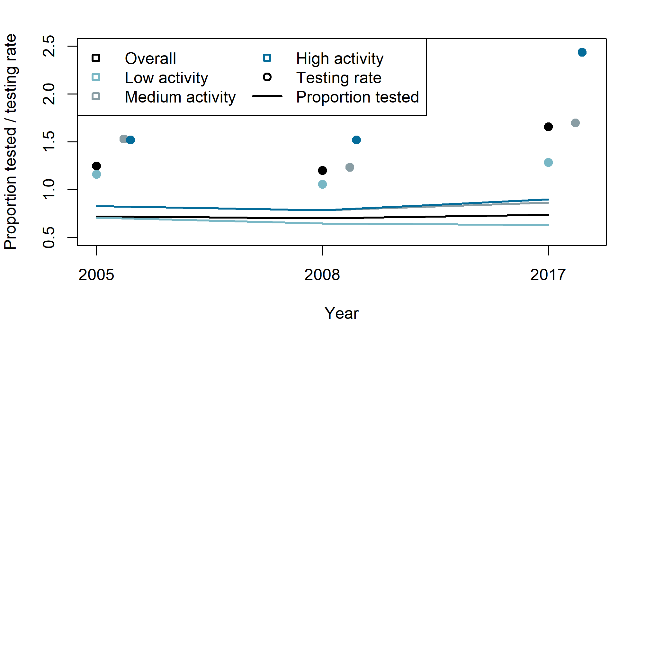

**Figure 9:** Time varying HIV testing rates and the proportion tested within the past year. Although there was no large increase in the proportion tested, the testing rate increased over time within the high partnering group.

A)

B)

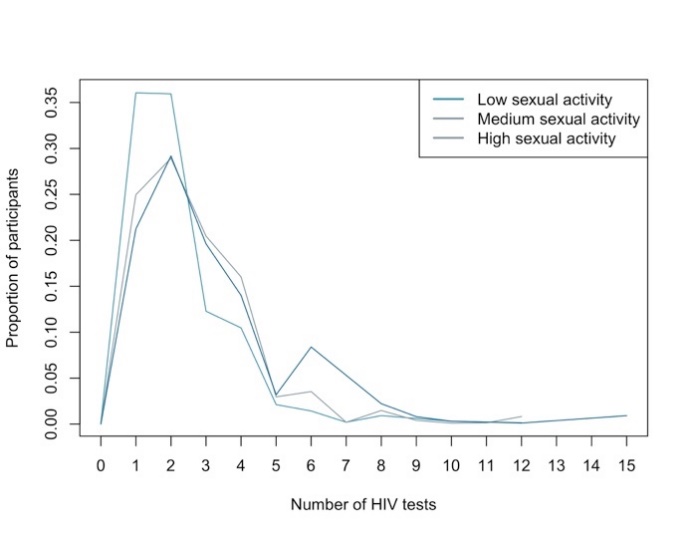

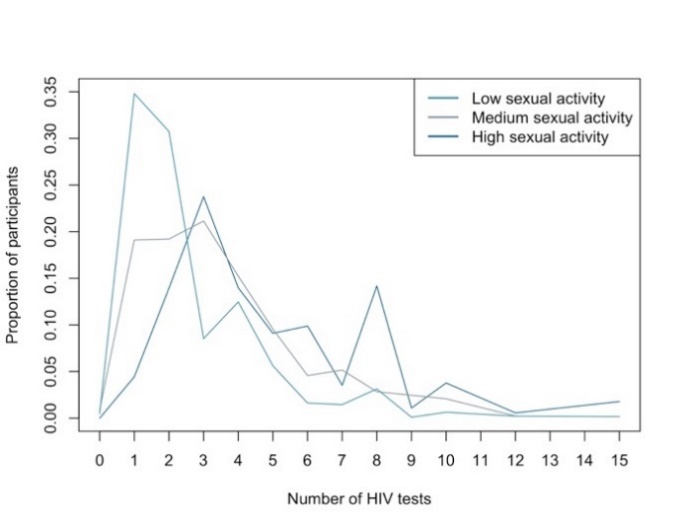

**Figure 10:** The number of HIV tests that participants in the low, medium, and high partnering levels reported over the 2 years prior to completing the respective survey. (A) Data derived from combined Argus I and Argus II survey (2005 and 2008). (B) Data derived from Engage cohort (2017).

##### 3.5.3 HIV Treatment

Changes in the ART guidelines are reflected in the model. Men who had previously tested positive for HIV but were ineligible to receive ART at the time of their diagnosis are prescribed ART as soon as they become eligible. To account for factors, such as treatment hesitancy, which result in a delay between HIV diagnosis and ART initiation, all men are assigned a treatment delay period, $\theta$, upon HIV diagnosis (Figure 11). Specifically, $\theta,$ was calculated by subtracting the results from the Argus I and Engage survey question:

- “*when did you first start taking prescription medication to treat your HIV infection*?” from the results from the question “*when did you first test positive for HIV?*”.

The $\theta$ parameter was fitted to the combined Argus I (for men diagnosed between 1996 and 2005) and Engage (for men diagnosed from 2006 onwards) surveys. Each man was assigned a treatment delay period using a negative binomial distribution. The treatment delay period varied by year of diagnosis (1985-1995, 1996-2003, 2004-2012, 2013 onwards) (Eq. 11). The confidence intervals were calculated using 1,000 bootstrap replicates. Since ART was not available before 1996, the year of diagnosis was assumed to be 1996 for individuals who were diagnosed with HIV prior to 1996. It was assumed that subsequent to 2013, there was minimal delay between diagnosis and ART initiation such that individuals started ART within the same 2-week interval as HIV diagnosis.

$\theta_{t}\sim NB(\mu_{t}, s_{t})$	(11)

After treatment initiation, men are categorized as *sART* for a randomly assigned duration ranging from 3-6 months ^73^. At the end of the assigned period, men attain a status of *lART*. Neither *sART* nor *lART* transition between the HIV stages. Furthermore, *sART* and *lART* can transmit HIV at a reduced rate ($\epsilon_{ART\_sART}$ = 80% ^73^ and $\epsilon_{ART_{lART}}$ = 90-100% ^21^).

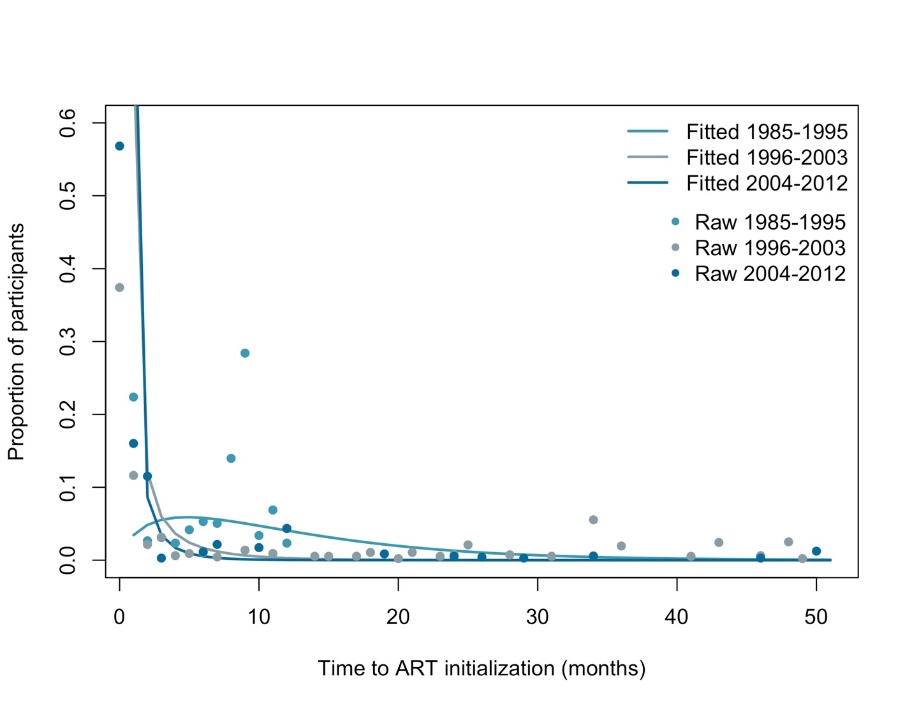

**Figure 11:** Delay between HIV diagnosis and time to anti-retroviral treatment (ART) initiation. The data was obtained from the Argus I (2005) and Engage (2017) population surveys and was stratified by year of diagnosis.

###### 3.5.3.1 ART discontinuation and re-initiation

Men who discontinue ART return to the CD4 cell count category at which they started ART and disease progression will resume at the same rate as ART-naïve men ^74^. The ART discontinuation rate was calculated using data from the *Argus II* survey. Namely, the discontinuation rate was measured from two questions in the *Argus II* survey:

- “*when did you first start taking anti-HIV medications?*”
- “*if you are not currently taking anti-HIV medications, when did you last take anti-HIV medication?*”

Both the ART discontinuation rate (Eq. 12), and the time varying ART re-initiation rates (Eq. 13) were converted to probabilities and were used in a Bernoulli distribution to determine which men discontinued or re-initiated ART use at each time step. The re-introduction of ART is followed by a delay period after which the man enters the *sART* category ^73^, followed by the *lART* category. The delay period was calculated as the inverse time between the first positive HIV test and ART prescription obtained from the *Argus I* and *Engage* surveys. The range was bootstrapped using 1000 replicates

$$ART\_discontinuation \sim Bernoulli\left( \psi\right) \left( 12 \right)$$

$$Art\_reinitiation_{t} \sim Bernoulli\left( \delta_{t} \right) (13)$$

##### 3.5.4 Condom use

Condoms can be used to protect individuals from transmitting or acquiring of HIV (with a 91.2% efficacy for anal sex ^75^) and other sexually transmitted infections. Longitudinal trends in condom use were based on the analysis of the *Argus I*, *Argus II*, and *Engage* surveys. The surveys asked individuals to report how often they used a condom with partners of different sero-statuses when they were either the insertive or receptive partner. These analyses indicate that condom use has changed over time, and varies based on age, sexual partnering level, sero-concordance, and partnership type (i.e., regular vs. casual).

Condom use prior to 1985 was assumed to be 1-5%. For the period between 1986 and 2005, condom use was shown to increase, however, given the scarcity of published data, an overall estimate was used for both regular and casual partnerships. Linear interpolation was used to infer the annual probability of condom use in between the respective time periods.

The probability of condom use at the remaining time points were inferred from data from the *Argus I*, *Argus II*, and *Engage* surveys. The following questions were used to calculate the probability of condom use for regular and casual partnerships:

- Argus I: *[For] sex with regular [HIV-positive/ unknown HIV status/ HIV-negative] male partners: during the past 6 months [when] you [fucked / were fucked by] these partners, how often was a condom used?*
- Argus II: *During the past 6 months, [if] have you [been fucked (bottom)/ fucked (top)] by an [occasional/ one night stand/ regular] partner who was [infected by HIV / not infected by HIV/ of unknown HIV status], how often was a condom used?*
- Engage: *[For] the most recent [5 men you have had sex with in the past 6 months]… The most recent time you had sex, when [he fucked you/ you fucked him], did [he/ you] use (or attempt to use) a condom?*

As the *Argus I* survey had no equivalent questions for casual partners, the *Argus II* survey questions were used to calculate the condom use probability for casual partners for the 2005-2016 period. Given the categorical answer scheme in the *Argus II* survey, the following assumptions were made: “never” = 0%, “occasionally” = 25%, “often” = 75%, and “always” = 100%. The average probability of condom use was calculated for each group.

With respect to age, participants were grouped into the following age groups: 15-24, 25-34, 35-49, and 50+ years. Although disclosure of HIV status is not explicitly modelled, the data was further stratified by sero-concordance within the partnership and the sexual partnering level for a randomly selected partner. For partnerships in which the men had different sexual partnering levels, one partner’s partnering category was chosen at random to determine whether the partnership was considered low, medium, or high partnering. For the purposes of this model, men whose HIV status was unknown were considered to be HIV negative. As a consequence of the complex stratification scheme, a referent group of men aged 35-49, with a low sexual partnering level who were in sero-concordant relationships was used for regular partnerships and the subgroup consisting of men aged 35-49, with high sexual partnering levels and sero-concordant relationships was used as the referent category for casual partnerships. Log odds multipliers were used to calculate the probability of condom use for the remaining subgroups (Table 12 and Eq. 14)

$P_{condom use\_t}= logit^{-1} (\mathrm{logit} \left( P_{referent\_t} \right)+\log\left( OR_{age} \right)+\log\left( OR_{sero-concordance} \right)+\log\left( OR_{sexual acitivty level} \right))$	(14)

Different condom use strategies were implemented in the model for casual and regular partnerships. It is assumed that only a proportion of partnerships use condoms. Furthermore, the per-act probability for men in casual partnerships that do use condoms is 100% (i.e., all-or-nothing) given that men in casual partnerships have 1-3 sex acts per partnership. In contrast, regular partnerships that use condoms do not use them 100% of the time. Since the collected data asked participants to indicate the proportion of sex acts in which condoms were used using a categorical scale of “*never*” (0% of the time), “*rarely*” (1-24%), “*sometimes*” (25-49%), “*most of the time*” (50-74%), “*almost every time*” (75-99%), and “*all the time*” (100%), these values were translated into the per act probability of condom use within a partnership (Table 13). A Bernoulli distribution was used to determine the per partnership probability of using condoms (Eq. 15) and a multinomial distribution was used to determine the per act probability of condom use for regular partners (Eq. 16).

$prob\_per\_partnership\_condom\_use_{t, partnership\_type} \sim Bernoulli({P_{condoms}}_{t, partnership\_type})$	(15)

$prob\_per\_act\_condom\_use_{t} \sim Multi(P_{act\_condoms_{t}})$	(16)

**Table 12:** Per partnership probability of condom use stratified by time, type of relationship, serostatus, and sexual partnering level. The referent group for regular partnerships is men aged 35-49, with a low sexual partnering level who were in sero-concordant relationships. The referent group for casual partnerships is men aged 35-49, with a high sexual partnering levels and sero-concordant relationships. The odds ratio multipliers were calculated from the *Argus I*, *Argus II*, and *Engage* surveys.

| Probability of condom use | | | |
| --- | --- | --- | --- |
| Period | **Regular** **partner** | **Casual partner** | **Reference** |
| 1975-1985^1^ | [1.0 – 5.0%] | [1.0 – 5.0%] | Assumed |
| 1994^1^ | 60.0% ± 6.0% | 90.0% ± 9.0% | ^76^ |
| 1996^1^ | 66.0%± 6.6% | 86.2% ± 86.2% | ^77^ |
| 2004^2^ | 50.2% [39.2 - 59.7%] | 66.0% [57.7 -72.9%] | Argus I |
| 2009^2^ | 42.4% [33.5 - 51.0%] |  | Argus II |
| 2017 onwards^2^ | 30.0% [0.0 - 63.9%] | 42.3% [31.1 -54.9 %] | Engage |
| Condom use multiplier (odds ratio) | | | |
|  |  | **Regular** **partner** | **Casual partner** |
| Age | **15-24** | 2.23 | 1.30 |
|  | **25-34** | 1.48 | 1.00 |
|  | **35-49** | 1.00 | 1.00 |
|  | **50 +** | 1.00 | 0.95 |
| Sexual partnering level | **Low** | 1.00 | 1.79 |
|  | **Medium** | 7.59 | 0.81 |
|  | **High** | 6.54 | 1.00 |
| Sero-concordance | **Concordant** | 1.00 | 1.00 |
|  | **Discordant** | 1.80 | 1.31 |

^1^ The confidence intervals were calculated using the point probability ± 10%

^2^ The confidence intervals were calculated using 1000 bootstraps.

**Table 13:** Per act probability of condom use for men in a regular partnership for the respective proportion of sex acts protected with condoms. The probability is used in a multinomial distribution to determine the proportion of sex acts protected by a condom. Data from Table 12 were used to calculate each probability.

| **1983-1995** | | | | | | | | | | |
| --- | --- | --- | --- | --- | --- | --- | --- | --- | --- | --- |
| **Proportion^1^** | **100.0%** | | | | | | | | | |
| **Probability** | 100% | | | | | | | | | |
| **1996-2003** | | | | | | | | | | |
| **Proportion** | **25.0%** | | | **75.0%** | | **100.0%** | |  | | |
| **Probability** | 9% | | | 27% | | 64% | |  | | |
| **2004-2008** | | | | | | | | | | |
| **Sero-concordant** | | | | | | | | | | |
| **Proportion** | **6.3%** | **12.5%** | **16.7%** | | **18.8%** | **25.0%** | **33.3%** | | **37.5%** | **41.7%** |
| **Probability** | 0.2% | 2% | 0.2% | | 0.1% | 27% | 2% | | 0.8% | 0.7% |
| **Proportion** | **43.8%** | **50.0%** | **56.3%** | | **62.5%** | **66.7%** | **68.8%** | | **75.0%** | **81.3%** |
| **Probability** | 0.1% | 6% | 0.2% | | 5% | 0.2% | 0.4% | | 16% | 0.1% |
| **Proportion** | **83.3%** | **87.5%** | **100.0%** | |  |  |  | |  |  |
| **Probability** | 0.7% | 3% | 37% | |  |  |  | |  |  |
| **Sero-discordant** | | | | | | | | | | |
| **Proportion** | **8.3%** | **12.5%** | **25.0%** | | **37.5%** | **43.8%** | **50.0%** | | **56.3%** | **62.5%** |
| **Probability** | 0.1% | 2% | 29% | | 0.4% | 0.1% | 5% | | 0.1% | 5% |
| **Proportion** | **75.0%** | **87.5%** | **91.7%** | | **100.0%** |  |  | |  |  |
| **Probability** | 21% | 2% | 0.9% | | 36.% |  |  | |  |  |
| **2009 onwards** | | | | | | | | | | |
| **Sero-concordant** | | | | | | | | | | |
| **Proportion** | **6.0%** | **8.0%** | **12.0%** | | **18.5%** | **24.5%** | **30.8%** | | **33.3%** | **37.0%** |
| **Probability** | 2% | 0.1% | 9% | | 0.7% | 2% | 0.1% | | 1% | 9% |
| **Proportion** | **41.3%** | **43.5%** | **46.8%** | | **49.5%** | **50.0%** | **55.8%** | | **56.0%** | **62.0%** |
| **Probability** | 0.2% | 0.4% | 0.1% | | 4% | 0.5% | 0.5% | | 0.3% | 7% |
| **Proportion** | **66.3%** | **68.3%** | **68.5%** | | **70.7%** | **71.5%** | **74.5%** | | **80.8%** | **81.0%** |
| **Probability** | 0.2% | 0.2% | 0.6% | | 0.3% | 0.1% | 2% | | 0.2% | 0.7% |
| **Proportion** | **84.0%** | **87.0%** | **93.5%** | | **95.7%** | **96.8%** |  | |  |  |
| **Probability** | 0.2% | 12% | 1% | | 0.1% | 0.9% |  | |  |  |
| **Sero-discordant** | | | | | | | | | | |
| **Proportion** | **6.0%** | **8.0%** | **12.0%** | | **18.5%** | **24.5%** | **37.0%** | | **43.5%** | **49.5%** |
| **Probability** | 2% | 0.1% | 9% | | 0.4% | 2% | 9% | | 0.5% | 3% |
| **Proportion** | **50.0%** | **56.0%** | **62.0%** | | **68.5%** | **74.5%** | **81.0%** | | **87.0%** | **93.5%** |
| **Probability** | 0.3% | 0.8% | 9% | | 0.5% | 3% | 0.1% | | 13% | 2% |
| **Proportion** | **100.0%** |  |  | |  |  |  | |  |  |
| **Probability** | 47% |  |  | |  |  |  | |  |  |

**^1^Proportion:** Proportion of sex acts protected by a condom

##### 3.5.5 Pre-exposure prophylaxis (PrEP)

PrEP became available for use among gbMSM in Québec in 2013. Following the guidance provided in Québec guidelines for PrEP initiation ^78,79^, from 2013 onward, HIV-negative men in the model were considered eligible to use PrEP if they had any of their anal sex acts unprotected by condoms in the past six months and either:

1. Had two or more partnerships in the past six months; or
2. Ever used non-occupational PEP twice or more.

The parameterization of the PrEP module specifically utilized the *Engage* cohort data, as well as clinical data from the *L’Actuel PrEP cohort* ^30^, a prospective open cohort of patients consulting for and prescribed PrEP over 2013-2019 at Montréal’s *Clinique l’Actuel*.

The following question from *Engage* was used to determine PrEP uptake and inform the model parameterization:

- *“When did you first take PrEP?”*

Among the *Engage* participants eligible for PrEP according to the Québec criteria, annual PrEP uptake was estimated by the RDS-adjusted proportion that initiated PrEP in a given year. Inverse probability of censoring weights were used to adjust estimates from follow-up visits that could be affected by loss to follow-up. We determined the number of men eligible for PrEP at each study visit, with the baseline count from 2017 used as the denominator of the proportions in preceding years since we could not determine eligibility prior to this. No *Engage* participant reported initiating PrEP in 2013, and, consequently, the RDS-adjusted proportion could not be calculated for that year. As the annual proportion in 2014 was very low, and the confidence interval included zero, the probability of uptake in 2013 was assumed equal to the 2014 estimate. For all subsequent years, up to and including 2018, annual adjusted proportions were calculated. Due to an increased uncertainty related to loss to follow-up at the last available study visit and smaller sample size, the probability of uptake in 2019 was assumed equal to that estimated for 2018. For calibration, we allowed for additional uncertainty in these estimates.

The following clinical information from Clinique L’Actuel was used to determine PrEP persistence:

- *PrEP consultation and follow-up visit dates*
- *Reported initiation and/or discontinuation of PrEP following a PrEP consultation*

PrEP persistence was calculated as the median time from PrEP initiation to discontinuation. This estimate was age-standardized to Engage (it was not possible to additionally standardize based on ethnicity, as this data is not available for *l’Actuel*).

At each time step, PrEP initiation and discontinuation are determined by a Bernoulli distribution. Men who acquire HIV while on PrEP discontinue PrEP upon testing positive for HIV. As described above, the rate of PrEP uptake varies by year. Men who discontinue PrEP can re-initiate PrEP, provided that they again meet the eligibility criteria. Men taking PrEP get tested after one month on PrEP, and every three months thereafter, and it was assumed that all men on PrEP are 100% compliant with these testing requirements.

##### 3.5.6 Post exposure prophylaxis (PEP)

Within the model, men who engage in casual, condomless anal sex, can attempt to obtain non-occupational PEP if one of the following conditions are true:

1. The man is not diagnosed as living with HIV
2. The man is not already on PEP
3. The man is not on PrEP

The following question from the *Engage* survey was used to inform the model in which it was assumed that PEP use is equivalent to the attempted PEP use:

- “*in the past 6 months, have you tried to get PEP?*”

The PEP uptake rate was converted to a probability, and a Bernoulli distribution is used to determine which men attempt to get PEP at each time step, using the time varying rate, $\rho_{PEP_{t}}$ (Eq. 17). It was assumed that there was no PEP before 2001. Furthermore, the Engage survey was used to calculate the proportion who attempted PEP use in 2017. Linear interpolation is used to calculate $\rho_{PEP_{t}}$ between 2001 and 2017. The PEP uptake rate was held constant after 2017, in line with data from the *Engage* cohort. Since PEP eligibility requires a pre-prescription HIV negative test result ^70^, all men attempting to receive PEP are first tested and only those who receive a negative test result will be prescribed PEP. Men who are prescribed PEP are assumed to be 100% compliant in taking the drug for a 1-month period. Lastly, men are tested for HIV at 1 and 3 month periods after PEP initiation ^70^. A 68% compliance for post-PEP HIV testing was assumed ^69^.

$PEP\_use\sim Bernoulli(\rho_{PEP_{t}})$	(17)

##### 3.5.7 Circumcision

Circumcision reduces the risk of HIV acquisition among heterosexual couples ^80,81^. Given role versatility in gbMSM, the effect of circumcision could be different among this group ^80,81^. Several meta-analyses provided inconclusive results ^80-82^ but a potential protective effect could not be ruled out for gbMSM in low-income countries. Given the lack of conclusive evidence, circumcision is not considered in the model.

### 4 Model simulation

The model was implemented using a modular coding structure and was run in discrete time. Each module is run sequentially at each time step. The stochastic nature of the model results from the individual level occurrence of the respective event. For example, each module assesses the eligibility of each man in the population for the respective event (i.e., HIV testing, PrEP eligibility, etc.). A Bernoulli distribution is the used to determine whether the event occurs for each eligible man. If the specific event does occur to an individual, then their individual-level characteristics are updated to reflect their new status.

### 5 Table of parameters

**Table 14:** Model parameters and their prior distributions, if applicable.

| Parameter | Value | Prior distribution for calibrated parameters | Reference |
| --- | --- | --- | --- |
| Demographic parameters | | | |
| Initial model population size | 10,000 |  |  |
| Annual population growth rate | See Table 2 | fixed | ^32,33,36^ |
| Initial age distribution | See Table 3 | fixed | ^35^ |
| HIV prevalence in 1975 |  | ~U(0.02- 0.50%) | Assumed |
| Sexual behaviours | | | |
| Proportion of men whose sexual preference is “versatile” | 0.64 | fixed | *Engage* |
| Proportion of men whose sexual preference is “receptive” | 0.17 | fixed |  |
| Proportion of men whose sexual preference is “insertive” | 0.20 | fixed |  |
| Proportion of regular partnerships (≥1 month) $\boldsymbol{(}\boldsymbol{p}_{\boldsymbol{longterm}}\boldsymbol{)}$ |  | ~U(0.0 - 0.4) | Informed from *Argus I, Argus II*, and *Engage* |
| Number of anal sex partners per 6-month-period for the referent age group (15-20 years) |  | $\sim NB\left( \mu,s \right)$  $\mu$= [3.7 – 4.18]  $s= \left[ 0.56 - 1.29 \right]$ | *Engage Argus II Argus I* |
| Partner change rate scaling factor for men aged 20-24 years ($\boldsymbol{R}\boldsymbol{R}_{\boldsymbol{22-24}}$) | 1.22 | fixed |  |
| Partner change rate scaling factor for men aged 25-29 years ($\boldsymbol{R}\boldsymbol{R}_{\boldsymbol{25-29}}$) | 1.41 | fixed |  |
| Partner change rate scaling factor for men aged 30-34 years ($\boldsymbol{R}\boldsymbol{R}_{\boldsymbol{30-34}}$) | 1.54 | fixed |  |
| Partner change rate scaling factor for men aged 35-39 years ($\boldsymbol{R}\boldsymbol{R}_{\boldsymbol{35-39}}$) | 1.60 | fixed |  |
| Partner change rate scaling factor for men aged 40-44 years ($\boldsymbol{R}\boldsymbol{R}_{\boldsymbol{40-44}}$) | 1.56 | fixed |  |
| Partner change rate scaling factor for men aged 45-49 years ($\boldsymbol{R}\boldsymbol{R}_{\boldsymbol{45-49}}$) | 1.44 | fixed |  |
| Partner change rate scaling factor for men aged 50-54 years ($\boldsymbol{R}\boldsymbol{R}_{\boldsymbol{50-54}}$) | 1.26 | fixed |  |
| Partner change rate scaling factor for men aged 55-59 years ($\boldsymbol{R}\boldsymbol{R}_{\boldsymbol{55-59}}$) | 1.04 | fixed |  |
| Partner change rate scaling factor for men aged 60-64 years ($\boldsymbol{R}\boldsymbol{R}_{\boldsymbol{60-64}}$) | 0.81 | fixed |  |
| Partner change rate scaling factor for men aged 65-69 years ($\boldsymbol{R}\boldsymbol{R}_{\boldsymbol{65-69}}$) | 0.60 | fixed |  |
| Partner change rate scaling factor for men aged 70+ years ($\boldsymbol{R}\boldsymbol{R}_{\boldsymbol{70-}}\boldsymbol{)}$ | 0.42 | fixed |  |
| Duration of a regular partnership  ($\boldsymbol{duration\_long-term)}$ |  | $\sim NB\left( \mu,s \right)$  $\mu$= [1.00- 55.81]  s= [0.83 - 0.94] |  |
| Number of anal sex acts per short-term partnership | 1-3 | Not calibrated |  |
| Number of anal sex acts over a six month period per regular partnership (Poisson distributed) |  | $\lambda=$ [20.49 – 22.35] | *Engage* |
| HIV-disease progression parameters | | | |
| Per-act probability of HIV acquisition for insertive anal intercourse  $\boldsymbol{(}\boldsymbol{p}_{\boldsymbol{insertive}}\boldsymbol{)}$ |  | ~U(0.04-0.28%) | ^83^ |
| Relative risk of HIV acquisition for receptive anal intercourse $\boldsymbol{(R}\boldsymbol{R}_{\boldsymbol{receptive}}\boldsymbol{)}$ | 3.846 | fixed | ^83^ |
| Relative risk of HIV transmission during the acute infection stage ($\boldsymbol{R}\boldsymbol{R}_{\boldsymbol{CD}\boldsymbol{4}_{\boldsymbol{1}}}\boldsymbol{)}$ | 9.17 | fixed | ^84^ |
| Relative risk of HIV transmission when CD4^+^ cell counts < 200 $\boldsymbol{cells \mu}\mathbf{L}^{\mathbf{-1}}$ $\boldsymbol{(R}\boldsymbol{R}_{\boldsymbol{CD}\boldsymbol{4}_{\boldsymbol{2}}}\boldsymbol{)}$ | 7.27 | fixed |  |
| Fraction of individuals who transition from primary stage of infection to a CD4 cell count > 500 $\boldsymbol{cells \mu}\mathbf{L}^{\mathbf{-1}}$ ($\boldsymbol{p}_{\boldsymbol{0}}$) | 76% | fixed | ^53^ |
| Fraction of individuals going from primary stage to a CD4 cell count between 350 and 500$\boldsymbol{cells \mu}\mathbf{L}^{\mathbf{-1}}$($\boldsymbol{p}_{\boldsymbol{1}}$) | 19% | fixed |  |
| Fraction of individuals going from primary stage to a CD4 cell count between 200 and 350$\boldsymbol{cells \mu}\mathbf{L}^{\mathbf{-1}}$($\boldsymbol{p}_{\boldsymbol{2}}$) | 5% | fixed |  |
| Transition rate from the primary stage of infection to a CD4 cell count > 500 $\boldsymbol{cells \mu}\mathbf{L}^{\mathbf{-1}}$  ($\boldsymbol{\gamma}_{\boldsymbol{1}}\boldsymbol{)}$ | $\frac{1}{0.25}$ year^-1^ | fixed |  |
| Transition rate from a CD4 cell count > 500 $\boldsymbol{cells \mu}\mathbf{L}^{\mathbf{-1}}$ to a CD4 cell count between 350 and 500$\boldsymbol{cells \mu}\mathbf{L}^{\mathbf{-1}}$  ($\boldsymbol{\gamma}_{\boldsymbol{2}}\boldsymbol{)}$ | $\frac{1}{3.32}$year^-1^ | fixed |  |
| Transition rate from a CD4 cell count between 350 and 500 $\boldsymbol{cells \mu}\mathbf{L}^{\mathbf{-1}}$ to a CD4 cell count between 200 and 350$\boldsymbol{cells \mu}\mathbf{L}^{\mathbf{-1}}$ ($\boldsymbol{\gamma}_{\boldsymbol{3}}\boldsymbol{)}$ | $\frac{1}{2.70}$year^-1^ | fixed |  |
| Transition rate from a CD4 cell count between 200 $\boldsymbol{cells \mu}\mathbf{L}^{\mathbf{-1}}$ and 350 $\boldsymbol{cells \mu}\mathbf{L}^{\mathbf{-1}}$ to a CD4 cell count <200$\boldsymbol{cells \mu}\mathbf{L}^{\mathbf{-1}}$  ($\boldsymbol{\gamma}_{\boldsymbol{4}}\boldsymbol{)}$ | $\frac{1}{5.50}$year^-1^ | fixed |  |
| HIV and AIDS related death rate ( $\boldsymbol{p}_{\boldsymbol{death\_age\_CD}\boldsymbol{4}}\boldsymbol{)}$ | See Tables 6 and 7 | fixed | ^55^ |
| Interventions | | | |
| Testing | | | |
| Proportion of men who are symptomatic when CD4 cell counts are <200$\boldsymbol{cells \mu}\mathbf{L}^{\mathbf{-1}}$  ($\boldsymbol{\rho}_{\boldsymbol{t,2}}\boldsymbol{)}$ | 93% | fixed | ^85^ |
| Proportion of men on PrEP who get tested every three months $\boldsymbol{(\rho}_{\boldsymbol{PrEP}}\boldsymbol{)}$ | 100% | fixed | Assumed |
| Proportion of men on PEP who get tested at 1 and 3 months after PEP initiation | 68% | fixed | ^69^ |
| Proportion of men ever tested for HIV | See Table 9 |  |  |
| Annual proportion of men tested for HIV | See Table 10 |  |  |
| HIV testing rate for the low partnering group  $\boldsymbol{(}\boldsymbol{\rho}_{\boldsymbol{tes}\boldsymbol{t}_{\boldsymbol{t}}\boldsymbol{l,}}\boldsymbol{)}$ | See Table 11 |  | *Engage Argus II Argus I* |
| Minimum time (months) between HIV tests |  | $\sim ZTPoisson\left( \lambda\right)$  $\lambda=\frac{1}{\rho_{test_{t}}}$ | Interpolation *Argus II* *Argus I Engage* |
| HIV test sensitivity and specificity | 100% | fixed | ^66^ |
| Testing interval for individuals on PrEP | 3 months | fixed | Assumed |
| Rate ratio for HIV testing rate ($\boldsymbol{\rho}_{\boldsymbol{tes}\boldsymbol{t}_{\boldsymbol{t}}}\boldsymbol{)}$ among the combined medium and high partnering individuals (>5partners per annum)  ($\boldsymbol{R}\boldsymbol{R}_{\boldsymbol{test}}$) | 1.46 | fixed | Calculated from the combined *Argus I*, *Argus II*, and *Engage* data |
| Period between HIV infection and the ability to detect HIV | See Table 8 |  | ^71^ |
| Treatment | | | |
| ART efficacy (for partial viral load suppression)  ($\boldsymbol{\epsilon}_{\boldsymbol{ART\_sART}}$) | 80% | fixed | ^50^ |
| ART efficacy (for full viral load suppression)  ($\boldsymbol{\epsilon}_{\boldsymbol{ART\_lART}}$) |  | ~U(90-100%) | ^21,48^ |
| Time delay (months) between HIV diagnosis and ART uptake (for diagnosis prior to 1996)  ($\boldsymbol{\theta}_{\boldsymbol{1996}}\boldsymbol{)}$ |  | $\sim NB\left( \mu,s \right)$  s = [0.10, 3.11]  $\mu$= [3.65, 6.80] | *Argus I*  *Engage* |
| Time delay (months)between HIV diagnosis and ART uptake (for diagnosis between 1996-2004)  ($\boldsymbol{\theta}_{\boldsymbol{1996-2004}}\boldsymbol{)}$ |  | $\sim NB\left( \mu,s \right)$  s = [0.14, 0.28]  μ= [12.30, 41.19] |  |
| Time delay (months)between HIV diagnosis and ART uptake (for diagnosis between 2004-2012)  ($\boldsymbol{\theta}_{\boldsymbol{2004-2012}}\boldsymbol{)}$ |  | $\sim NB\left( \mu,s \right)$  s = [0.06, 0.24]  $\mu$= [1.79, 10.11] |  |
| Time delay (months)between HIV diagnosis and ART uptake (for diagnosis after 2012) ($\boldsymbol{\theta}_{\boldsymbol{2012}}\boldsymbol{)}$ | Instantaneous | fixed |  |
| ART discontinuation rate  ($\boldsymbol{\psi}$) |  | ~U$\left( \frac{1}{113.91}, \frac{1}{89.40} \right)$ month^-1^ | Argus II |
| ART re-initiation rate (2005) $\boldsymbol{(}\boldsymbol{\delta}_{\boldsymbol{2005}}\boldsymbol{)}$ |  | ~U($\frac{1}{32.71},\frac{1}{5.4})$ month^-1^ | *Argus I* |
| ART re-initiation rate (2017) $\boldsymbol{(}\boldsymbol{\delta}_{\boldsymbol{2017}}\boldsymbol{)}$ |  | ~U($\frac{1}{3.28},\frac{1}{1.1}$) month^-1^ | *Engage* |
| Duration of short-term ART user categorization ($\boldsymbol{\phi)}$ | 3-6 months | Not calibrated | ^73^ |
| Condom use | | | |
| Condom efficacy | 91% | fixed | ^75^ |
| Per partner probability of condom use  (${\boldsymbol{P}_{\boldsymbol{condoms}}}_{\boldsymbol{t, partnership\_type}}\boldsymbol{)}$ | See Table 12 | fitted | *Engage Argus II Argus I Omega*  ^76,77^ |
| Per act probability of condom use (${\boldsymbol{P}_{\boldsymbol{act\_condoms}}}_{\boldsymbol{t, partnership\_type}}\boldsymbol{)}$ | See 13 | fixed | *Engage Argus II Argus I* |
| Pre-exposure prophylaxis (PrEP) | | | |
| PrEP efficacy ($\boldsymbol{\epsilon}_{\boldsymbol{PrEP}}$) | 86% | fixed | ^86^ |
| Proportion of all eligible men who initiate PrEP annually (2013 - 2014)  ($\boldsymbol{\vartheta}_{\boldsymbol{PrEP}_{\boldsymbol{2013}}}\boldsymbol{)}$ |  | ~U(0, 0.01) | *Engage* |
| Proportion of all eligible men who initiate PrEP annually (2015)  ($\boldsymbol{\vartheta}_{\boldsymbol{PrEP}_{\boldsymbol{2015}}}\boldsymbol{)}$ |  | ~U(0.01, 0.24) | *Engage* |
| Proportion of all eligible men who initiate PrEP annually (2016)  ($\boldsymbol{\vartheta}_{\boldsymbol{PrEP}_{\boldsymbol{2016}}}\boldsymbol{)}$ |  | ~U(0.01, 0.17) | *Engage* |
| Proportion of all eligible men who initiate PrEP annually (2017)  ($\boldsymbol{\vartheta}_{\boldsymbol{PrEP}_{\boldsymbol{2017}}}\boldsymbol{)}$ |  | ~U(0.01, 0.26) | *Engage* |
| Proportion of all eligible men who initiate PrEP annually (2018)  ($\boldsymbol{\vartheta}_{\boldsymbol{PrEP}_{\boldsymbol{2018}}}\boldsymbol{)}$ |  | ~U(0.06, 0.32) | *Engage* |
| Proportion of all eligible men who initiate PrEP annually (2019)  ($\boldsymbol{\vartheta}_{\boldsymbol{PrEP}_{\boldsymbol{2019}}}\boldsymbol{)}$ |  | ~U(0.06, 0.32) | *Engage* |
| PrEP discontinuation rate |  | ~U($\frac{1}{8.44},\frac{1}{13.00}$) month^-1^ | *l’Actuel* |
| Post-exposure prophylaxis (PEP) | | | |
| PEP efficacy $\boldsymbol{(}\boldsymbol{\epsilon}_{\boldsymbol{PEP}}\boldsymbol{)}$ | 80% | fixed | ^87^ |
| Proportion of all eligible men who attempted PEP use in 2000   ($\boldsymbol{\rho}_{\boldsymbol{PE}\boldsymbol{P}_{\boldsymbol{2001}}}\boldsymbol{)}$ | 0% | fixed | assumed |
| Proportion of all eligible men who attempted PEP use in 2017 onwards  ($\boldsymbol{\rho}_{\boldsymbol{PE}\boldsymbol{P}_{\boldsymbol{2017}}}\boldsymbol{)}$ |  | ~U(0.03, 0.11) | *Engage* |

### 6 Model calibration

An *Approximate Bayesian Computation Sequential Monte Carlo* (ABC-SMC) algorithm was used to calibrate the model. A detailed description of the ABC-SMC algorithm is described in ^88^. A 2-step procedure was used to calibrate the model. In the first step, the partnership dynamics modules were calibrated to sexual behaviour data. The resulting particle sets were used in the disease dynamics calibration process. Given the two-step process, there is potential variability in partnership dynamics after the second step of the calibration process. Therefore, the partnership dynamics outcomes were used as cross-validation targets in the final calibrated model. The calibration outcomes are presented below (Table 15).

**Table 15:** Table of calibration outcomes and cross validation outcomes

|  | **Type** | **Source** |
| --- | --- | --- |
| **Stage 1: partnership-level targets** | | |
| Distribution of number of anal sex partners that participants had in the past 6 months | Included in calibration | Combined data from: *Argus I, Argus II*, and *Engage* |
| Proportion of the population in a regular partnership | Included in calibration | Combined data from: *Argus II*, and *Engage* |
| Duration of regular partnerships | Included in calibration | Combined data from: *Argus I, Argus II*, and *Engage* |
| **Stage 2: disease and intervention-level targets** | | |
| Distribution of number of anal sex partners that participants had in the past 6 months | Cross-validation | Combined data from: *Argus I, Argus II*, and *Engage* |
| Proportion of the population in a regular partnership | Cross-validation | Combined data from: *Argus II,* and *Engage* |
| Duration of regular partnerships | Cross-validation | Combined data from: *Argus I, Argus II*, and *Engage* |
| Number of new diagnoses: 2002-2017^1^ | Cross-validation | INSPQ |
| Overall CD4 cell count at diagnosis by year: 2013-2017^1^ | Included in calibration | ^89-93^ |
| CD4 cell count at diagnosis by age:15-24, 25-34, 35-44, 45-54, 55+ and year: 2013-2017^1^ | Cross-validation | INSPQ |
| HIV prevalence by age (18-29, 30-49, 50 +) and year: 2005, 2008, 2017-2019 | Included in calibration | *Argus I* (self-reported), *Argus II* (self-reported), *Engage* (biomarker) |
| HIV incidence^2^ | Cross-validation | ^6,94,95^ |
| Lifetime use of PrEP by year (%): 2017-2019 | Included in calibration | *Engage* |
| PrEP coverage (current use) among HIV-negative men by year (%): 2017 -2019 | Included in calibration | *Engage* |
| PrEP coverage (use in the past six months) among HIV-negative men by year (%): 2017 -2019 | Cross-validation | *Engage* |
| ART coverage (%) by year (2005, 2017-2018)^3^ | Included in calibration | *Argus I* (self-reported), *Engage* (biomarker) |
| Proportion of PLHIV with a suppressed viral load (2017-2019) | Cross-validation | *Engage* |
| Proportion of PLHIV who are aware of their status^3^ | Included in calibration (years: 2005, 2008, 2017)  Cross validation (year: 2018) | ^96^ |

^1 We were unable to compute confidence intervals from the census data. Therefore, the point estimates were used to approximate the respective measure.
2 The HIV incidence estimates represent national estimates 94,97 and estimates among higher risk individuals than are represented in this study 95, and are therefore used as an approximate measure. Caution should be used when comparing these estimates to the simulated incidence.
3 The 2019 estimates were excluded from the calibration process due to spurious estimates resulting from loss to follow-up.^

For both fitting stages, 100 particle sets were sampled in each population (Table 16). The model was simulated with each particle set from 1975-2019, using a two-week time step and a population size of 10,000. The first 50 particle sets for which the distance between the target outcomes and the simulation outcomes was less than $\epsilon$ were used to define the sampling space for the subsequent population. The prior distributions were defined using a uniform distribution (Table 14) and the perturbation kernel followed a Gaussian random walk. Up to 1000 parameter sets were sampled in each population. If after 1000 attempts the calibration process did not produce 50 eligible particle sets, then the calibration process was assumed to be completed.

**Table 16:** Approximate Bayesian Computation Sequential Monte Carlo calibration settings and parameters. The fitting was accomplished in two stages: 1) partnership dynamics, and 2) disease dynamics.

|  | **Partnership dynamics modules** | **Disease dynamics modules** |
| --- | --- | --- |
| Modules included | Birth, natural death, aging, partnership formation and dissolution, population growth | All, e.g., intervention strategies, disease dynamics, partnerships dynamics modules |
| Initial number of particle sets sampled ($n_{p}$) | 100 | |
| Accepted number of particle sets ($n_{a}$) | 50 | |
| Distance function | $\frac{\left\vert Modeled outcome- target outcome \right\vert}{Standard error of target outcome}$ | |
| Perturbation kernel | Gaussian walk with standard deviation = $\frac{CI_{upper}-CI_{lower}}{5}$ | |

#### 6.1 Posterior parameter ranges

Figures 12-17 show the prior and posterior distributions for the disease dynamics modules.

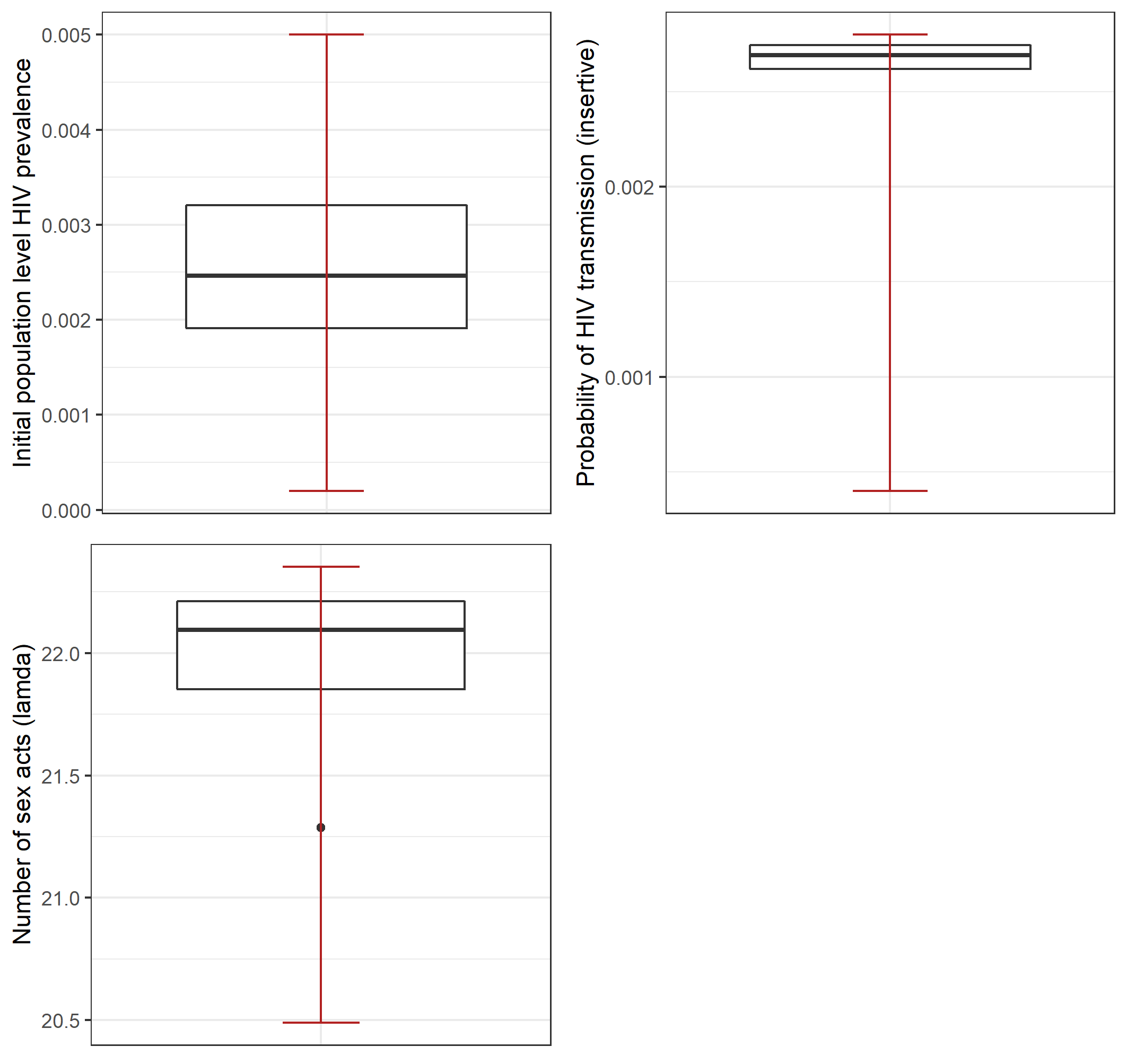
A)	B)

C)

**Figure 12**: Prior and posterior distributions for the disease dynamic modules. All prior distributions are represented with a red line and posterior distributions are represented using boxplots. A) Initial population level HIV prevalence, B) the probability of HIV transmission for insertive anal sex, C) the Poisson distribution lambda parameter used to assign the number of sex acts to men in a regular relationship.

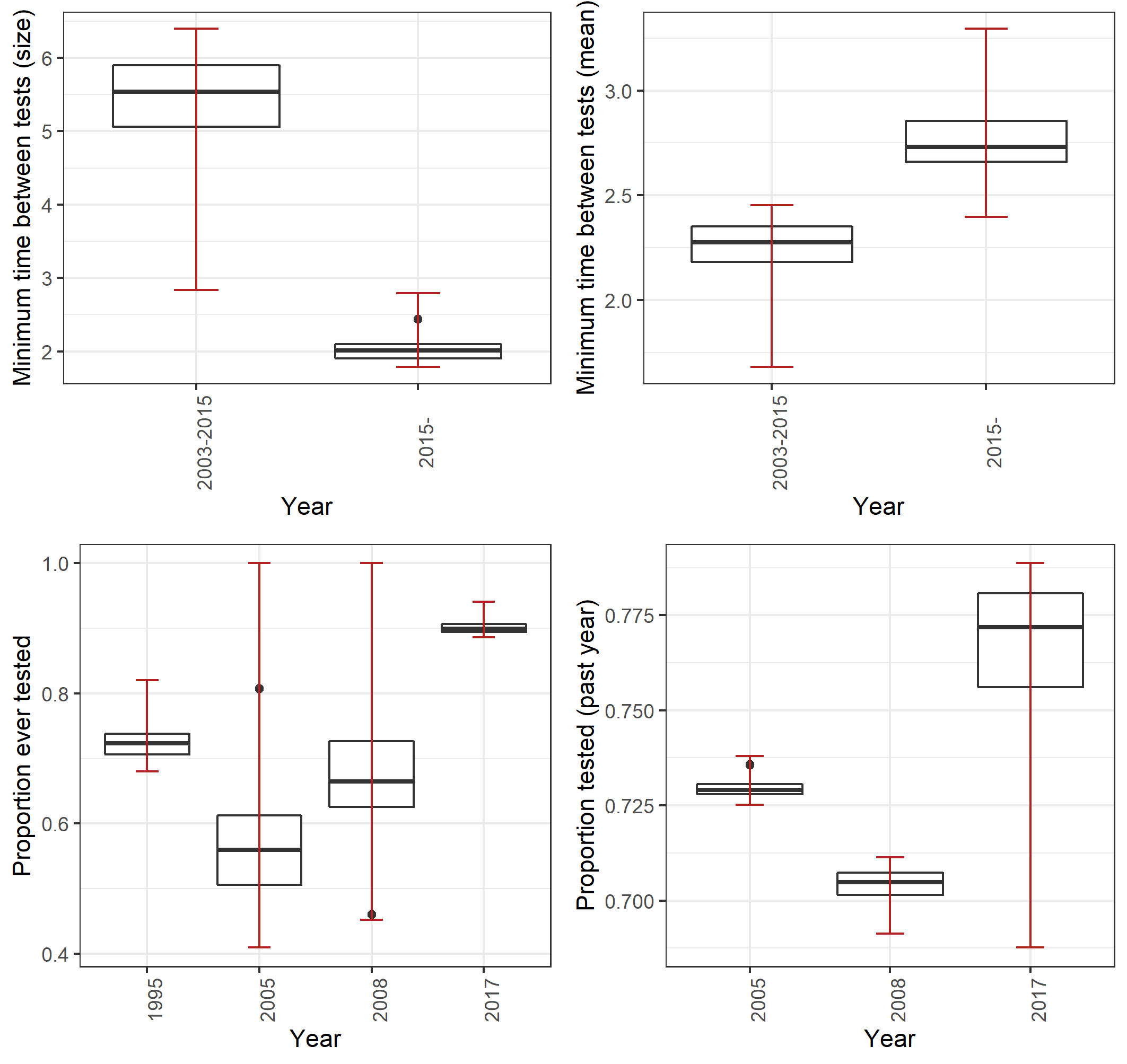
A)	B)

C)

**Figure 13**: Prior and posterior distributions for HIV testing parameters. All prior distributions are represented with a red line and posterior distributions are represented using boxplots. A) size parameter for the negative binomial distribution used to assign the minimum time between HIV tests, B) mean parameter for the negative binomial distribution used to assign the minimum time between HIV tests, C) the proportion of men who reported ever having received an HIV test, D) the proportion of men who received an HIV test within the respective year.

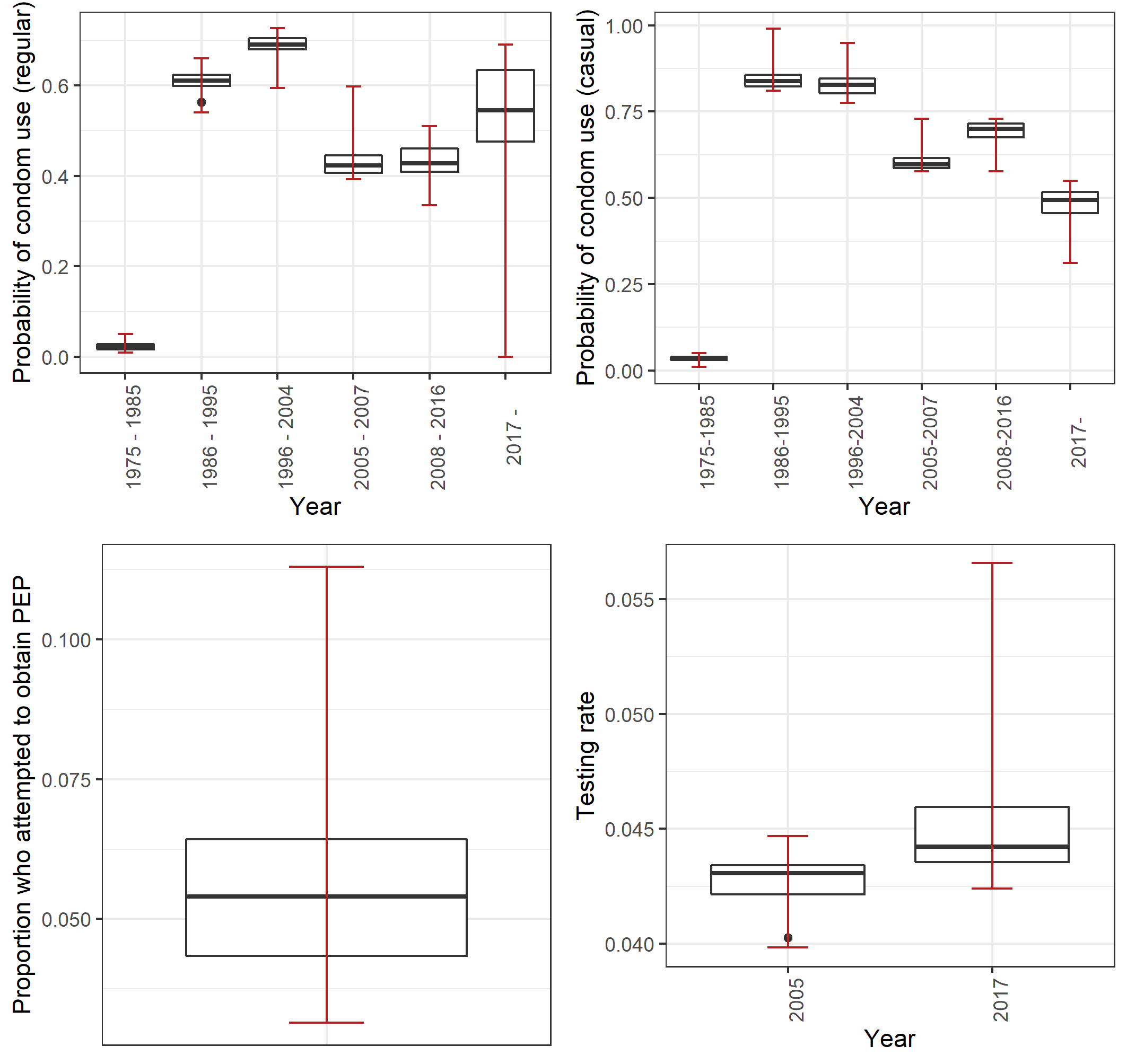
A)	B)

C)	D)

**Figure 14**: Prior and posterior distributions for the condom, PEP, and testing parameters. All prior distributions are represented with a red line and posterior distributions are represented using boxplots. A) Probability of condom use among regular partners, B) Probability of condom use among casual partners, C) the proportion who attempted to use post-exposure prophylaxis (PEP) in 2017, D) HIV testing rate

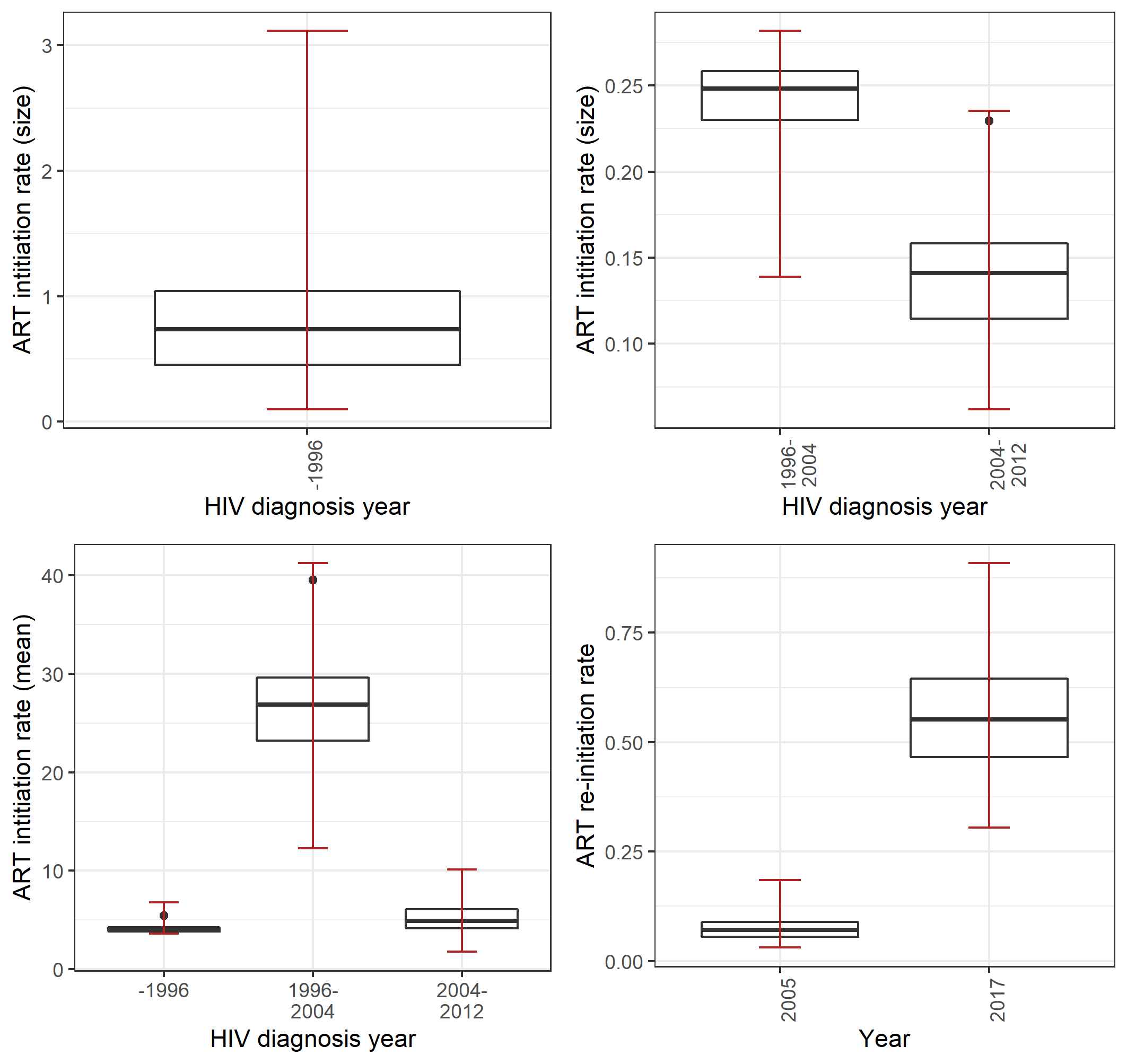

**Figure 15:** Prior and posterior distributions for disease dynamic modules. All prior distributions are represented with a red line and posterior distributions are represented using boxplots. A) Proportion who reported ever having received an HIV test, B) who reported having received an HIV test within the previous year.

B) negative binomial distribution size parameter for the period between HIV diagnosis and ART initiation, C) zero truncated negative binomial size parameter used to assign the minimum time between HIV tests, D) zero truncated negative binomial mean parameter used to assign the minimum time between HIV tests.

negative binomial distribution mean parameter for the period between HIV diagnosis and ART initiation, D) monthly ART re-initiation rate.

**Figure 16:** Prior and posterior distributions for anti-retroviral therapy (ART) parameters. All prior distributions are represented with a red line and posterior distributions are represented using boxplots. A) ART initiation rate; size parameter for the negative binomial distribution (for those diagnosed prior to 1996), B) ART initiation rate; size parameter for the negative binomial distribution (for those diagnosed after 1996), C) ART initiation rate; mean parameter for the negative binomial distribution, D) ART re-initiation rate

A)	B)

C)	D)

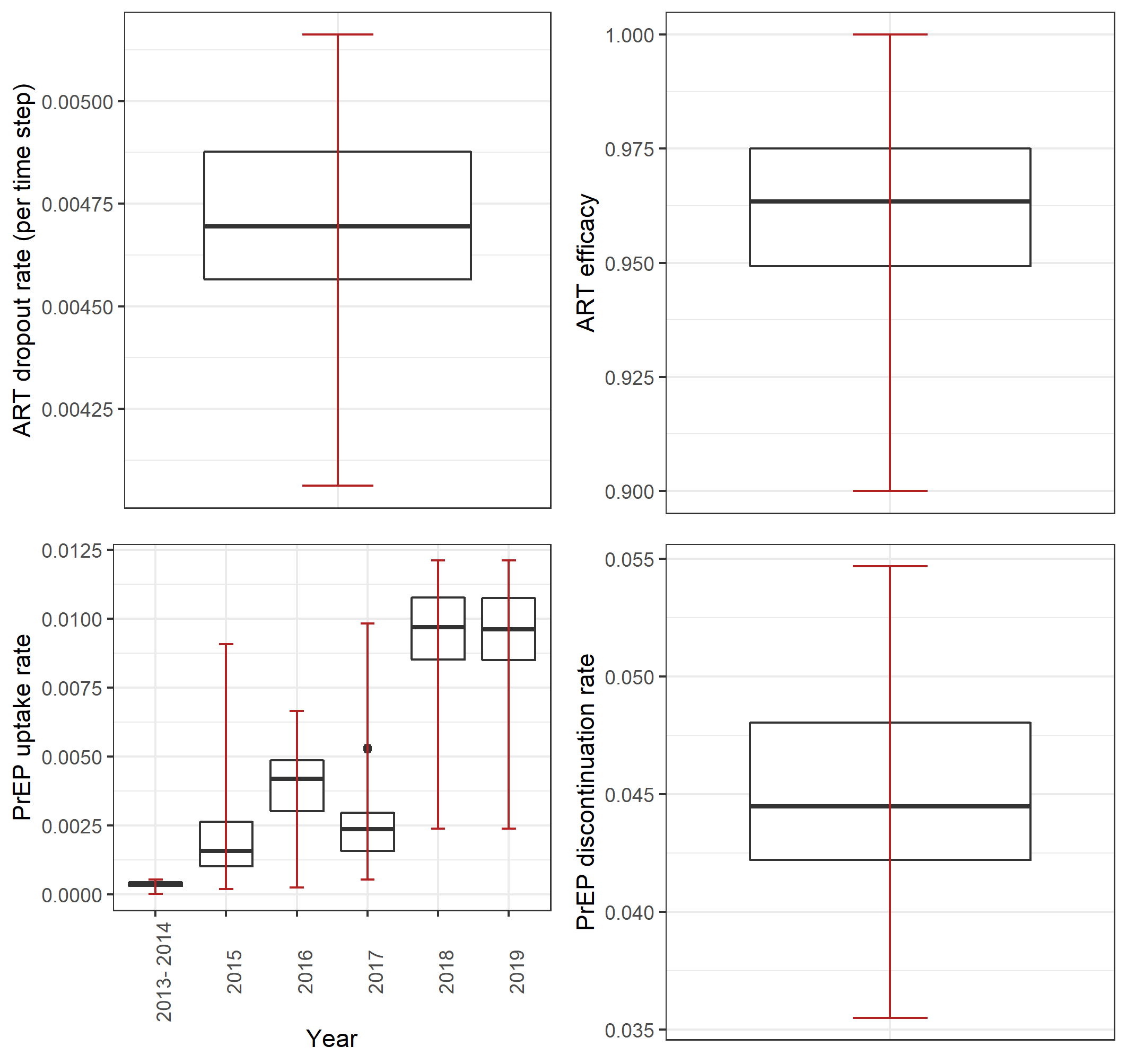
A)	B)

C)	D)

**Figure 17:** Prior and posterior distributions for anti-retroviral therapy (ART) and PrEP parameters. All prior distributions are represented with a red line and posterior distributions are represented using boxplots. A) ART discontinuation rate, B) ART efficacy for men with a fully suppressed viral load, C) Post-exposure prophylaxis (PrEP) uptake rate, D) PrEP discontinuation rate.

#### 6.2 Model fits

##### 6.2.1 Calibration targets

The final model calibration strategy resulted in 50 parameter sets. Each parameter set was simulated once in order to obtain the plots below.

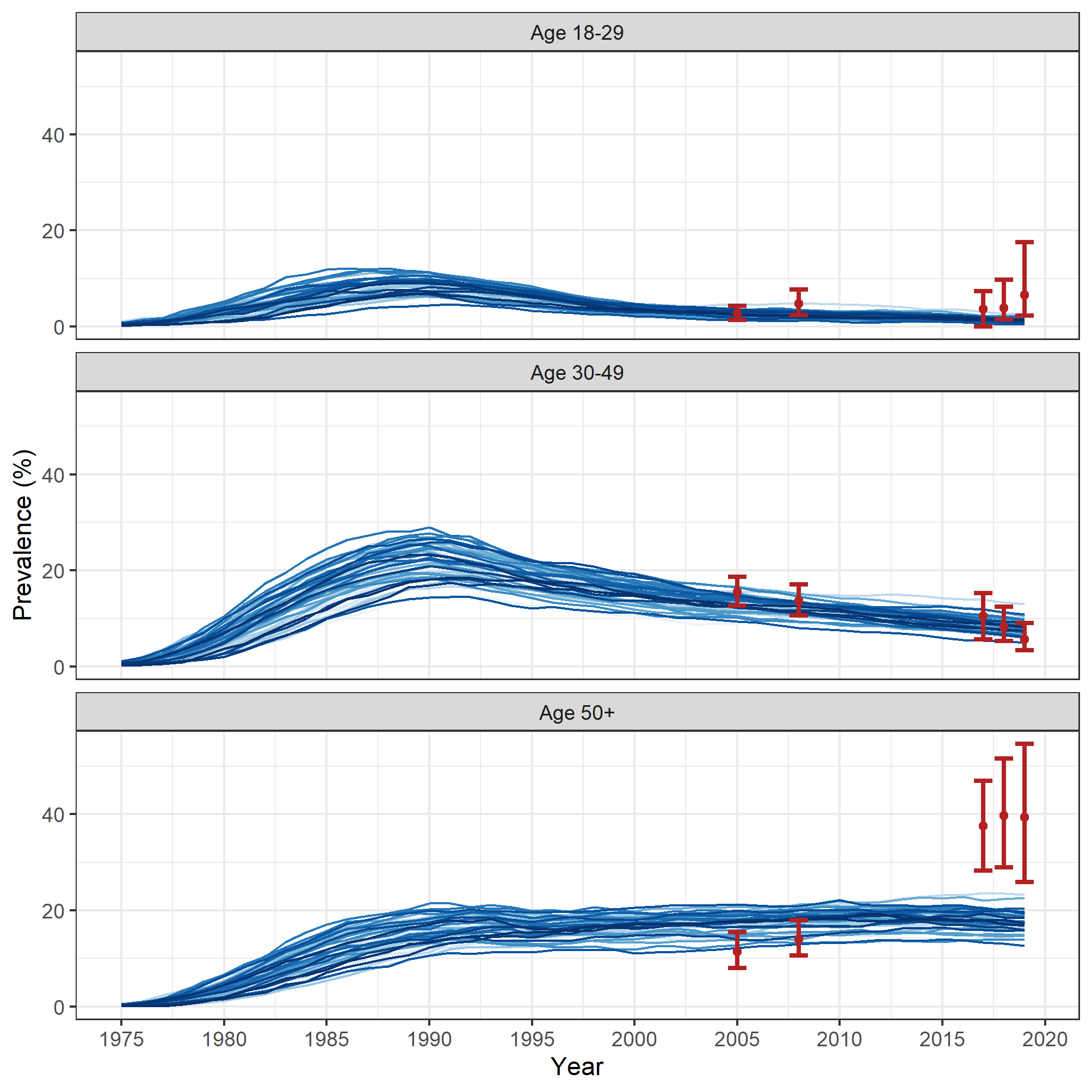
 **Figure 18:** Model calibration of the age-stratified HIV prevalence from 1975 until the end of 2019. The red bars represent the data. A) HIV prevalence among gbMSM 18-29 years. B) HIV prevalence among gbMSM 30-49 years. C) HIV prevalence among gbMSM 50 years and older.

A)

B)

C)

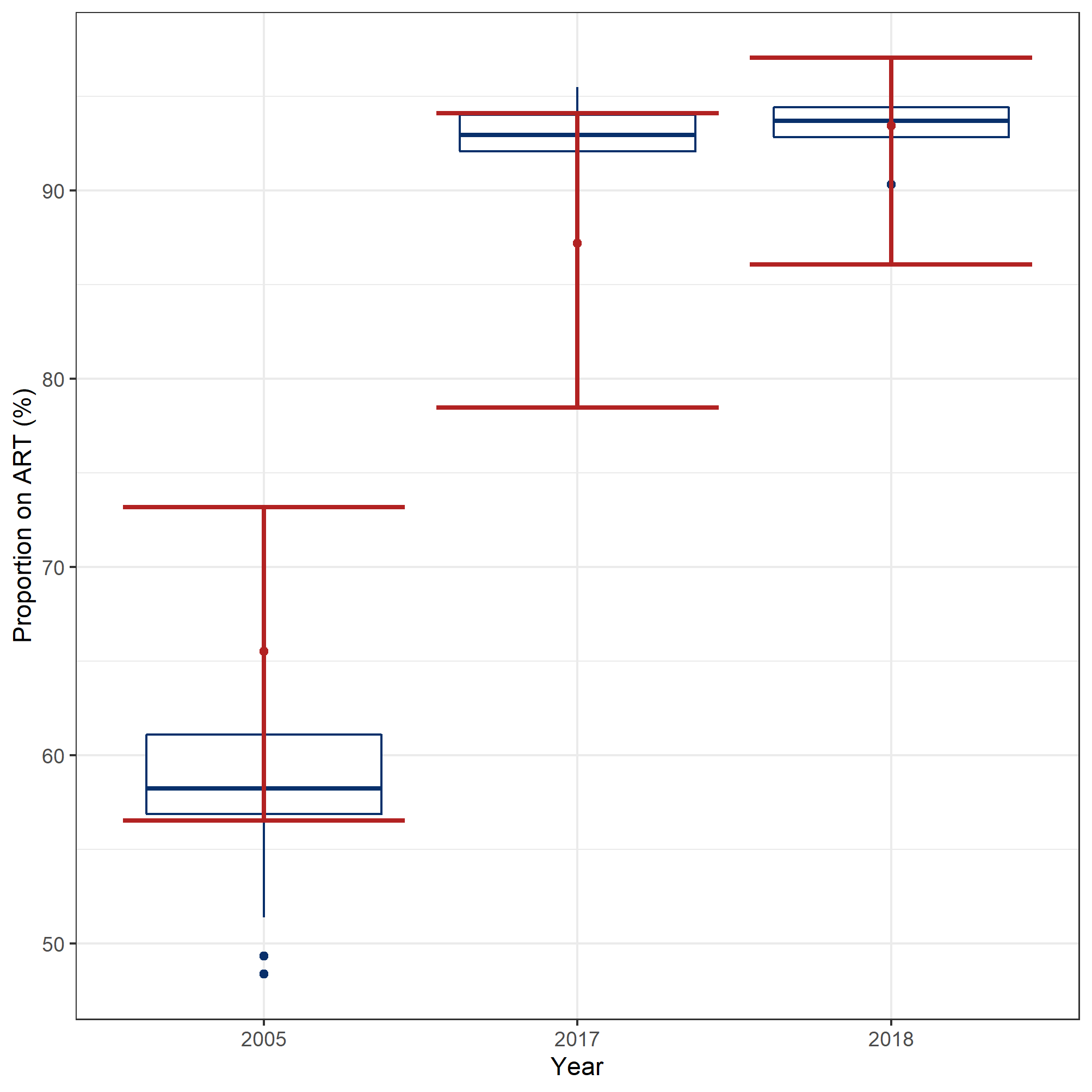

**Figure 19:** Calibration outcomes for the proportion of people living with HIV who are on anti-retroviral therapy (ART) in 2005, 2017, and 2018 The boxplots represent the model simulations and the red bars represent the calibration targets.

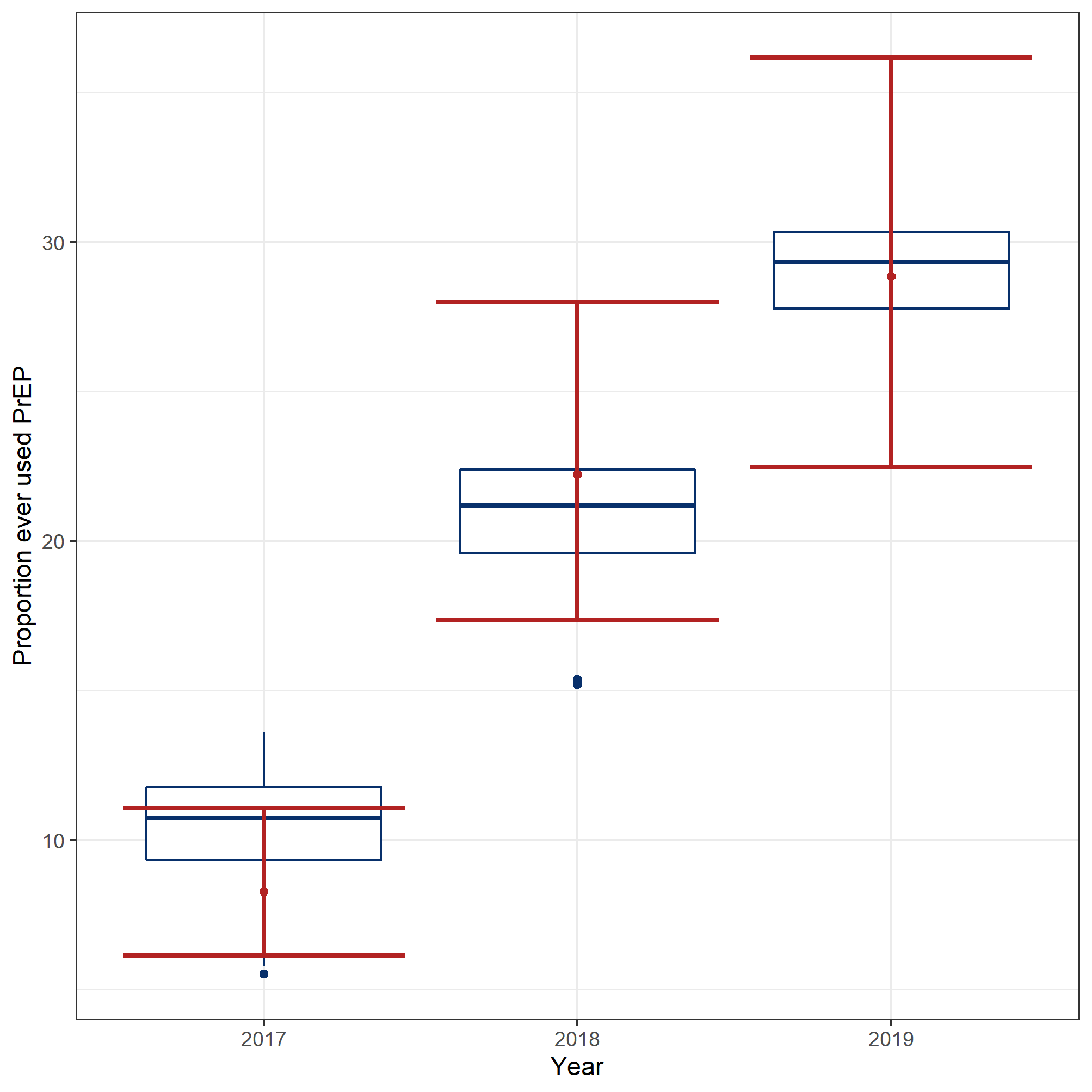

**Figure 20:** Calibration outcomes for the proportion of people who ever used pre-exposure prophylaxis (PrEP) in 2017-2019. The boxplots represent the modelled simulations and the red bars represent the calibration targets.

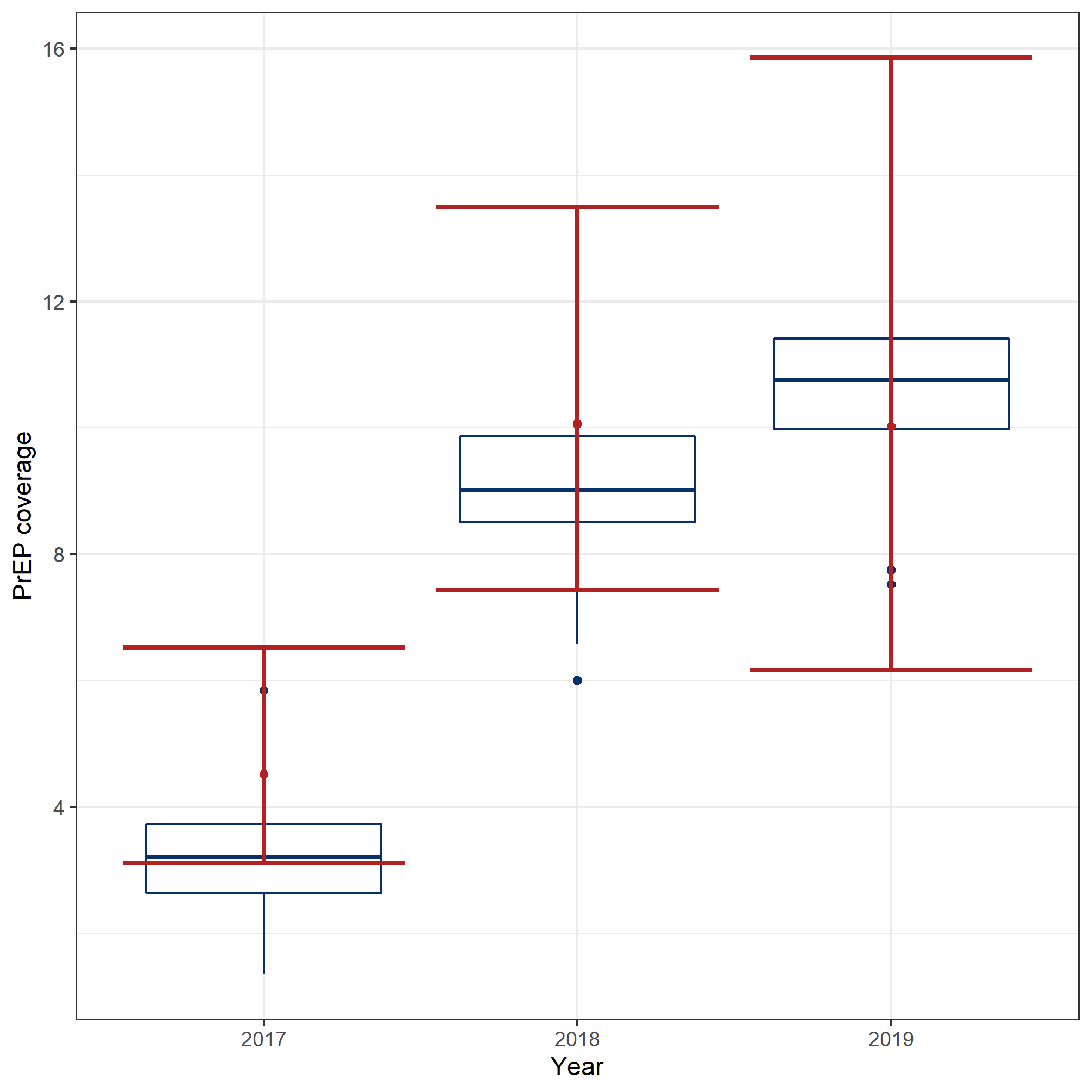

**Figure 21:** Calibration outcomes for the pre-exposure prophylaxis (PrEP) coverage among HIV-negative men in 2017-2019. The boxplots represent the model simulations and the red bars represent the calibration targets.

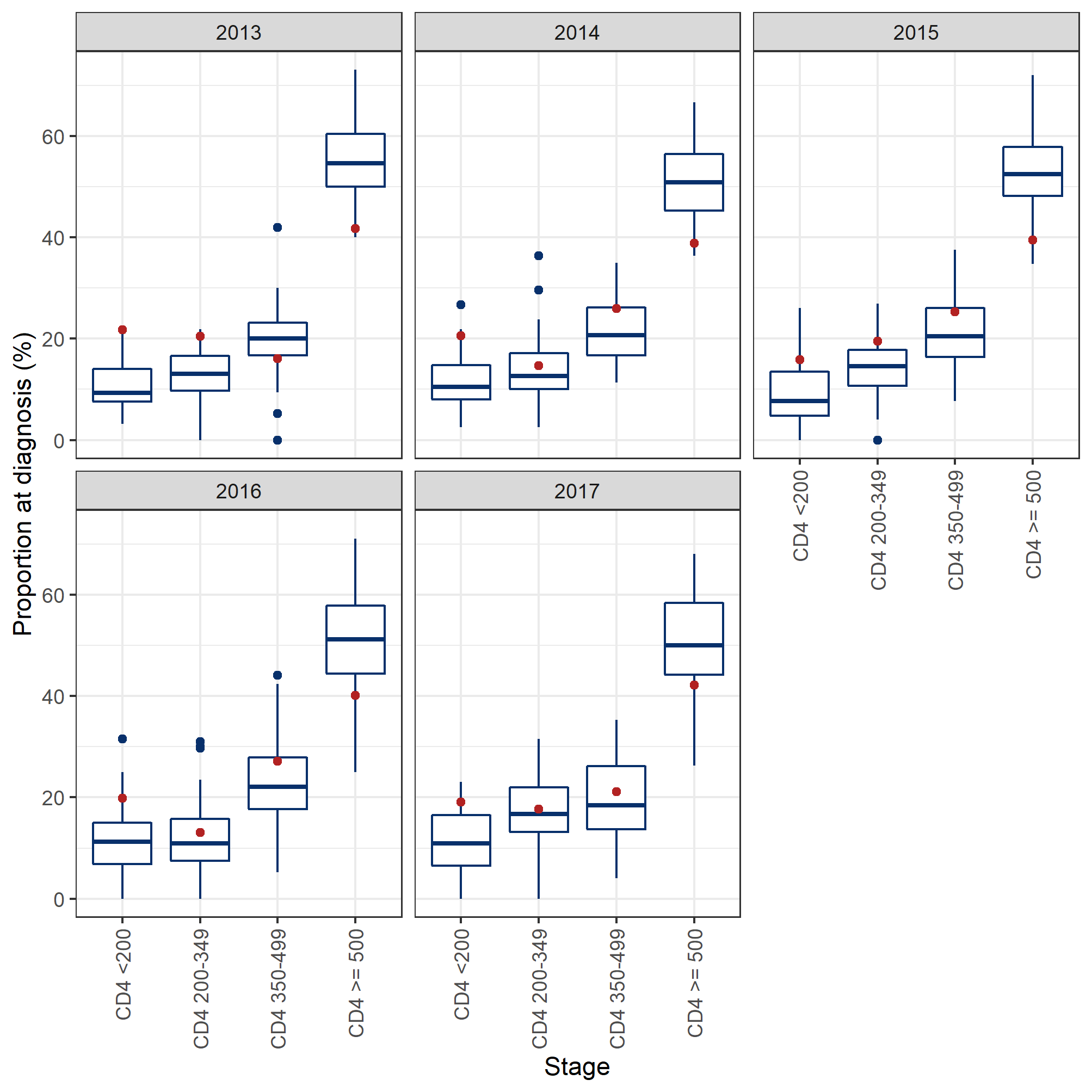

**Figure 22:** Calibration outcomes of the CD4 cell count at diagnosis between 2013 and 2017. The boxplots represent the model simulations and the red dots represent the data ^98^.

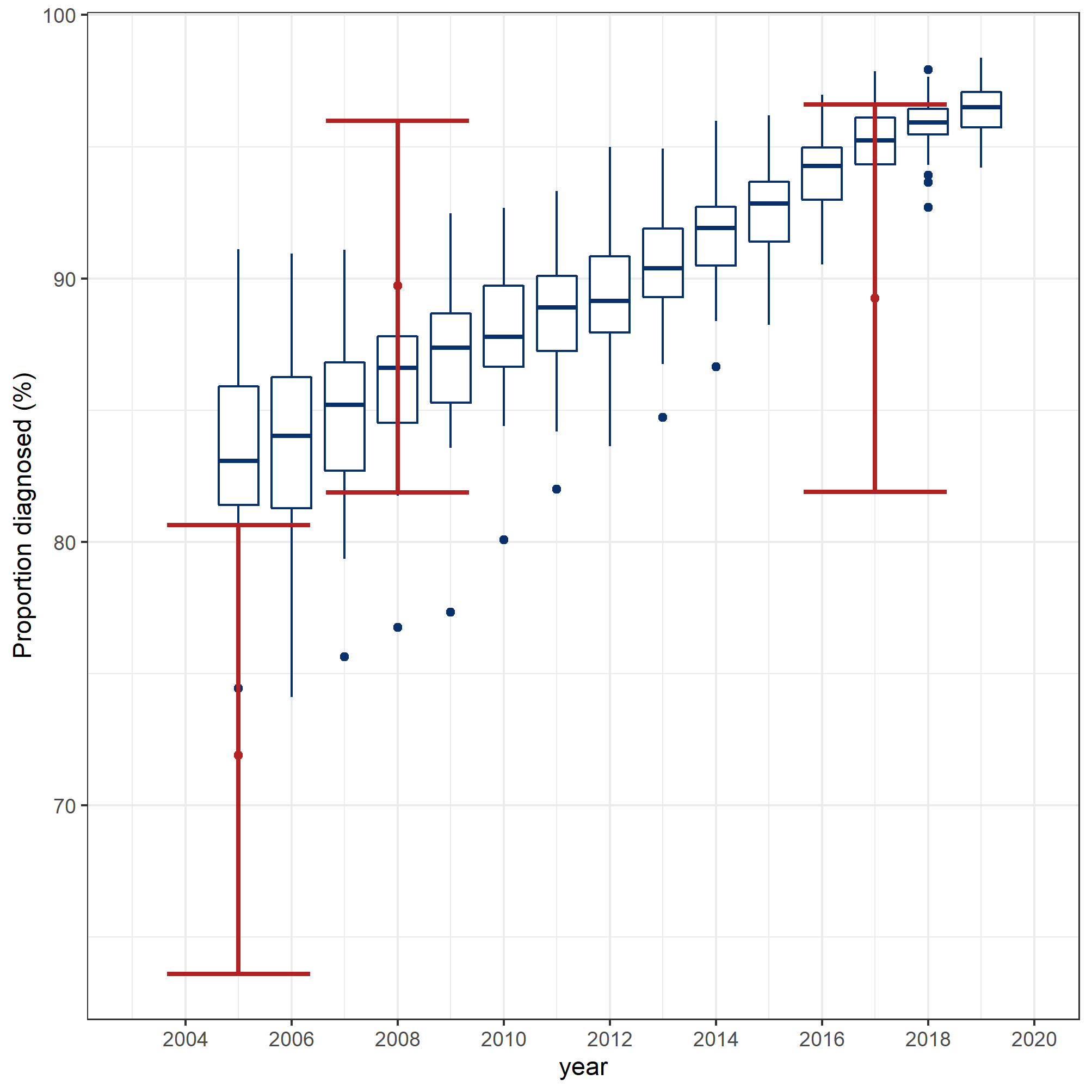

**Figure 23:** Calibration outcomes of the proportion with knowledge of status. The boxplots represent the modelled simulations and the red bars represent the data.

##### 6.2.2 Cross validation targets

###### 6.2.2.1 Partnership dynamics

The partnership dynamics modules were simulated once per parameter set to obtain the model fits below. The calibration outcomes included the proportion of regular partners, the number of anal sex partners that participants reported having within the past 6 months, and the duration of each long-term relationship. Regular partners were defined as relationships of duration 1 month or longer. The data was obtained from the combined *Argus I*, *Argus II*, and *Engage* surveys.

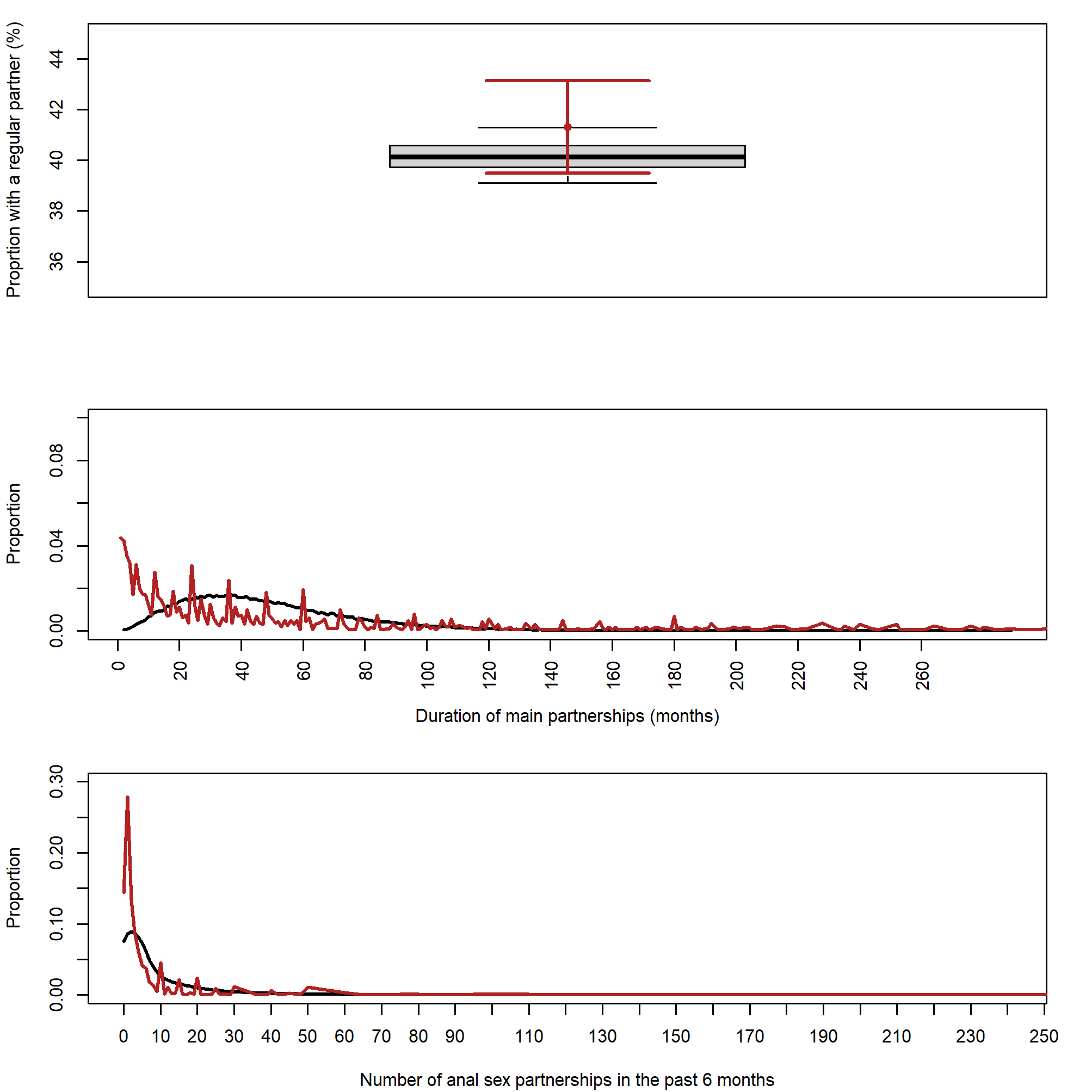

**A)**

**B)**

**C)**

**Figure 24:** Partnership dynamics calibration fits. The boxplot and black curves represent the simulated outcomes, and the red curves represent the data. A) The proportion of the population with a regular partner, B) the duration (months) of regular partnerships, and C) number of anal sex partners that men had in the past 6 months.

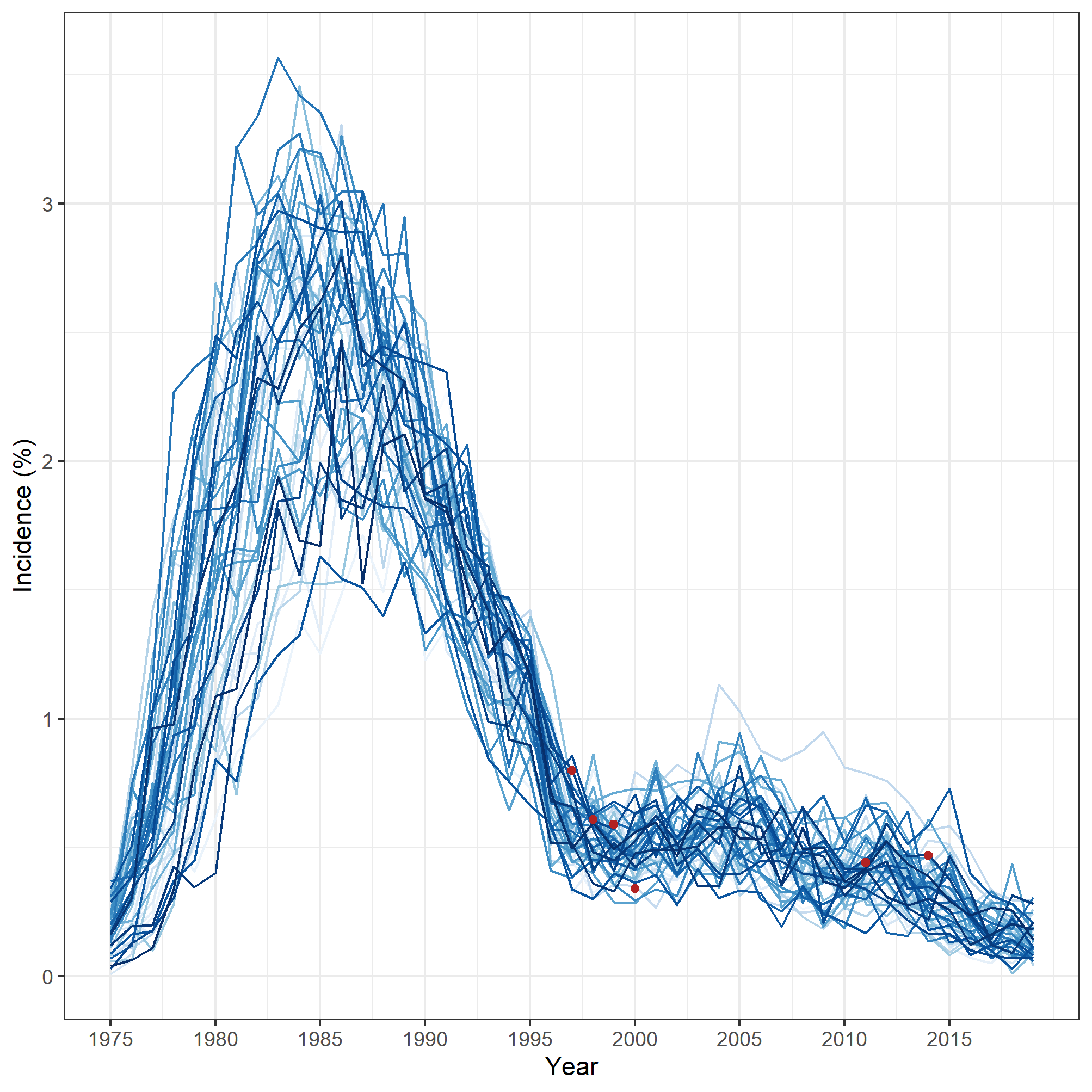

**Figure 25:** Calibrated HIV incidence among gbMSM in Montréal. Each line represents model simulations with an individual parameter set, while the red dots represent the empirical data ^6,94,95^.

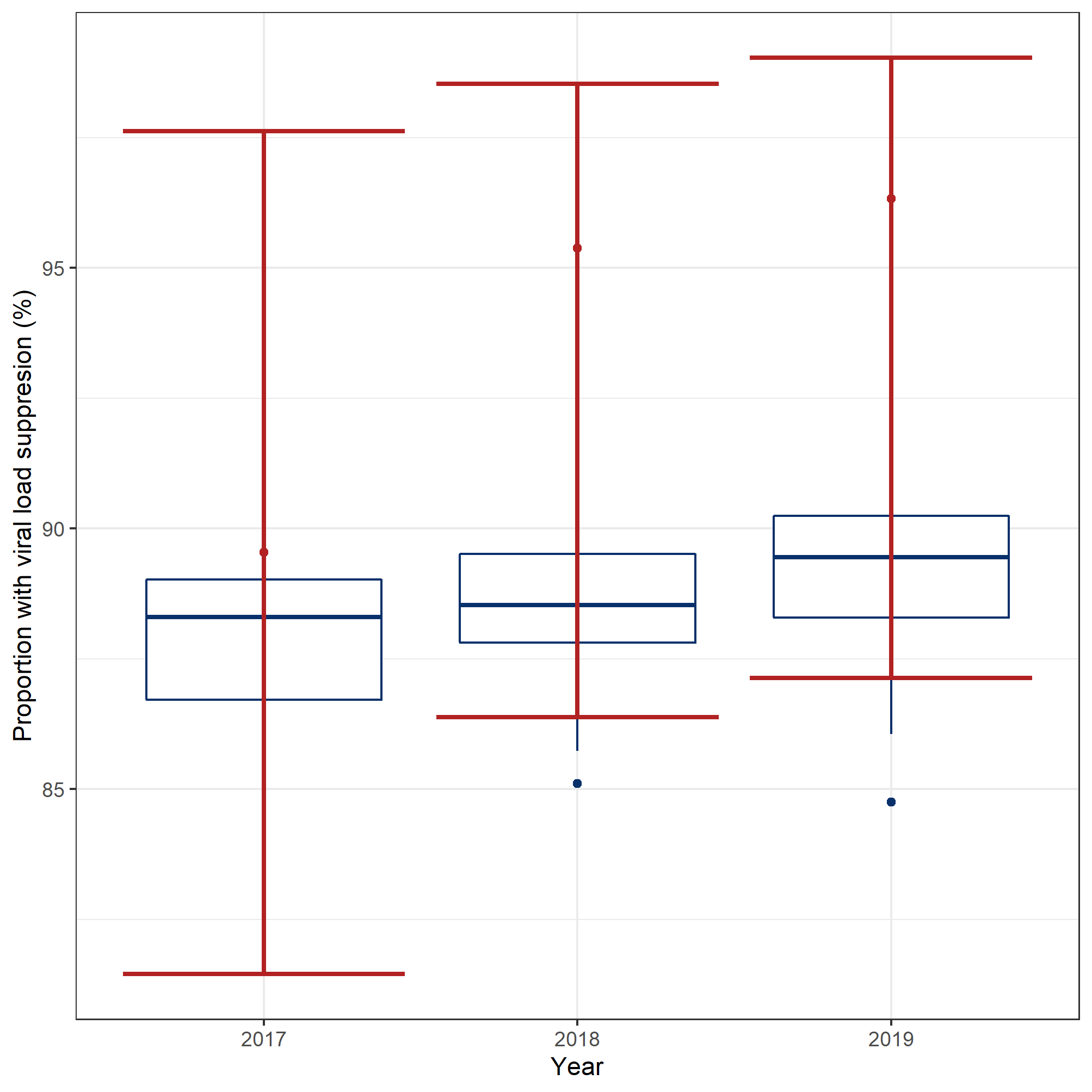

A)

B)

**Figure 26:** Calibrated proportion of people living with HIV who have attained viral load suppression. The boxplots represent the model simulations and the red lines represent the calibration targets obtained from the *Engage* cohort.

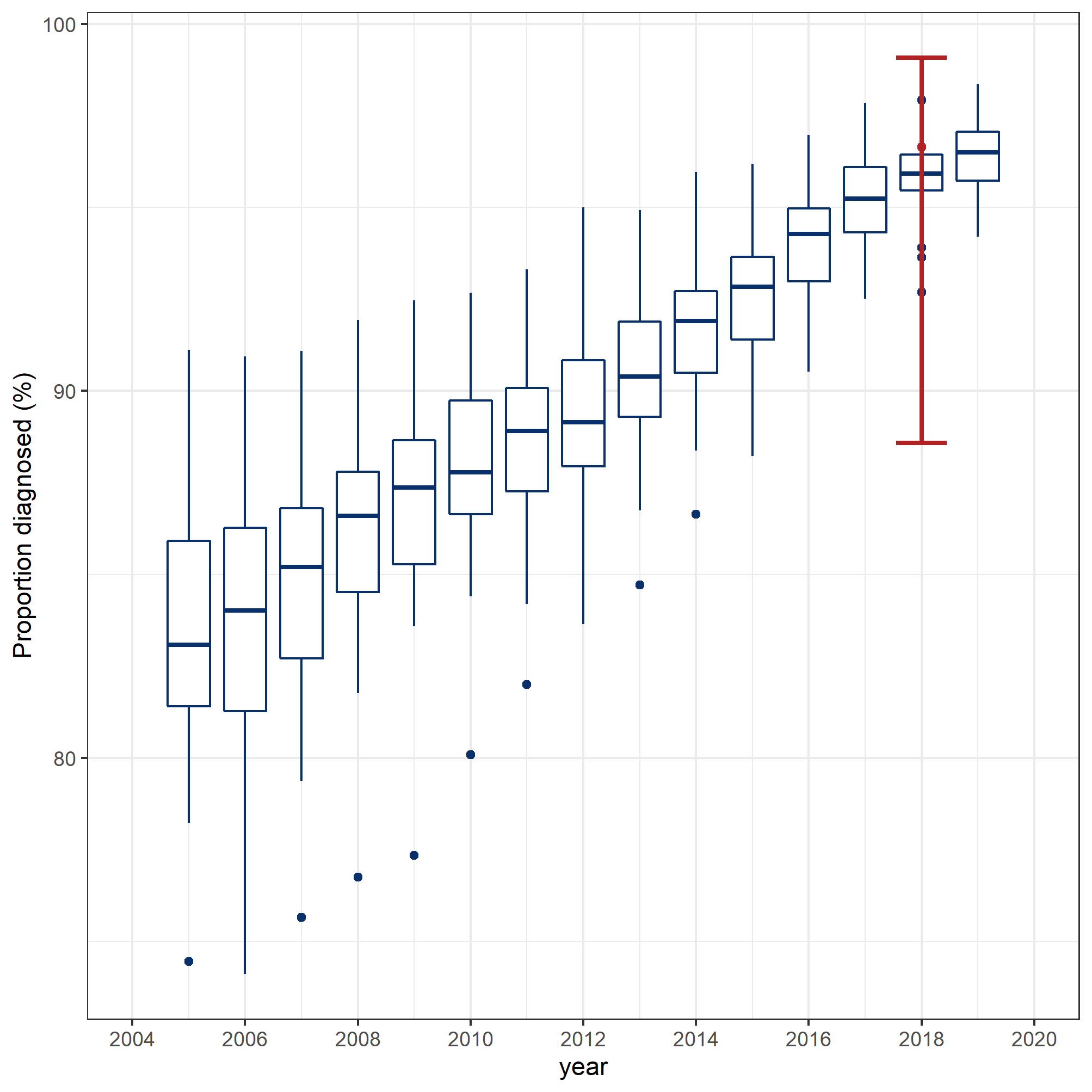

**Figure 27:** Calibrated proportion of people living with HIV who have knowledge of status (2019- cross-validation target). The boxplots represent the simulation outcomes and the red bars represent the baseline data from the *Engage* cohort.

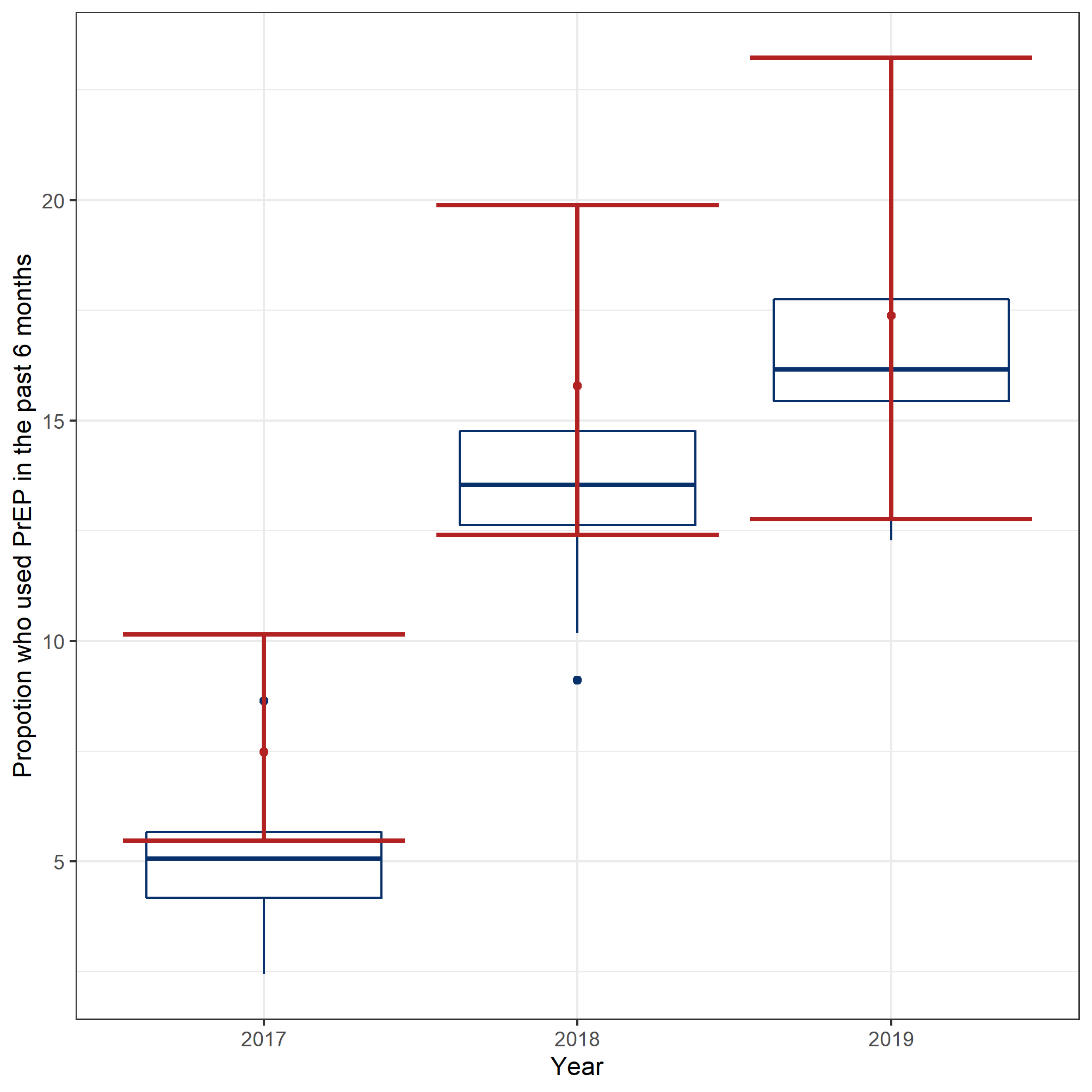

**Figure 28:** Calibrated PrEP use in the past six months. The boxplots represent the model simulations and the red bars represent the calibration targets. The proportion of HIV-negative men who reported using pre-exposure prophylaxis (PrEP) in the past six months in 2017, 2018, and 2019 in the *Engage* cohort.

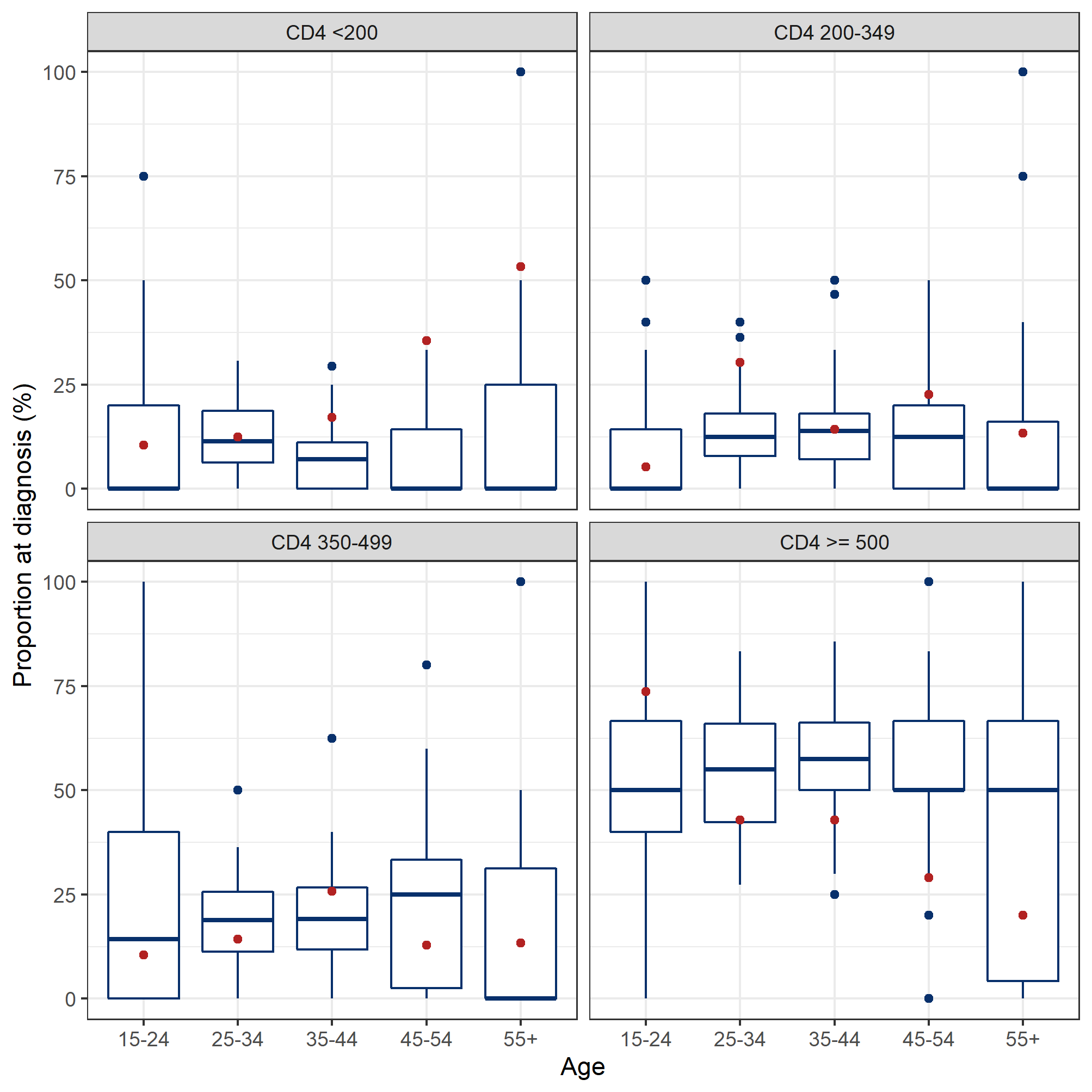

**Figure 29:** Simulated results of CD4 cell count at diagnosis in 2013. The boxplots represent the simulation outcomes and the red dots represent the surveillance data ^98^ (from the *Institut national de santé publique du Québec*). The model outcomes were stratified by 5 10-year age groups for each CD4 cell count level.

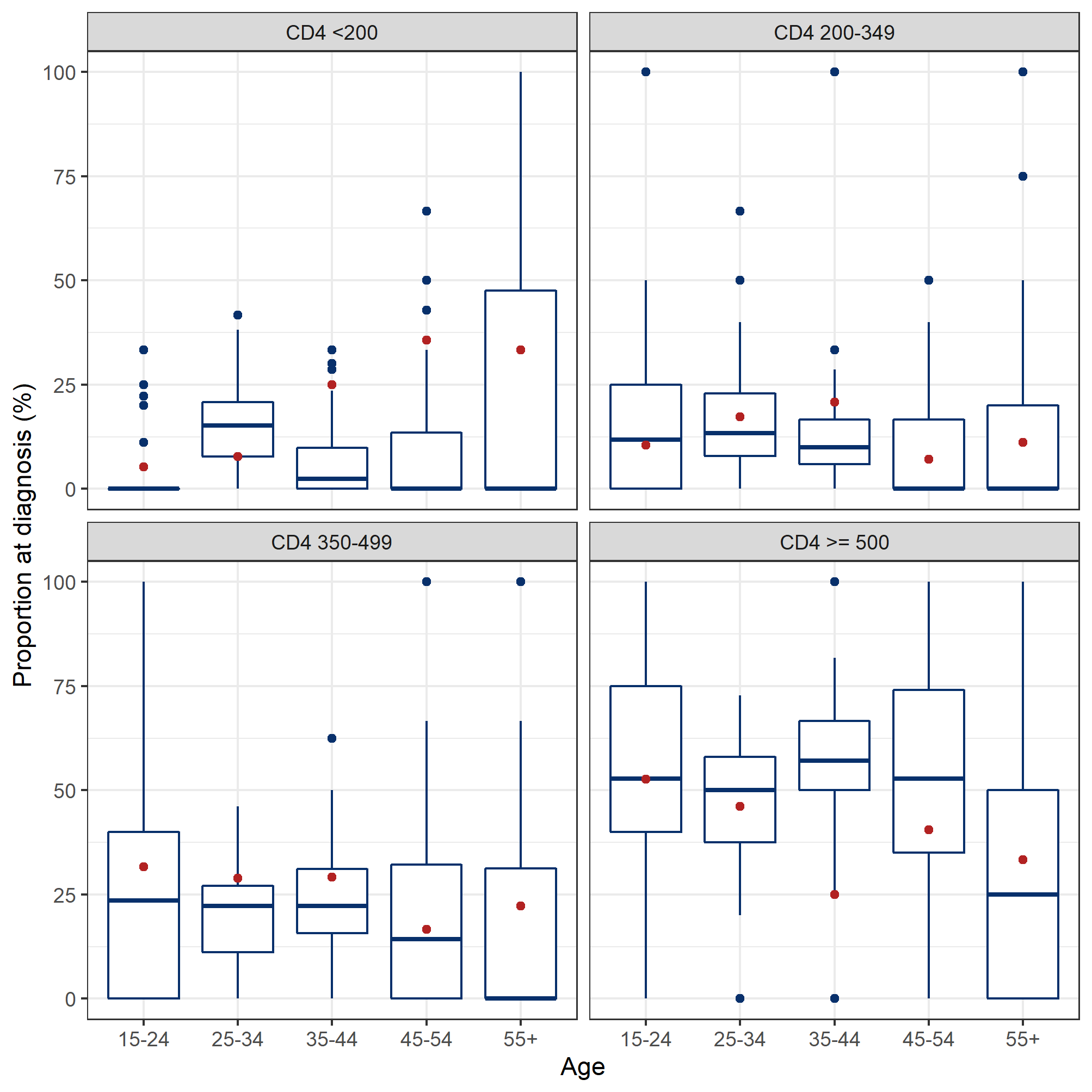
**Figure 30:** Simulated results of CD4 cell count at diagnosis in 2014. The boxplots represent the simulation outcomes and the red dots represent the surveillance data ^98^ (from the *Institut national de santé publique du Québec*). The model outcomes were stratified by 5 10-year age groups for each CD4 cell count.

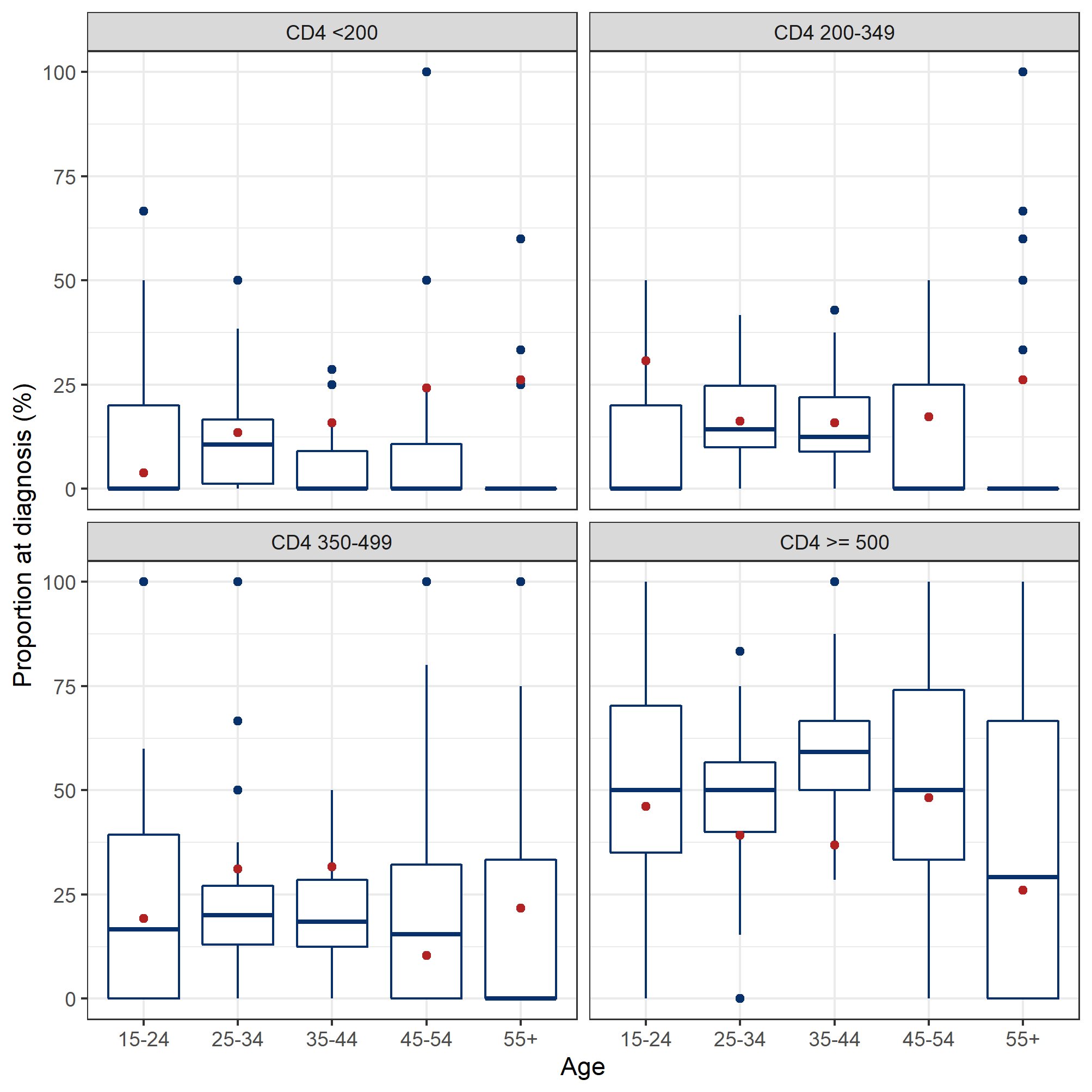

**Figure 31:** Simulated results of CD4 cell count at diagnosis in 2015. The boxplots represent the simulation outcomes and the red dots represent the surveillance data ^98^ (from the *Institut national de santé publique du Québec*). The model outcomes were stratified by 5 10-year age groups for each CD4 cell count.

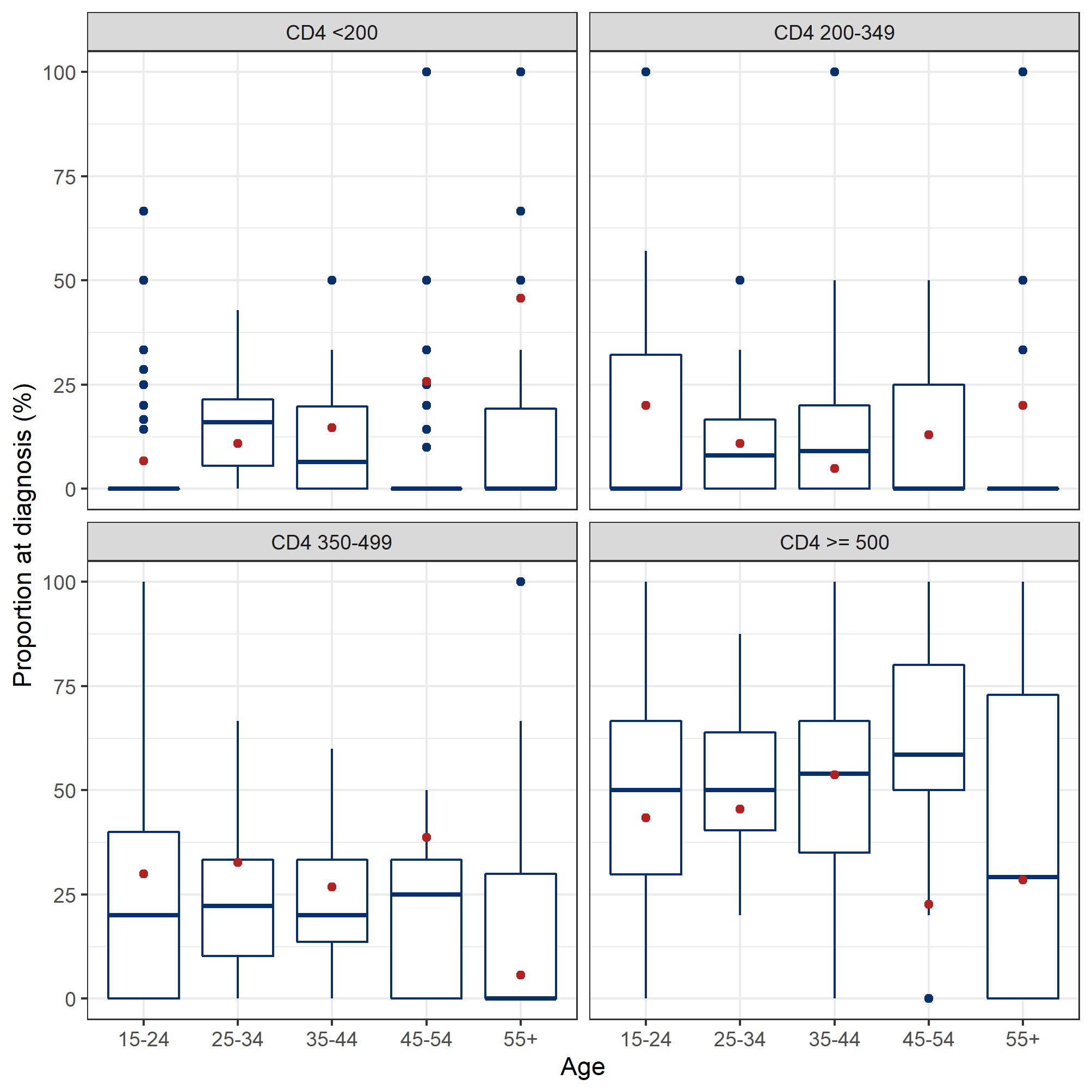
 **Figure 32:** Simulated results of CD4 cell count at diagnosis in 2016. The boxplots represent the simulation outcomes and the red dots represent the surveillance data ^98^ (from the *Institut national de santé publique du Québec*). The model outcomes were stratified by 5 10-year age groups for each CD4 cell count.

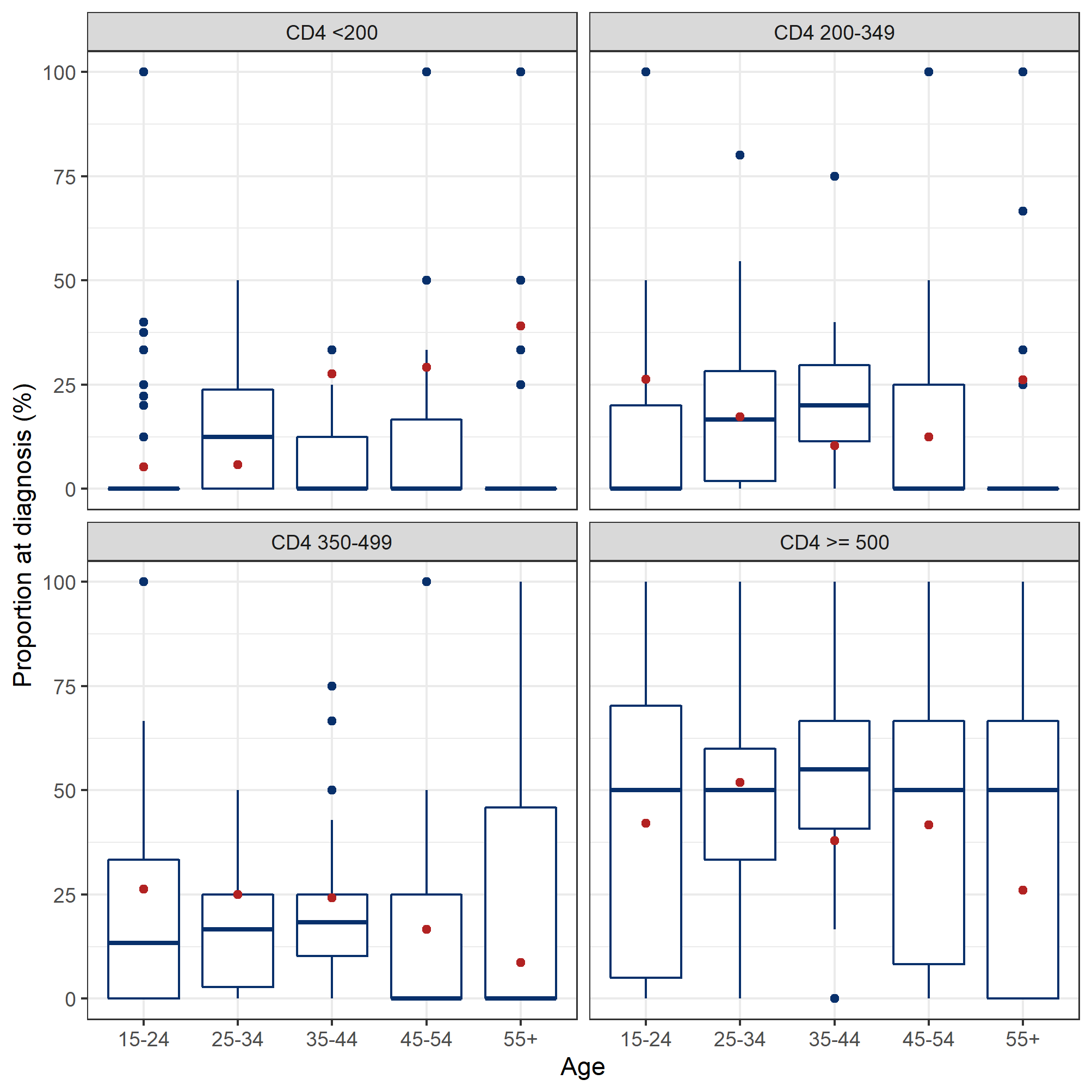
**Figure 33:** Simulated results of CD4 cell count at diagnosis in 2017. The boxplots represent the simulation outcomes and the red dots represent the surveillance data ^98^ (from the *Institut national de santé publique du Québec*). The model outcomes were stratified by 5 10-year age groups for each CD4 cell count.

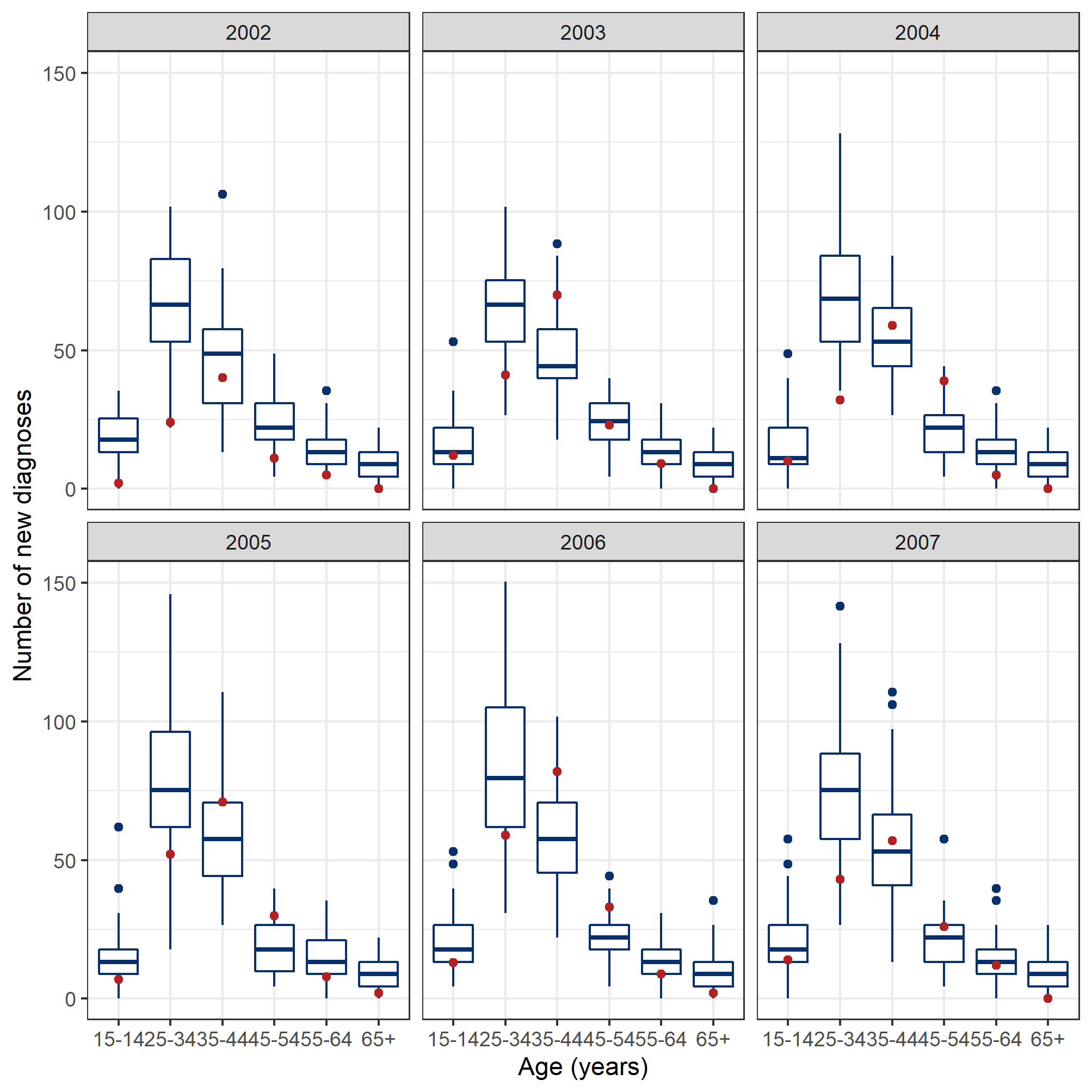
**Figure 34:** Simulated number of new diagnoses from 2002-2007. The boxplots represent the simulation outcomes and the red dots represent the surveillance data ^98^ (from the *Institut national de santé publique du Québec*). The model outcomes were stratified by 6 10-year age groups.

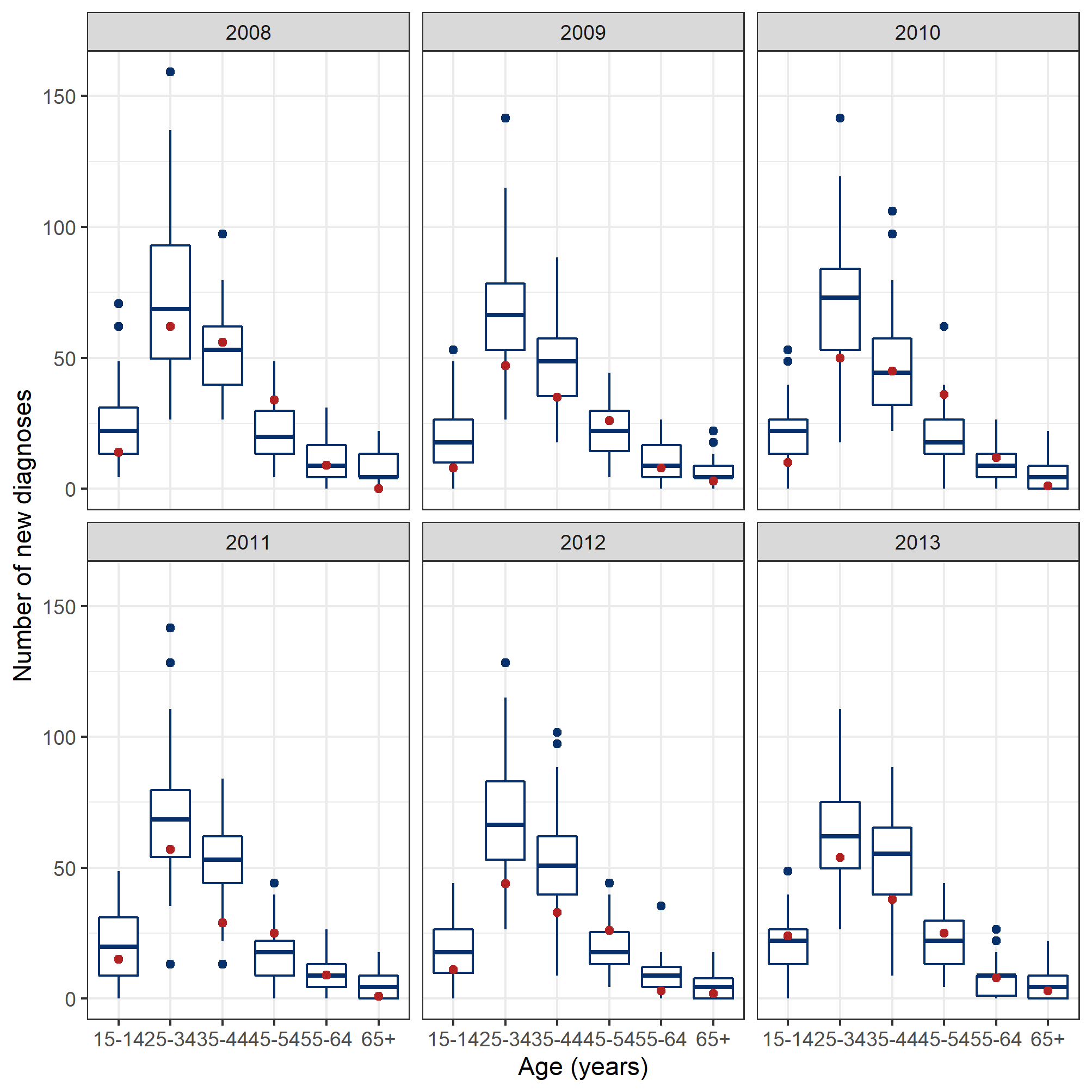
**Figure 35:** Simulated number of new diagnoses from 2008-2013. The boxplots represent the simulation outcomes and the red dots represent the surveillance data ^98^ (from the *Institut national de santé publique du Québec*). The model outcomes were stratified by 6 10-year age groups.

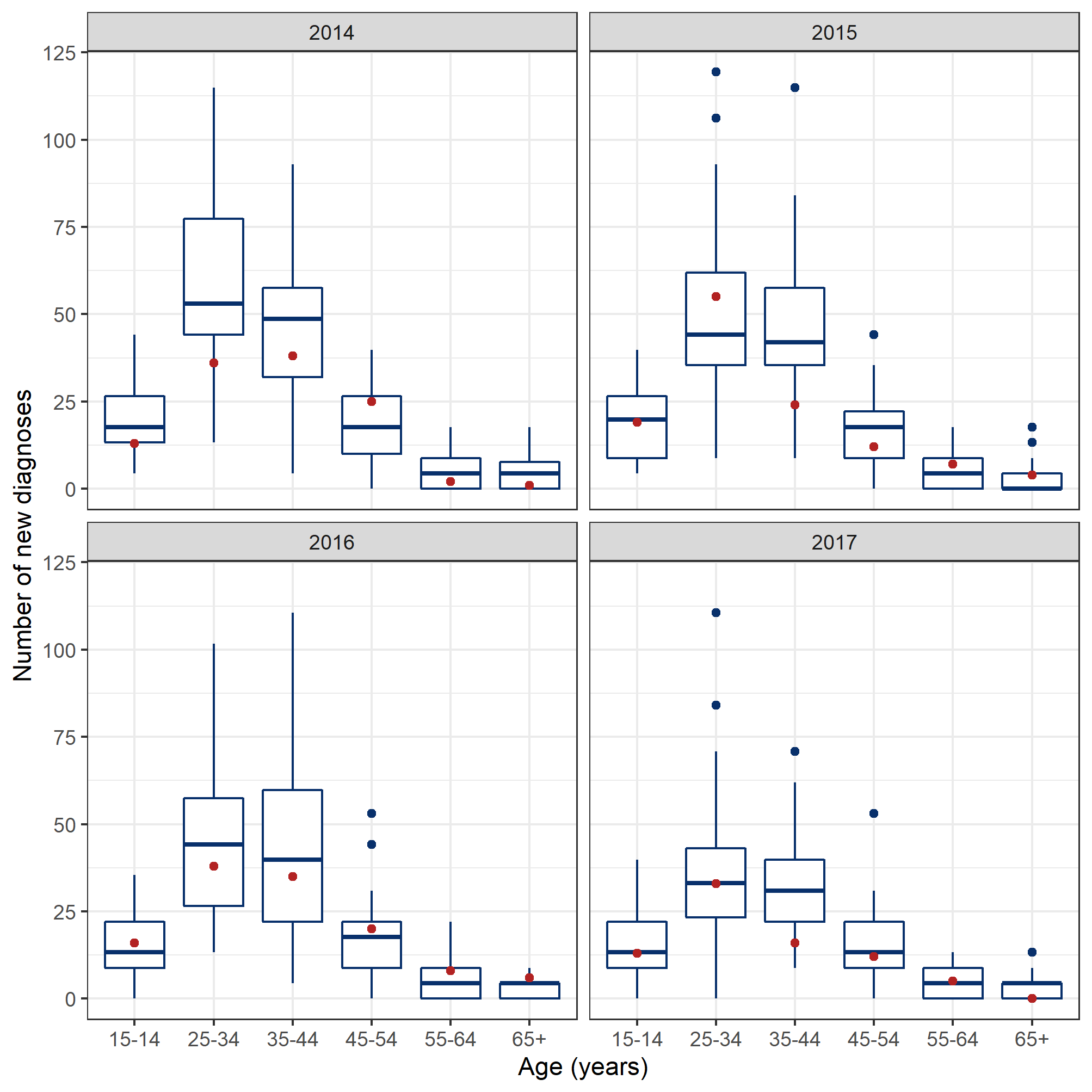

**Figure 36:** Simulated number of new diagnoses from 2014-2017. The boxplots represent the simulation outcomes and the red dots represent the surveillance data ^98^ (from the *Institut national de santé publique du Québec*). The model outcomes were stratified by 6 10-year age groups.

### 8 Bibliography

1.	Sullivan P. The disease that transformed medicine: AIDS turns 20. *CMAJ: Canadian Medical Association Journal.* 2002;166(6):789.

2.	Public Health Agency of Canada. *HIV and AIDS in Canada: Surveillance report to December 31, 2014.* Ottawa: Minister of Public Works and Government Services Canada;2015.

3.	Public Health Agency of Canada. *HIV in Canada - 2017 surveillance highlights.* 2018.

4.	Public Health Agency of Canada. *Summary: Estimates of HIV incidence, prevalence and Canada’s progress on meeting the 90-90-90 HIV targets, 2016.* 2018.

5.	Hogg RS, Heath K, Lima VD, et al. Disparities in the burden of HIV/AIDS in Canada. *PLoS One.* 2012;7(11):e47260.

6.	Public Health Agency of Canada. *HIV/AIDS Epi Updates: National HIV prevalence and incidence estimates for 2011.*: Centre for Communicable Diseases and Infection Control, Public Health Agency of Canada;2014.

7.	Institut national de santé publique du Québec. Programme de surveillance de l’infection par le virus de l’immunodéficience humaine (VIH) au Québec: rapport annuel 2019. 2020.

8.	Baggaley RF, White RG, Boily MC. HIV transmission risk through anal intercourse: systematic review, meta-analysis and implications for HIV prevention. *Int J Epidemiol.* 2010;39(4):1048-1063.

9.	Haddad N, Robert A, Weeks A, Popovic N, Siu W, Archibald C. HIV in Canada-surveillance report, 2018. *Can Commun Dis Rep.* 2019;45(12):304-312.

10.	Trussell J, Fineberg HV, Stoto MA, et al. *No time to lose: getting more from HIV prevention.* Washington (DC): National Academies Press (US); 2001.

11.	Ross DA. Behavioural interventions to reduce HIV risk: what works? *AIDS.* 2010;24(pS4-S14).

12.	Marshall BD, Paczkowski MM, Seemann L, et al. A complex systems approach to evaluate HIV prevention in metropolitan areas: preliminary implications for combination intervention strategies. *PLoS One.* 2012;7(9):e44833.

13.	*R: A language and environment for statistical computing* [computer program]. R Foundation for Statistical Computing,; 2019.

14.	Eddelbuettel D, Balamuta JJ. Extending R with C++: A brief introduction to Rcpp. *PeerJ Preprints.* 2017;5:e3188v3181.

15.	Canadian AIDS Treatment Information Exchange (CATIE). A history of HIV/AIDS. 2018; <https://www.catie.ca/en/world-aids-day/history>. Accessed May 6, 2019.

16.	World Health Organization. *Antiretroviral therapy for HIV infection in adults and adolescents: Recommendations for a public health approach.* 2006.

17.	Doherty M, Ford N, Vitoria M, Weiler G, Hirnschall G. The 2013 WHO guidelines for antiretroviral therapy: evidence-based recommendations to face new epidemic realities. *Curr Opin HIV AIDS.* 2013;8(6):528-534.

18.	Panel on Antiretroviral Guidelines for Adults and Adolescents. Guidelines for the use of antiretroviral agents in HIV-1-infected adults and adolescents. In: Department of Health and Human Services.; 2013.

19.	Panel on Antiretroviral Guidelines for Adults and Adolescents. *Guidelines for the use of antiretroviral agents in HIV-1-infected adults and adolescents.* 2016.

20.	Thaker H, Snow M. HIV viral suppression in the era of antiretroviral therapy. *Postgraduate medical journal.* 2003;79(927):36--42.

21.	Rodger AJ, Cambiano V, Bruun T, et al. Sexual activity without condoms and risk of HIV transmission in serodifferent couples when the HIV-positive partner is using suppressive antiretroviral therapy. *JAMA.* 2016;316(2):171-181.

22.	Thomas R, Galanakis C, Vezina S, et al. Adherence to post-exposure prophylaxis (PEP) and incidence of HIV seroconversion in a major North American cohort. *PLoS One.* 2015;10(11):e0142534.

23.	Smith DK, Grohskopf LA, Black RJ, et al. Antiretroviral postexposure prophylaxis after sexual, injection-drug use, or other nonoccupational exposure to HIV in the United States: Recommendations from the US Department of Health and Human Services. *Morbidity and Mortality Weekly Report: Recommendations and Reports.* 2005;54(2):1-20.

24.	Baeten JM, Haberer JE, Liu AY, Sista N. Preexposure prophylaxis for HIV prevention: where have we been and where are we going? *J Acquir Immune Defic Syndr.* 2013;63 Suppl 2:S122-129.

25.	Buchbinder SP. Maximizing the benefits of HIV preexposure prophylaxis. *Topics in antiviral medicine.* 2018;25(4):138.

26.	Public Health Agency of Canada. Survey of HIV, viral hepatitis, STIs and associated risk behaviours among Quebec men who have sex with men. 2009; <http://argusquebec.ca/english/index.html>. Accessed 2019-080-06.

27.	Public Health Agency of Canada. *M-Track: Enhanced surveillance of HIV, sexually transmitted and blood-borne infections, and associated risk behaviours among men who have sex with men in Canada. Phase 1 report.* 2011.

28.	Engage Montreal. What is Engage Montreal? <https://www.engage-men.ca/montreal/>. Accessed 08-12, 2019.

29.	Lambert G, Cox J, Messier-Peet M, Apelian H, Moodie E, Engage research team. *Engage Montréal, Portrait of the sexual health of men who have sex with men in Greater Montréal, Cycle 2017-2018, Highlights.* 2019.

30.	Greenwald ZR, Maheu-Giroux M, Szabo J, et al. Cohort profile: l'Actuel pre-exposure prophylaxis (PrEP) cohort study in Montreal, Canada. *BMJ Open.* 2019;9(6):e028768.

31.	Camirand H, Issouf T, Jimmy B. *L’Enquête québécoise sur la santé de la population, 2014-2015: pour en savoir plus sur la santé des Québécois. Résultats de la deuxième édition.* Québec: Institut de la statistique du Québec.

32.	Statistics Canada. *Census Canada 1986.* 1987.

33.	Statistics Canada. *1991 Census of population.* 1991.

34.	Statistics Canada. *1981 Census of Canada, census divisions and subdivisions - population, occupied private dwellings, private households, census families in private households.* 1981.

35.	Statistics Canada. *1976 Census of Canada. Volume 2, population, demographic characteristics.* 1976.

36.	Institut de la statistique du Québec. *Estimations de la population des MRC selon le groupe d'âge et le sexe, âge médian et âge moyen, Québec, 1er juillet 1996 à 2018.* 2020.

37.	Statistics Canada. Table 13-10-0114-01 Life expectancy and other elements of the life table, Canada, all provinces except Prince Edward Island. <https://www150.statcan.gc.ca/t1/tbl1/en/tv.action?pid=1310011401>. Accessed 06-06-2019.

38.	Loue S. Defining men who have sex with men (MSM). In: Loue S. (eds) Health issues confronting minority men who have sex with men. In: Springer, New York, NY; 2008.

39.	Truong HM, O'Keefe KJ, Pipkin S, et al. How are transgender women acquiring HIV? Insights from phylogenetic transmission clusters in San Francisco. *AIDS.* 2019.

40.	Scheim A, Santos G, Arreola S, et al. Inequities in access to HIV prevention services for transgender men: results of a global survey of men who have sex with men. *Journal of the International AIDS Society.* 2016;19.

41.	Becasen JS, Denard CL, Mullins MM, Higa DH, Sipe TA. Estimating the prevalence of HIV and sexual behaviors among the US transgender population: A systematic review and meta-analysis, 2006–2017. *American Journal of Public Health* 2019;109:e1-e8.

42.	Millett GA, Peterson JL, Flores SA, et al. Comparisons of disparities and risks of HIV infection in black and other men who have sex with men in Canada, UK, and USA: a meta-analysis. *The Lancet.* 2012;380(9839):341-348.

43.	Morris M, Kretzschmar M. Concurrent partnerships and the spread of HIV. *Aids.* 1997;11(5):641--648.

44.	Hertog S. Heterosexual behavior patterns and the spread of HIV/AIDS: the interacting effects of rate of partner change and sexual mixing. *Sex Transm Dis.* 2007;34(10):820-828.

45.	Wong NS, Kwan TH, Tsang OTY, et al. Pre-exposure prophylaxis (PrEP) for MSM in low HIV incidence places: should high risk individuals be targeted? *Sci Rep.* 2018;8(1):11641.

46.	Sood N, Wagner Z, Jaycocks A, Drabo E, Vardavas R. Test-and-treat in Los Angeles: a mathematical model of the effects of test-and-treat for the population of men who have sex with men in Los Angeles County. *Clin Infect Dis.* 2013;56(12):1789-1796.

47.	Remis RS, Alary M, Liu J, Kaul R, Palmer RW. HIV transmission among men who have sex with men due to condom failure. *PLoS One.* 2014;9(9):e107540.

48.	Cohen MS, Chen YQ, McCauley M, et al. Antiretroviral therapy for the prevention of HIV-1 transmission. *N Engl J Med.* 2016;375(9):830-839.

49.	Centers for Disease Control and Prevention. HIV treatment as prevention. 2019; <https://www.cdc.gov/hiv/risk/art/index.html>. Accessed 2019-10-25.

50.	Wilson DP, Law MG, Grulich AE, Cooper DA, Kaldor JM. Relation between HIV viral load and infectiousness: a model-based analysis. *The Lancet.* 2008;372(9635):314-320.

51.	Hollingsworth TD, Anderson RM, Fraser C. HIV-1 transmission, by stage of infection. *J Infect Dis.* 2008;198(5):687-693.

52.	Vergis EN, Mellors JW. Natural history of HIV-1 infection. *Infectious Disease Clinics.* 2000;14(4):809-825.

53.	Cori A, Pickles M, van Sighem A, et al. CD4+ cell dynamics in untreated HIV-1 infection: overall rates, and effects of age, viral load, sex and calendar time. *AIDS.* 2015;29(18):2435-2446.

54.	Johnson LF, May MT, Dorrington RE, et al. Estimating the impact of antiretroviral treatment on adult mortality trends in South Africa: A mathematical modelling study. *PLoS Med.* 2017;14(12):e1002468.

55.	Avenir Health. Spectrum system of policy models. 2019; <https://www.avenirhealth.org/software-spectrum.php>. Accessed 2019-10-25.

56.	Brennan DJ, Welles SL, Miner MH, Ross MW, Rosser BS, Team PC. HIV treatment optimism and unsafe anal intercourse among HIV-positive men who have sex with men: findings from the positive connections study. *AIDS Education & Prevention.* 2010;22(2):126--137.

57.	Stephenson J, Imrie J, Davis M, et al. Is use of antiretroviral therapy among homosexual men associated with increased risk of transmission of HIV infection? *Sexually Transmitted Infections.* 2003;79(1):7-10.

58.	Remien RH, Halkitis PN, O'Leary A, Wolitski RJ, Gomez CA. Risk perception and sexual risk behaviors among HIV-positive men on antiretroviral therapy. *AIDS Behav.* 2005;9(2):167-176.

59.	Ostrow DE, Fox KJ, Chmiel JS, et al. Attitudes towards highly active antiretroviral therapy are associated with sexual risk taking among HIV-infected and uninfected homosexual men. *Aids.* 2002;16(5):775--780.

60.	George C, Alary M, Otis J, et al. Nonnegligible increasing temporal trends in unprotected anal intercourse among men who have sexual relations with other men in Montreal. *JAIDS Journal of Acquired Immune Deficiency Syndromes.* 2006;41(3):365--370.

61.	Cox J, Beauchemin J, Allard R. HIV status of sexual partners is more important than antiretroviral treatment related perceptions for risk taking by HIV positive MSM in Montreal, Canada. *Sex Transm Infect.* 2004;80(6):518-523.

62.	Otis J, McFadyen A, Haig T, et al. Beyond condoms: risk reduction strategies among gay, bisexual, and other men who have sex with men receiving rapid HIV testing in Montreal, Canada. *AIDS Behav.* 2016;20(12):2812-2826.

63.	Schechter M, Do Lago RF, Mendelsohn AB, et al. Behavioral impact, acceptability, and HIV incidence among homosexual men with access to postexposure chemoprophylaxis for HIV. *JAIDS Journal of Acquired Immune Deficiency Syndromes.* 2004;35(5):519-525.

64.	Jain S, Oldenburg CE, Mimiaga MJ, Mayer KH. Subsequent HIV infection among men who have sex with men who used non-occupational post-exposure prophylaxis at a Boston community health center: 1997-2013. *AIDS Patient Care STDS.* 2015;29(1):20-25.

65.	Donnell D, Mimiaga MJ, Mayer K, Chesney M, Koblin B, Coates T. Use of non-occupational post-exposure prophylaxis does not lead to an increase in high risk sex behaviors in men who have sex with men participating in the EXPLORE trial. *AIDS Behav.* 2010;14(5):1182-1189.

66.	Kimmig P. Sensitivity and specificity of test results in HIV. *Das Offentliche Gesundheitswesen.* 1990;52(8-9):419 - 424.

67.	Alary M, Josette C. Risk factors for HIV seropositivity among people consulting for HIV antibody testing: a pilot surveillance study in Quebec. *Canadian Medical Association Journal.* 1990;143(1):25.

68.	Archibald CP, Jayaraman GC, Major C, Patrick DM, Houston SM, Sutherland D. Estimating the size of hard-to-reach populations: a novel method using HIV testing data compared to other methods. *Aids.* 2001;15:S41--S48.

69.	Xia Y, Greenwald ZR, Milwid RM, et al. Pre-exposure prophylaxis uptake among men who have sex with men who used non-occupational post-exposure prophylaxis: a longitudinal analysis of attendees at a large sexual health clinic in Montreal (Canada). *J Acquir Immune Defic Syndr.* 2020.

70.	L'Actuel Centre de Santé Sexuelle. Post-exposure prophylaxis (PEP). <https://cliniquelactuel.com/>. Accessed 06-03-2019.

71.	Alexander TS. Human immunodeficiency virus diagnostic testing: 30 years of evolution. *Clin Vaccine Immunol.* 2016;23(4):249-253.

72.	FDA. HIV/AIDS historical time line 1981-1990. 20181; <https://www.fda.gov/patients/hiv-timeline-and-history-approvals/hivaids-historical-time-line-1981-1990>. Accessed 05-28-2019, 2019.

73.	Kasaie P, Pennington J, Shah MS, et al. The impact of preexposure prophylaxis among men who have sex with men: An individual-based model. *J Acquir Immune Defic Syndr.* 2017;75(2):175-183.

74.	Sucharitakul K, Boily MC, Dimitrov D, Mitchell KM. Influence of model assumptions about HIV disease progression after initiating or stopping treatment on estimates of infections and deaths averted by scaling up antiretroviral therapy. *PLoS One.* 2018;13(3):e0194220.

75.	Johnson WD, O'Leary A, Flores SA. Per-partner condom effectiveness against HIV for men who have sex with men. *AIDS.* 2018;32(11):1499-1505.

76.	Lévy JJ, Dupras A, Doras M, Perreault M, Frigault L-R. Styles de relations et conduites sexuelles d’hommes francophones montréalais qui ont des rapports sexuels avec d’autres hommes. *Service social.* 1996;45(1).

77.	Dufour A, Alary M, Otis J, et al. Risk behaviours and HIV infection among men having sexual relations with men: baseline characteristics of participants in the Omega Cohort Study, Montreal, Quebec, Canada. *Canadian Journal of Public Health.* 2000;91(5):345--349.

78.	Ministère de la Santé et des Services sociaux. *La prophylaxie préexposition au virus de l’immunodéficience humaine: Guide pour les professionnels de la santé du Québec.* Gouvernement du Québec,; November 2017.

79.	Ministère de la Santé et des Services sociaux. *La prophylaxie préexposition au virus de l’immunodéficience humaine: Guide pour les professionnels de la santé du Québec.* Gouvernement du Québec,; January 2019.

80.	Beyrer C, Baral SD, van Griensven F, et al. Global epidemiology of HIV infection in men who have sex with men. *The Lancet.* 2012;380(9839):367-377.

81.	Millett GA, Flores SA, Marks G, Reed JB, Herbst JH. Circumcision status and risk of HIV and sexually transmitted infections among men who have sex with men: a meta-analysis. *Jama.* 2008;300(14):1674--1684.

82.	Yuan T, Fitzpatrick T, Ko N-Y, et al. Circumcision to prevent HIV and other sexually transmitted infections in men who have sex with men: a systematic review and meta-analysis of global data. *The Lancet Global Health.* 2019;7(4):e436-e447.

83.	Patel P, Borkowf CB, Brooks JT, Lasry A, Lansky A, Mermin J. Estimating per-act HIV transmission risk: a systematic review. *AIDS.* 2014;28(10):1509-1519.

84.	Boily M-C, Baggaley RF, Wang L, et al. Heterosexual risk of HIV-1 infection per sexual act: systematic review and meta-analysis of observational studies. *Lancet Infect Dis.* 2009;9(2):118-129.

85.	Prabhu S, Harwell JI, Kumarasamy N. Advanced HIV: diagnosis, treatment, and prevention. *The Lancet HIV.* 2019;6(8):e540-e551.

86.	Molina JM, Capitant C, Spire B, et al. On-demand preexposure prophylaxis in men at high risk for HIV-1 infection. *N Engl J Med.* 2015;373(23):2237-2246.

87.	Centers for Disease Control and Prevention. *Updated guidelines for antiretroviral postexposure prophylaxis after sexual, injection drug use, or other nonoccupational exposure to HIV— United States, 2016.* 2016.

88.	Toni T, Welch D, Strelkowa N, Ipsen A, Stumpf MP. Approximate Bayesian computation scheme for parameter inference and model selection in dynamical systems. *J R Soc Interface.* 2009;6(31):187-202.

89.	Institut national de santé publique du Québec. Programme de surveillance de l’infection par le virus de l’immunodéficience humaine (VIH) au Québec;rapport annuel 2016. 2017.

90.	Institut national de santé publique du Québec. Programme de surveillance de l’infection par le virus de l’immunodéficience humaine (VIH) au Québec;rapport annuel 2014. 2015.

91.	Institut national de santé publique du Québec. Programme de surveillance de l’infection par le virus de l’immunodéficience humaine (VIH) au Québec;rapport annuel 2015. 2016.

92.	Institut national de santé publique du Québec. Programme de surveillance de l’infection par le virus de l’immunodéficience humaine (VIH) au Québec;rapport annuel 2017. 2019.

93.	Institut national de santé publique du Québec. Programme de surveillance de l’infection par le virus de l’immunodéficience humaine (VIH) au Québec;rapport annuel 2013. 2014.

94.	Yang Q, Ogunnaike-Cooke S, Halverson J, et al. Estimated national HIV incidence rates among key subpopulations in Canada, 2014. Paper presented at: 25th Annual Canadian Conference on HIV/AIDS Research (CAHR)2016; Winnipeg, Canada.

95.	Remis RS, Alary M, Otis J, et al. No increase in HIV incidence observed in a cohort of men who have sex with other men in Montreal. *AIDS.* 2002;16(8):1183-1185.

96.	Sante et Services Sociaux Quebec. Portrait des infections transmissibles sexuellement et par le sang (ITSS) au Québec Année 2012 (et projection 2013). 2013.

97.	Zang X, Krebs E, Min JE, et al. Development and Calibration of a Dynamic HIV Transmission Model for 6 US Cities. *Med Decis Making.* 2020;40(1):3-16.

98.	Institut national de santé publique du Québec. INSPQ Public health expertise and reference centre. <https://www.inspq.qc.ca/en/institute/about-us>. Accessed 08-12, 2019.
